# Lipid Trajectories Improve Risk Models for Alzheimer’s Disease and Mild Cognitive Impairment

**DOI:** 10.1101/2024.09.27.24314494

**Authors:** Bruce A. Chase, Roberta Frigerio, Chad J. Yucus, Smita Patel, Demetrius Maraganore, Alan R. Sanders, Jubao Duan, Katerina Markopoulou

**Author notes:** Corresponding Author: Bruce A. Chase, PhD, Information Technology, Endeavor Health Suite 160, 4901 Searle Parkway, Skokie, IL 60077 USA, / cell. **Funding:** This development of the structured clinical documentation support toolkits within the electronic health record was funded by the Agency for Healthcare Research and Quality (R01HS024057) to DM and the generous support of the Auxiliary of NorthShore University HealthSystem (now Endeavor Health). The analyses presented in this work were funded by the National Institute on Aging (R01AG081374 and R01AG063175) to JD.

## Abstract

To assess the relationship between lipids and cognitive dysfunction, we retrospectively analyzed blood-lipid levels in clinically well-characterized individuals with stable mild cognitive impairment (MCI) or Alzheimer’s disease (AD) over the decade prior to first cognitive symptoms. In this case/control cohort study, AD and MCI cases were diagnosed using DSM-IV criteria; MCI cases had not progressed to dementia for ≥5 years; and controls were propensity matched to cases at age of symptom onset (MCI: 116 cases, 435 controls; AD: 215 cases, 483 controls). Participants were grouped based on longitudinal trajectories and quintile of variability independent of the mean (VIM) for total cholesterol, HDL-C, LDL-C, non-HDL-C and ln(triglycerides). Models for the risk of cognitive dysfunction evaluated trajectory and VIM groups, *APOE* genotype, polygenic risk scores (PRS) for AD and lipid levels, age, comorbidities, and longitudinal correlates of blood-lipid concentrations. Lower HDL-C trajectories (OR = 3.8, 95% CI = 1.3–11.3) and the lowest VIM quintile of non-HDL-C (OR = 2.2, 95% CI = 1.3–3.0) were associated with higher MCI risk. Lower HDL-C trajectories (OR = 3.0, 95% CI = 1.6–5.7) and the lowest VIM quintile of total cholesterol (OR = 2.4, 95% CI = 1.5–3.9) were associated with higher AD risk. The inclusion of lipid-trajectory and VIM groups improved risk-model predictive performance independent of *APOE* genotype or PRS for AD and lipid levels. These results provide an important real-world perspective on the influence of lipid metabolism and blood-lipid levels on the development of stable MCI and AD.

## Introduction

Alzheimer’s Disease (AD) is the most prevalent neurodegenerative disorder, affecting almost 58 million people globally, including an estimated 6 million in the United States (1,2). Mild cognitive impairment (MCI) typically precedes the onset of dementia, but may not progress to AD; some individuals with MCI either do not experience further cognitive decline or revert to normal cognition (3).

Mounting genetic and biological evidence suggests a central role for lipid dysregulation in AD and other neurodegenerative conditions (4–13). Several genetic risk factors for AD implicate pathways in lipid metabolism (14). *APOE* ε4, the strongest genetic risk factor for both early and late onset AD, is associated with increased plasma cholesterol levels, primarily due to an increase in low-density lipoproteins (LDL-C). The *APOE*-ε4 allele disrupts cholesterol homeostasis in astrocytes (15) and other cell types (16–18) and is associated with accumulation of lipid droplets in glia and tau phosphorylation in neurons (19). Variants of ApoB, the main lipoprotein in LDL-C, are associated with early onset AD (20) and overexpression of ApoB in mice results in hyperlipidemia, neurodegeneration, increased APP expression, and amyloid plaques (21,22). Genome-wide association studies (GWAS) have also identified associations between AD risk and several genes involved in brain cholesterol metabolism/transport, including ATP-binding cassette, subfamily A, member 7 (*ABCA7*) (23), Phospholipase-D3 (*PLD*3), clusterin/apolipoprotein J (*CLU*), sortilin-related receptor 1 (*SORLI*) (24), phosphatidylinositol binding clathrin assembly protein (*PICALM*) and phospholipase C-gamma 2 (*PLCG2*) (25). Polygenic risk scores (PRS) for AD developed from these GWAS are associated with high cholesterol levels, and a high PRS for LDL-C is associated with AD (26). Several risk factors for AD, including aging, education level, and socioeconomic status are also associated with changes in serum lipid levels (27). AD and MCI risk has also been associated with specific types of triglycerides (28) and with dietary fat intake (29).

Some studies investigating the relationship between blood-lipid levels and AD risk have suggested that high total cholesterol and LDL-C levels during mid-life are associated with increased risk of AD (30–32), including early onset AD (20), and that high total cholesterol levels are associated with an increased cerebral burden of Aβ (33). Conversely, some studies suggest that low total cholesterol and LDL-C levels in later life are associated with an increased risk of dementia (34,35), although other studies showed no such correlation (36). High cholesterol in young and middle-aged people is associated with obesity, metabolic syndrome, and diabetes, which are AD risk factors; cholesterol levels at older ages may reflect loss of body mass and fat, as well as use of cholesterol-lowering medicines. Whereas higher serum triglyceride levels in older adults may reduce AD risk and slow cognitive decline, higher levels in mid-life are associated with abnormal Aβ and tau levels two decades later (32,37,38), although these risks may be modified by *APOE* genotype (39). Other reports have suggested that increased variability in metabolic parameters (40), including lipid levels (41,42), over time is associated with increased dementia risk.

Brain lipids are involved in modulation of membrane-protein interactions, including with AD-related molecules such as Aβ or tau (43–45) and disruption of brain lipid homeostasis is associated with cognitive deficits and neurodegeneration (46). Dysregulation of cholesterol metabolism has been implicated in the production and clearance of beta-amyloid (Aß) (47), the processing of amyloid precursor protein (APP) (48), and tau phosphorylation (49). Excess cholesterol in AD is converted to cholesterol esters (CE), which, along with other lipids, accumulate in glial cells as lipid droplets (50). CEs may increase Aß production, through altered trafficking of APP (45), and mediate tau phosphorylation (51). Although peripheral and brain cholesterol levels are independent due to the blood brain barrier (BBB), which is impermeable to cholesterol, serum cholesterol levels may affect brain cholesterol metabolism. In the brain and peripheral circulation, cholesterol is oxidized to oxysterol derivatives, 24-hydroxycholesterol (24-HC) and 27-hydroxycholesterol (27-HC), respectively, which can permeate the BBB. High levels of brain 27-HC (reflecting high plasma cholesterol levels), but not 24-HC, may accelerate cognitive deficits (52).

These studies demonstrate the complex interrelationship between lipid levels, lipid-associated comorbidities, metabolism, aging, and genetic factors that influence the risk of cognitive dysfunction. However, the extent to which blood-lipid levels—in particular longitudinal levels before and after disease onset and their interplay with genetic or non-genetic factors—predict AD and MCI risk remains elusive.

We sought to investigate the relative strength of risk factors for AD and MCI through a retrospective analysis of blood lipids in well-characterized patient cohorts from a real-world, community practice setting. We examined how two distinct measures of lipid level variation—relative levels or variability independent of the mean—during the pre-symptomatic period were associated with risk of AD or stable MCI and evaluated the magnitude of this risk relative to genetic risk.

## METHODS

### Study Overview

In this retrospective, clinical and genetic case/control study, we evaluated serum cholesterol and lipid levels over the ten years before cognitive symptom onset in cases and in propensity-matched controls (matched at age of symptom onset) without neurodegenerative disease. We used two distinct approaches to characterize blood-lipid levels (HDL-C, LDL-C, total cholesterol, non-HDL-C, triglycerides) in MCI and AD case/control cohorts. First, we used non-parametric group-based trajectory models to identify sets of patients sharing similar longitudinal lipid trajectories over time. Second, we calculated the variability independent of the mean (VIM), a measure that is robust to heteroscedasticity and uncorrelated with mean lipid levels. In unadjusted analyses, we assessed the magnitude of the association between trajectory group or lipid VIM quintile with MCI or AD risk and examined other patient characteristics associated with longitudinal lipid levels, MCI, or AD. We then used covariate-adjusted logistic regression to model the contribution of each lipid trajectory group and VIM quintile to AD or MCI risk in the context of genetic factors, patient demographics, and patient medical history.

### Study Patients, Diagnostic Criteria, Case/Control Matching and Data Elements

We analyzed data obtained from patients enrolled in *The DodoNA project: DNA Predictions to Improve Neurological Health* (DodoNA) (N = 12,498), which investigates the contribution of genetic risk to progression and outcomes in ten neurological conditions (53–66) and in an at-risk *Brain Health* cohort (61) (**Table 1**). Each DodoNA cohort is followed longitudinally using structured clinical documentation support (SCDS) tools embedded in the electronic health record (EHR) to record patient data at the point of care. The DodoNA project was approved by the NorthShore University HealthSystem Institutional Review Board (EH10-139, April 2011) and the studies in this work abide by the Declaration of Helsinki principles.

**Table 1.**
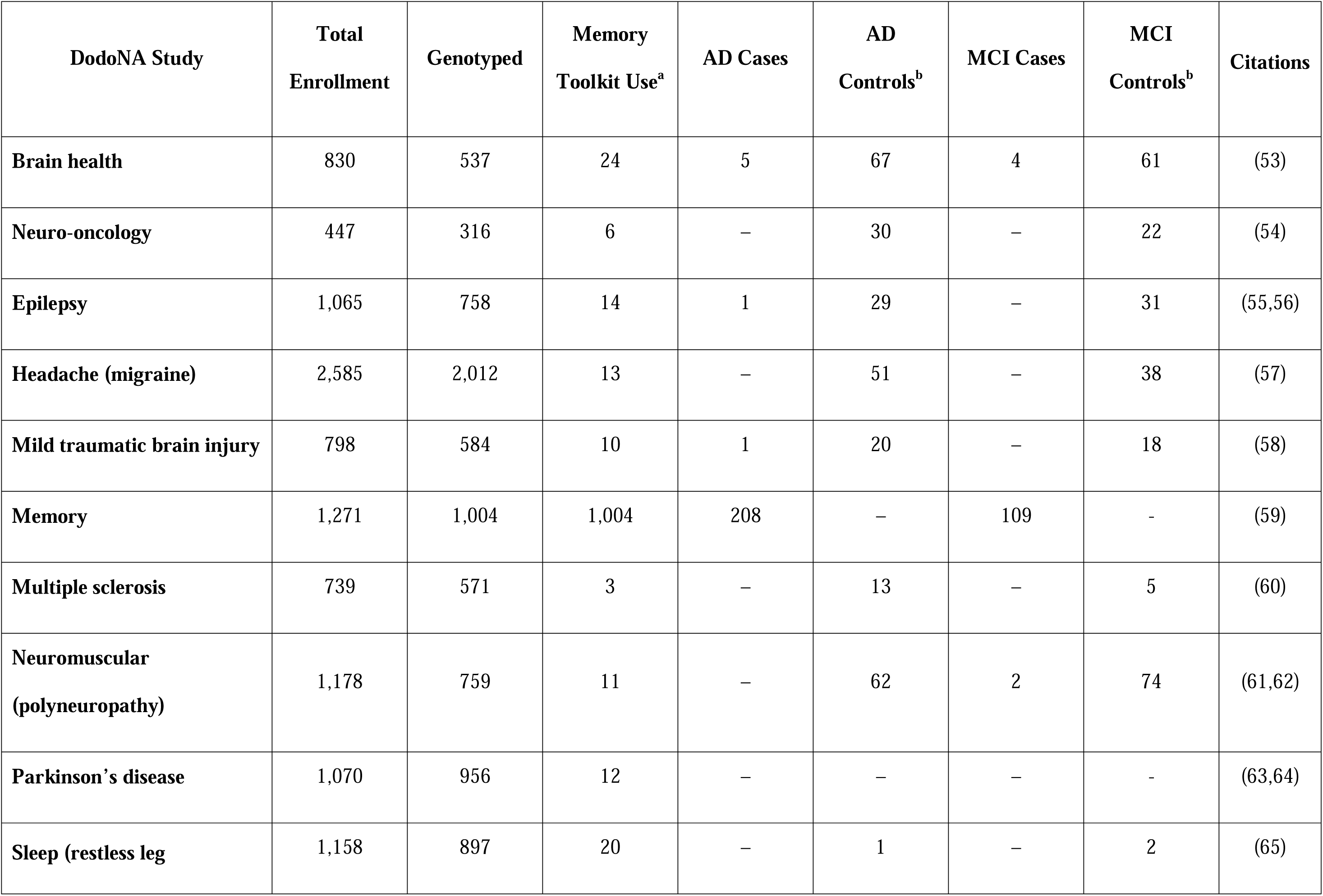
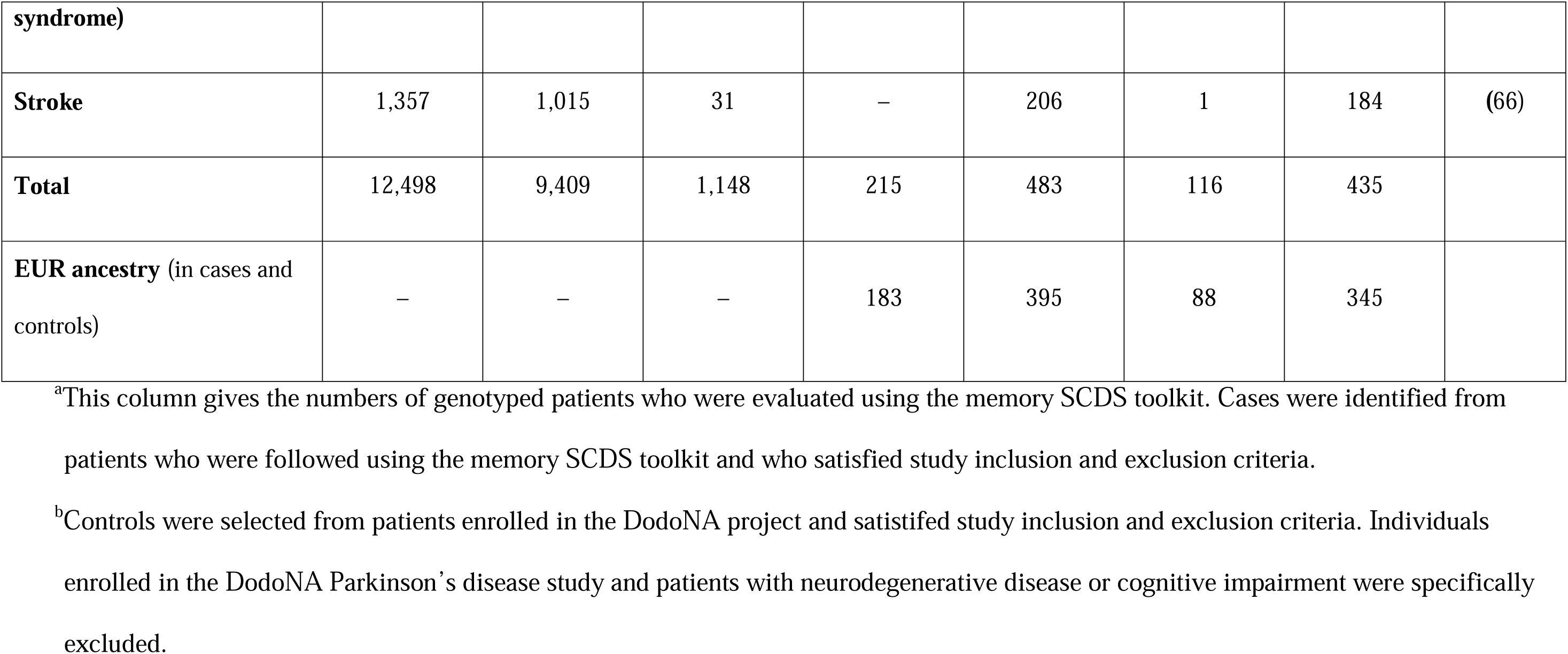
Participants by DodoNA-project study.

We analyzed clinical data collected from April 2011 through February 24, 2024, and data on lipid measurements, International Classification of Disease (ICD) codes and social history stored in our Electronic Data Warehouse (EDW) from its inception in 2003 through February 24, 2024. Cases with AD or MCI were selected from DodoNA patients followed using the memory toolkit who were diagnosed using DSM-IV criteria. The data elements collected in the memory toolkit are summarized in **Table S1**. Patients who had at least three annual serum lipid measurements, in different years, during the period up to 10 years before the year of symptom onset in cases or age at match in controls (i.e., within the 11-year period that included the decade prior and year of symptom onset or age match) were included in the study. Patients with fewer than three such lipid measurements were excluded (**Figure 1**).

**Figure 1:**
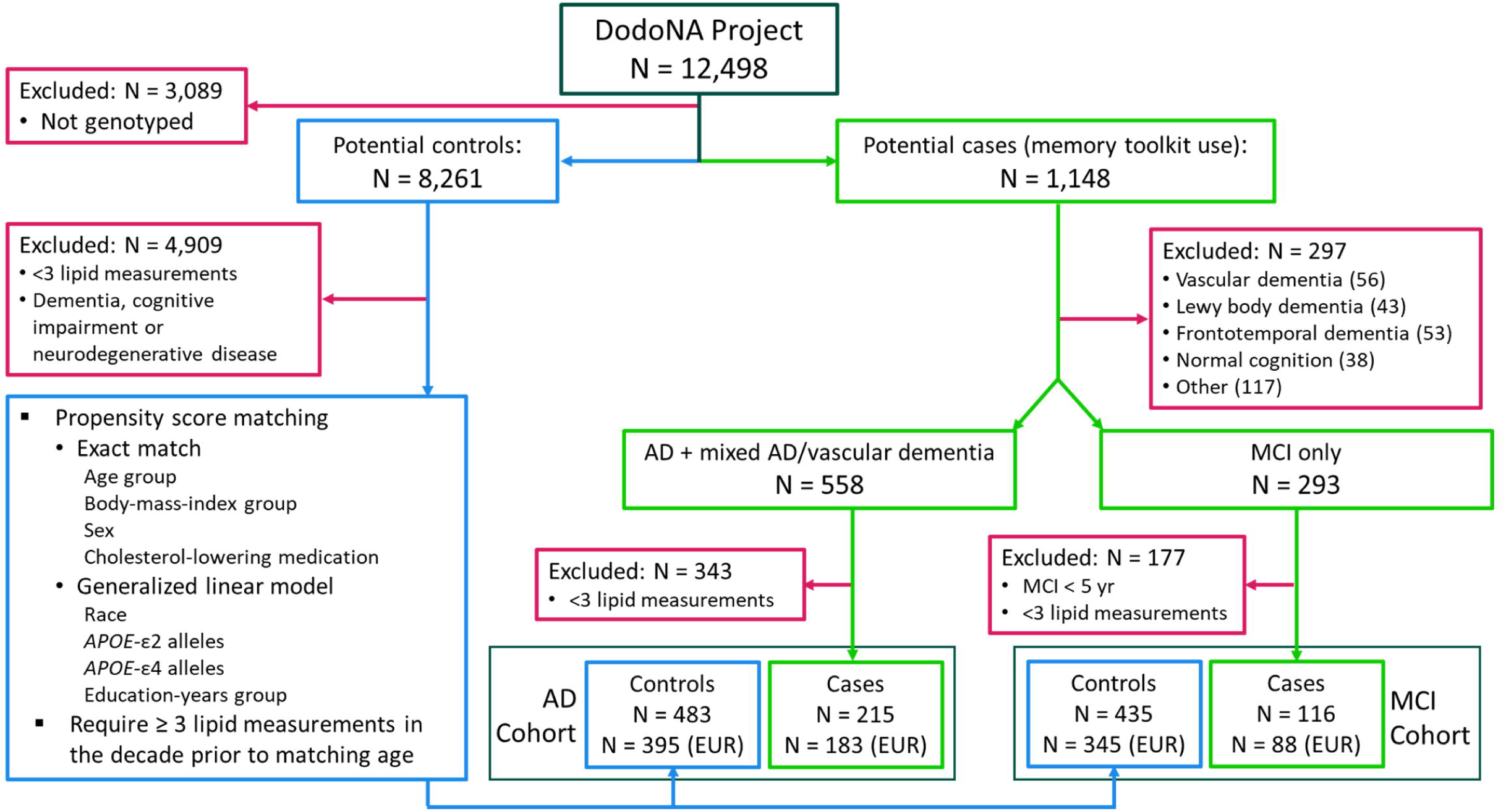
Selection of AD and MCI case/control cohorts. Case/control cohorts were selected from genotyped patients enrolled in the DodoNA project. Cases were selected from patients who were followed with a structured clinical documentation toolkit for memory disorders. AD (AD or mixed AD/vascular dementia) and MCI diagnoses utilized DSM-IV criteria. Exclusion criteria for cases were: normal cognition upon evaluation, an inconclusive diagnosis, a diagnosis of vascular dementia (only), Lewy body dementia, frontotemporal dementia, or cognitive impairment or dementia associated with another condition (parkinsonism, stroke, normal pressure hydrocephalus, cortical basal degeneration, progressive supranuclear palsy, stroke, alcohol use, end-stage-renal disease, subdural hematoma, B12 deficiency, a psychiatric disorder, or radiation therapy). AD and MCI cases were excluded if they had < 3 blood-lipid measurements in the decade prior to first symptom onset, and MCI cases were included only if MCI did not progress to dementia for at least five years. Controls were selected from patients enrolled in studies of other neurological diseases or a brain-health cohort (see **Table 1**). Controls did not have an ICD code for dementia or cognitive impairment (see Methods), any diagnosis of neurodegenerative disease, and had ≥ 3 blood lipid measurements. Cases were propensity-score matched to controls based on the criteria shown and controls were retained only if they had ≥ 3 blood-lipid measurements in the decade prior to their matching age. The number of cases and controls in each cohort, and the number with European ancestry (EUR), is shown.

Blood lipid concentrations were determined using standard clinical assays. LDL-C and non-HDL-C concentrations were calculated as described (67) if they were not recorded in the EDW. Triglyceride values were log transformed to normalize their distribution. Data on blood cholesterol-lowering medications—any statin, gemfibrozil, fenobibrate, clofibrate, bempedoic, ezetimibe, cholestyramine, colestipol, colesivelam, alirocumab, or evolucumab—were analyzed over the same period. We inferred that a patient was using this type of medication at the time of blood lipid measurement if, before or during that time, a medication order was listed as “Sent”, “Dispensed” or “Verified”, but not “Discontinued”, unless the order was replaced by another medication. Use of cholesterol-lowering medication was included as a time-dependent covariate in group-based trajectory models and as a criterion for matching cases and controls; however, lipid data were not adjusted for medication use.

We analyzed data from two cohorts (**Figure 1**): (i) The AD cohort included cases [N = 215 (183 European ancestry (EUR))] and matched controls [N = 483 (395 EUR)]. Cases had a diagnosis of AD or mixed AD and vascular dementia. Patients with only vascular dementia or with non-AD dementia (e.g., Pick’s disease, frontotemporal dementia, Lewy Body dementia, etc.) were excluded. (ii) The MCI cohort included cases [N = 116 (88 EUR)] and matched controls [N = 435 (345 EUR)]. Cases had a diagnosis of MCI with a duration of five or more years and with no diagnosis of another dementia or neurodegenerative disease. Since more than half of MCI cases progress to dementia within five years (68–70), we used the inclusion criterion of 5+ years with MCI to focus on individuals with stable MCI who were less likely to convert to dementia.

For both cohorts, controls had no neurologist diagnosis of any cognitive or neurodegenerative disorder or any International Classification of Diseases (ICD) codes for dementia, cognitive impairment or neurodegenerative disease (ICD-9: 290–, 294–, 323–, 330–, 331–, 332–, 333.4, 437–, 797–; ICD-10: F01–, F02–, F03–, F06–, G20–, G30–, G31–, I69–, R41–) in the EHR. Individuals included in the AD and MCI cohorts were enrolled from different DodoNA studies (see **Table 1**).

To generate case and control groups with similar covariate distributions for both cohorts, we propensity matched cases at the age of first cognitive symptoms to controls using the *R* package *MatchIt* (71). Exact matches were used for sex, age group (50–59, 60–64, 65–72, 73–77, 78–81, 82–85, 86–98), body mass index (BMI) group (<18.5, 18.5-24.9, 25-29.9, 30-34.9, 35-40, >40 kg/m^2^), and use of cholesterol-lowering medication. Nearest matches, selected using a generalized linear model (logistic regression), were used for patient-reported race, number of *APOE-*ε2 and -ε4 alleles, and patient-reported years-of-education (<12, 12, 12-15, ≥16 years).

The following were recorded as binomial variables: whether patients were smokers or moderate-to-heavy alcohol users (>7 drinks/week) prior to the matching age, based on patient-reported social history information stored in the EDW; presence of an ICD9 or ICD10 codes for hypertension (401– to 403–, 405–, I10– to I16–) or atherosclerosis (414–, 437–, I25–); cerebrovascular disease (437, I67.3, I69–), diabetes (249–, 250–, E09– to E11–, E13–), ischemic heart disease/myocardial infarction (21.29, 410– to 414–, 429.2, I20– to I25–, I31.2, I51.3, I51, I82.9, I97–, I99.8, Q21–, Q89.9, Z13.6), or malignant neoplasm (140– to 239–, C00–– to C96––, D00–– to D49––, H47–).

### Genotyping, Imputation and Calculation of Polygenic Risk Score

Single nucleotide polymorphism (SNP) genotyping on an Affymetrix Axiom™ array with custom content and quality control was performed as previously described (63). We used the Michigan Imputation Server (72) for genotype imputation, phasing, and ancestry estimation [Haplotype Reference Consortium reference panel, Version r1.1 2016, Eagle v2.4, *r*^2^ = 0.8], and to calculate PRS from the PGS Catalog (73). In models for the longitudinal correlates of lipid levels and in the models for AD and MCI risk, we evaluated PRS for AD (PGS004034, PGS004092, both of which include variants in the *APOE* region) (74), triglycerides (PGS003147, PGS003152), total cholesterol (PGS003142, PGS003137), LDL-C (PGS003037, PGS003032), HDL-C (PGS002957) (75), and non-HDL-C (PGS002782) (76). The PRS were standardized (mean = 0, standard deviation = 1) within each case/control cohort to facilitate a comparison of their effect sizes.

### Statistical Analyses

Differences between cases and controls for demographic and medical history data elements, lipid trajectory groups, and VIM quintiles were evaluated using χ^2^ tests for categorical variables and Kruskal-Wallis tests for continuous variables. A Bonferroni adjustment was applied to tests of association of AD or MCI with lipid trajectory groups and VIM quintiles and to tests of differences in cohort characteristics across these groups. The distribution of PRS in cases and controls was visualized using a kernel-density plot, obtained using an Epanechnikov kernel function. A Kolmogorov-Smirnov test evaluated the equality of PRS distribution in cases and controls. Inter-PRS correlations were assessed using the Pearson correlation method. Statistical analyses were performed using Stata BE 18 and R.

### Longitudinal Correlates of Lipid Levels

Prior to developing group-based trajectory models, we used multilevel mixed-effects models to address whether our patient population showed similar longitudinal correlates of lipid concentrations as those described by Duncan *et al.* (67) and to evaluate associations between genetic factors and longitudinal lipid concentrations. By analyzing data from the same individual over time as well as across individuals, we were able to estimate the impact of covariates on temporal patterns at the study-sample level as well as the overall pattern of change over time at the individual level. These models included a random intercept and random slope for age (to account for different initial lipid levels in each participant and a different slope for age). The models used maximum likelihood estimation as a maximization criterion with an unstructured covariance matrix that allowed for correlation between the random slopes and intercepts for age. Covariates initially evaluated were selected based on those identified in (67) for which data were available in the EHR. They included age, sex, cholesterol-lowering medication use (obtained as described above), BMI, smoker, years of education (dummy coded as <12 yr, 12 yr, >12 years), moderate-to-heavy alcohol use; ICD-code based diagnoses (as described above) of hypertension, cerebrovascular disease, diabetes ischemic heart disease or myocardial infarction, atherosclerosis, and malignant neoplasm; interactions between age and sex, BMI, diabetes, hypertension, cerebrovascular disease, or atherosclerosis; interactions between alcohol use and smoking; and interactions between sex and diabetes or lipid-lowering medication use. We also included genotypes at *APOE*-ε4 and ε2, *PICALM* AA, and *CLU* CC, the set of PRS for AD and blood lipids described above and 10 genetic principal components as covariates. Since these analyses were aimed at informing the selection of covariates that would later be used in AD and MCI risk models, we evaluated these covariates in all patients, not only those with EUR ancestry. Lipid data were not adjusted for use of cholesterol-lowering medications; however, medication use was included as a covariate. The best fitting model was identified using backwards selection and the Akaike Information Criterion.

### Group-Based Trajectory Models

Group-based lipid trajectory models were developed using the semi-parametric, model-based clustering method described by Jones and Nagin (77). This method allows for the inclusion of individuals with multiple measurements that contain gaps, i.e., that are not sequential, and estimates the probability of group membership for each individual. The models were produced by fitting longitudinal lipid concentration data to finite-mixture models using the *traj* plug-in in Stata/BE 18.0 (78). We evaluated model fit with two to five trajectory groups for each dependent variable (lipid measurement) over time (years prior to year of first symptom in cases or age match in controls); use of cholesterol-lowering medication was included as a time-varying covariate. The link function between time and the lipid measurement variable was censored-normal (a tobit model). The best-fitting models were identified through an iterative process using the fit-criteria assessment plot developed by Klijn *et al.* (79), as described previously (80). Briefly, the ‘best’ model was identified by fitting single- to five-group models with 0- to 4^th^-order polynomials and comparing model fit based on Bayesian Information Criteria (BIC), Akaike Information Criterion (AIC), and model-maximized likelihood. Following Nagin (81), we required each trajectory group to have an average posterior probability of group assignment ≥ 0.85 and odds of correct classification ≥ 5.0. To maintain clinical relevance, we also required that each group included at least 5% of a case/control cohort.

### Variability Independent of the Mean (VIM)

VIM assesses variation over time that is uncorrelated to the mean and is robust to heteroscedasticity. To address whether lipid variability, over the decade prior to cognitive symptom onset, was associated with disease risk, we calculated VIM for each lipid species, as described by Moser *et al.* (42), using the same lipid measurements that were used to develop group-based trajectory models. We then tested associations between lipid VIM quintile and AD or MCI onset.

### AD and MCI Risk Models

Covariate-adjusted logistic regression risk models were developed using backward selection, the Akaike information criterion (AIC), and cluster-robust standard errors that allowed for intra-matching-group correlation. Models with PRS as covariates (*case ∼ covariates + lipid-trajectory groups + 10 genetic principal components + PRS*) only included individuals with European ancestry, as most PRS were developed in European populations, while models with *APOE* genotypes as covariates (*case ∼ covariates + lipid-trajectory groups + APOE* genotypes) included entire case/control cohorts. Covariates considered in all models included age and matching weights as continuous variables, and female sex and >12 years of education as binomial variables. Other binomial variables included smoking, moderate-to-heavy alcohol use, hypertension, cerebrovascular disease, diabetes, atherosclerosis, or ischemic heart disease, prior to the year of first symptom (for cases) or age-match (for controls). Other categorical variables included trajectory group assignments and VIM quintiles for non-HDL-C, HDL-C, ln(triglycerides), LDL-C, and total cholesterol. PRS were evaluated as continuous variables.

When developing risk models, we considered previously reported correlations between some AD and lipid PRS (52) and expected correlations between PRS for certain lipids by evaluating inter-PRS correlations. In the population studied, different PRS for the same trait (AD, triglycerides, or total cholesterol LDL) showed very strong positive correlations (*r* = 0.8−1.0) and different PRS for total cholesterol, LDL-C and non-HDL-C showed strong to very strong positive correlations (*r* = 0.6−0.9); however, both AD PRS showed a very weak positive correlation (*r* = 0.0−0.2) with PRS for different lipids. For PRS showing strong positive correlations, we evaluated each PRS individually to identify the best model fit. For example, compared to PGS003142_total cholesterol_ and PGS004034_AD_, PGS003137_total cholesterol_ and PGS004092_AD_, respectively, gave better model fit (by AIC) and higher pseudo-R^2^, so these PRS were retained in the final models.

Models considering *APOE* genotypes evaluated *APOE-*ε4 and *APOE-*ε2, each relative to the no-*APOE-*ε4 or no-*APOE-*ε2 allele class, and race, dummy-coded as Caucasian, Black/African American or Asian. Models also evaluated interactions of age with atherosclerosis, hypertension, cerebrovascular disease, PGS004092_AD_, and HDL-C trajectory groups. All models passed link and Hosmer-Lemeshow goodness-of-fit tests. The area under the receiver operator characteristic (AUROC) was calculated for risk models using *roccomp* in *Stata* BE 18.5. We report the Bonferroni-corrected *p* value of a test of equality of area under the curves produced by each set of related models: (1) baseline covariates such as age, sex, genetic principal components; (2) baseline covariates plus lipid groups; (3) baseline covariates plus genetic factors (PRS or *APOE* genotypes); and (4) baseline covariates, lipid groups, and genetic factors.

## RESULTS

### Patient Demographics and Lipid Measurements

Demographics, *APOE* allele status, and medical history for the matched cases and controls are summarized in **Table 2**; genetic ancestry is summarized in **Table S2**. Our study population was more than 91% Caucasian and approximately 50% female. Cases and controls were similar except for three characteristics: (i) As expected, *APOE*-ε4 homozygotes and heterozygotes were more frequent in both AD and MCI cases than in controls; (ii) although over 60% of controls and cases in both cohorts had at least 16 years of education, the AD cohort had more cases with 12 years of education than controls; (iii) the AD cohort had more cases with history of cerebrovascular disease than controls, and the MCI cohort had fewer cases with a history of atherosclerosis than controls.

**Table 2.** Demographics in AD and MCI Case/Control Cohorts.

| Characteristic<br>*type of match<sup>a</sup>* |  | AD Cohort |  |  | MCI Cohort |  |  |
| --- | --- | --- | --- | --- | --- | --- | --- |
|  |  | Cases<br>N = 215 | Controls<br>N = 483 | *p<sup>b</sup>* | Cases<br>N = 116 | Controls<br>N = 435 | *p<sup>b</sup>* |
| **Sex**<br>*exact* | male | 103 (47.9) | 240 (49.7) | 0.664 | 58 (50) | 233 (53.6) | 0.495 |
|  | female | 112 (52.1) | 243 (50.3) |  | 58 (50) | 202 (46.4) |  |
| **Age group** *exact* | 50 – 59 | 7 (3.3) | 19 (3.9) | 0.154 | 3 (2.6) | 20 (4.6) | 0.873 |
|  | 60 – 64 | 3 (1.4) | 10 (2.1) |  | 1 (0.9) | 3 (0.7) |  |
|  | 65 – 72 | 36 (16.7) | 116 (24.0) |  | 28 (24.1) | 108 (24.8) |  |
|  | 73 – 77 | 42 (19.5) | 97 (20.1) |  | 28 (24.1) | 113 (26.0) |  |
|  | 78 – 81 | 48 (22.3) | 112 (23.2) |  | 23 (19.8) | 83 (19.1) |  |
|  | 82 – 85 | 44 (20.5) | 69 (14.3) |  | 21 (18.1) | 59 (13.6) |  |
|  | 86 – 98 | 35 (16.3) | 60 (12.4) |  | 12 (10.3) | 49 (11.3) |  |
| **BMI group**<br>*exact* | < 18.5 | 3 (1.4) | 11 (2.3) | 0.943 | 3 (2.6) | 8 (1.8) | 0.878 |
|  | 18.5 – 24.9 | 91 (42.3) | 195 (40.4) |  | 37 (31.9) | 152 (34.9) |  |
|  | 25 – 29.9 | 90 (41.9) | 205 (42.4) |  | 48 (41.4) | 186 (42.8) |  |
|  | 30 – 34.9 | 27 (12.6) | 62 (12.8) |  | 17 (14.7) | 60 (13.8) |  |
|  | 35 – 40 | 4 (1.9) | 10 (2.1) |  | 9 (7.8) | 22 (5.1) |  |
|  | > 40 | 0 (0) | 0 (0) |  | 2 (1.7) | 7 (1.6) |  |
| **Blood-**<br>**cholesterol**<br>**lowering**<br>**medication use**<br>*exact* | Used<sup>b</sup> | 158 (73.5) | 365 (75.6) | 0.558 | 89 (76.7) | 348 (80.0) | 0.439 |
| **Race**<br>*nearest* | Caucasian | 203 (94.4) | 442 (91.5) | 0.592 | 107 (92.2) | 397 (91.3) | 0.814 |
|  | Black /<br>African-American | 5 (2.3) | 15 (3.1) |  | 2 (1.7) | 10 (2.3) |  |
|  | Asian | 6 (2.8) | 23 (4.8) |  | 5 (4.3) | 24 (5.5) |  |
|  | Other | 1 (0.5) | 3 (0.6) |  | 2 (1.7) | 4 (0.9) |  |
| **APOE ε2 alleles**<br>*nearest* | 0 | 198 (92.1) | 419 (86.8) | 0.082 | 100 (86.2) | 381 (87.6) | 0.586 |
|  | 1 | 17 (7.9) | 60 (12.4) |  | 15 (12.9) | 53 (12.2) |  |
|  | 2 | 0 (0) | 4 (0.8) |  | 1 (0.9) | 1 (0.2) |  |
| **APOE ε4 alleles**<br>*nearest* | 0 | 102 (47.4) | 327 (67.7) | <0.001 | 73 (62.9) | 316 (72.6) | 0.017 |
|  | 1 | 86 (40) | 139 (28.8) |  | 36 (31.0) | 111 (25.5) |  |
|  | 2 | 27 (12.6) | 17 (3.5) |  | 7 (6.0) | 8 (1.8) |  |
| **Years of education**<br>*nearest* | < 12 | 5 (2.3) | 14 (2.9) | 0.026 | 3 (2.6) | 9 (2.1) | 0.387 |
|  | 12 | 45 (20.9) | 58 (12.0) |  | 22 (19.0) | 56 (12.9) |  |
|  | 12-15 | 32 (14.9) | 81 (16.8) |  | 20 (17.2) | 85 (19.5) |  |
|  | ≥ 16 | 133 (61.9) | 330 (68.3) |  | 71 (61.2) | 285 (65.5) |  |
| **Smoking** |  | 1 (0.5) | 11 (2.3) | 0.089 | 3 (2.6) | 9 (2.1) | 0.735 |
| **Moderate-to-heavy alcohol use** |  | 21 (9.8) | 52 (10.8) | 0.691 | 13 (11.2) | 50 (11.5) | 0.931 |
| **Hypertension** |  | 156 (72.6) | 364 (75.4) | 0.433 | 93 (80.2) | 334 (76.8) | 0.437 |
| **Atherosclerosis** |  | 10 (4.7) | 35 (7.2) | 0.197 | 2 (1.7) | 35 (8) | 0.016 |
| **Cerebrovascular disease** |  | 56 (26.0) | 83 (17.2) | 0.007 | 23 (19.8) | 71 (16.3) | 0.372 |
| **Diabetes** |  | 57 (26.5) | 123 (25.5) | 0.771 | 36 (31.0) | 126 (29.0) | 0.664 |
| **Ischemic heart disease/myocardial infarction** |  | 61 (28.4) | 162 (33.5) | 0.176 | 38 (32.8) | 165 (37.9) | 0.305 |
| **Malignant neoplasm** |  | 75 (34.9) | 205 (42.4) | 0.060 | 45 (38.8) | 188 (43.2) | 0.391 |

<sup>a</sup>Cases were propensity matched to controls using a generalized linear model (logistic regression) with exact or nearest-neighbor matching performed using the *MatchIt* package in *R*. The type of match is indicated in italics beneath variables used to match cases and controls.
<sup>b</sup> $\chi^2$ test
<sup>c</sup>at age of symptom onset in cases, or at age at match in controls

Data from a total of 698 (215 cases) and 551 (116 cases) individuals in the AD and MCI case/control cohorts, respectively, were available and included in the analysis. AD and MCI cases had a mean (standard deviation, range) follow up of 6.5 (2.7, 0.2-14) and 7.8 (1.8, 5-13) years, respectively. As shown in **Figure 2**, more than 95% of AD and MCI cases had first symptom onset after age 65. Among AD cases, 11.6% were also diagnosed with vascular dementia and 58.6% had a history of MCI.

**Figure 2:**
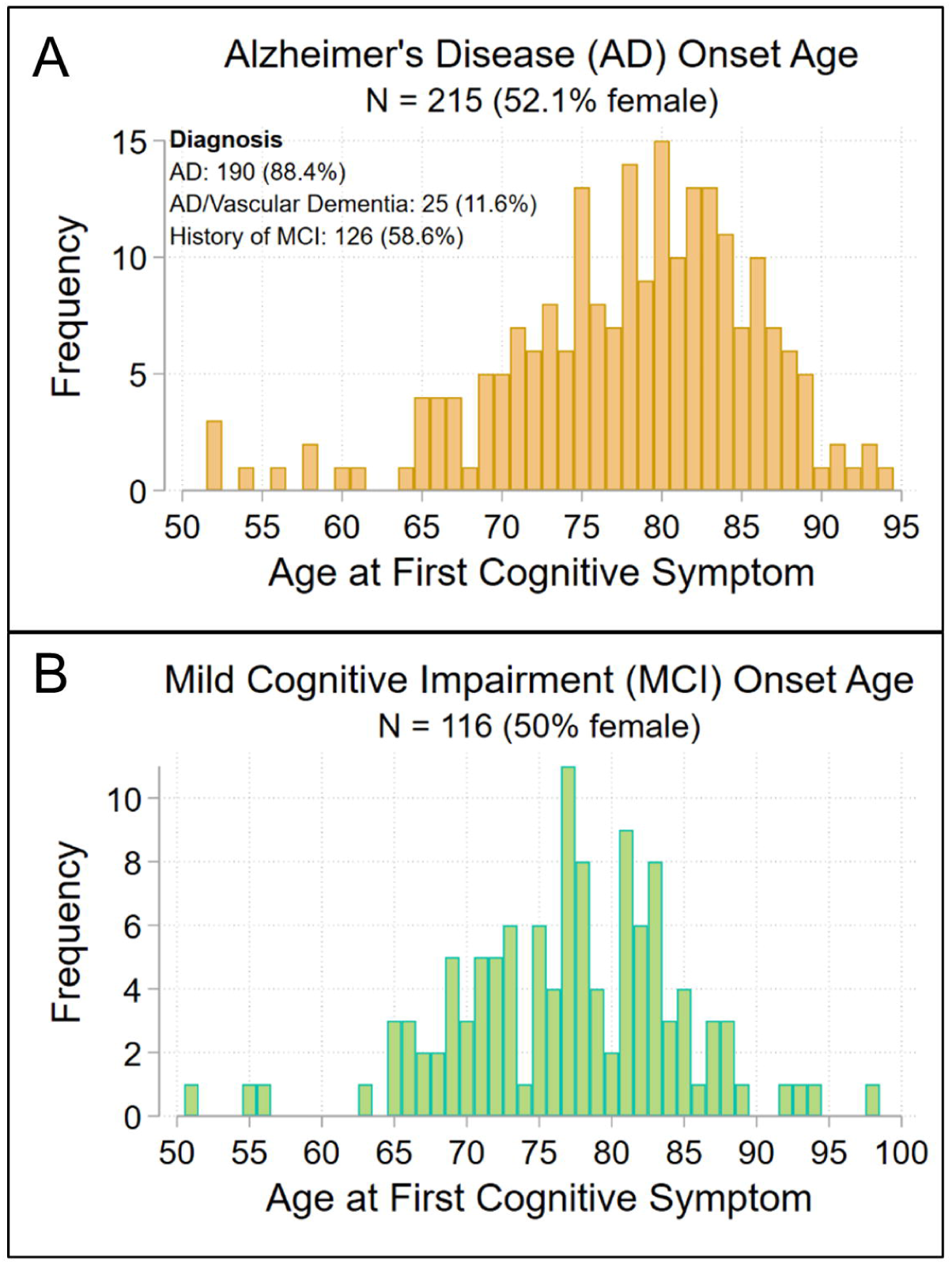
Age at first cognitive symptom in (A) AD and (B) MCI cases. Compare to Table 2.

Since this was a retrospective study, we were restricted to using blood-lipid data available in the EDW. As lipid measurements were not obtained every year for every patient, the number of available lipid measurements varied across patients. AD cases and controls had a median (range) of 5 (3–11) and 7 (3–11) lipid measurements, respectively, with a mean (range) of 63% (41–75%) of patients having lipid measurements each year of the study. MCI cases and controls had 6 (3–11) and 7 (3–11) lipid measurements, respectively, and a mean (range) of 61% (44–72%) of these patients had lipid measurements each year of the study (**Figure S1**).

### Longitudinal Correlates of Lipid Levels

To inform the choice of covariates used in AD and MCI risk models, we assessed longitudinal correlates of lipid levels in the AD and MCI case/control cohorts. For this, we used multilevel mixed-effects models to analyze lipid measurements from the same individual over time as well as across individuals. In these models, we evaluated longitudinal correlates of lipid concentrations similar to those previously identified (67), genotypes at *APOE*, *PICALM* and *CLU*, and PRS for AD and lipid subtypes.

**Table 3** summarizes the associations identified in the best fitting models. In the AD cohort, increasing age was positively associated with HDL-C but negatively associated with total cholesterol, LDL-C, ln(triglycerides), and non-HDL-C. Similar patterns were seen in the MCI cohort for LDL-C and non-HDL-C and, with a smaller effect size, for HDL-C. Except for ln(triglycerides) in the MCI cohort, female sex, or an interaction between female sex and age, was positively associated with each lipid subtype. Cholesterol-lowering medication use was negatively associated with total cholesterol and LDL-C in both cohorts, and with non-HDL-C in the MCI cohort. In the AD cohort, an interaction between age and BMI was negatively associated with HDL-C, and positively associated with ln(triglycerides), while in the MCI cohort, BMI was negatively associated with total cholesterol, HDL-C, LDL-C and positively associated with ln(triglycerides). In the AD cohort, smoking was negatively associated with total cholesterol and LDL-C. In both cohorts, moderate-to-heavy alcohol use was positively associated with HDL-C. Twelve and >12 years of education were negatively associated with total cholesterol, LDL-C and ln(triglycerides) in the AD cohort, and with non-HDL-C in the MCI cohort, while <12 years of education was negatively associated with HDL-C in the MCI cohort. In the AD cohort, when associations between lipids and comorbidities related to cardiovascular health (hypertension, cerebrovascular disease, diabetes, ischemic heart disease/myocardial infarction) were significant, they showed negative associations. This general pattern was also seen in the MCI cohort for diabetes, ischemic heart disease, and atherosclerosis, although the best fitting models included interactions with age. In the MCI cohort, hypertension was positively associated with total cholesterol, LDL-C, ln(triglycerides), and non-HDL-C, but an interaction between hypertension and age showed negative associations with total cholesterol, LDL-C, and non-HDL-C. Prior work has suggested that some negative associations reflect treatment effects. Many, but not all, of these results are similar to those reported by Duncan et al. (54). Differences likely reflect reduced sensitivity due to our smaller sample size, the limited 11-year interval over which we assessed lipid levels, and our older patient population.

**Table 3.**
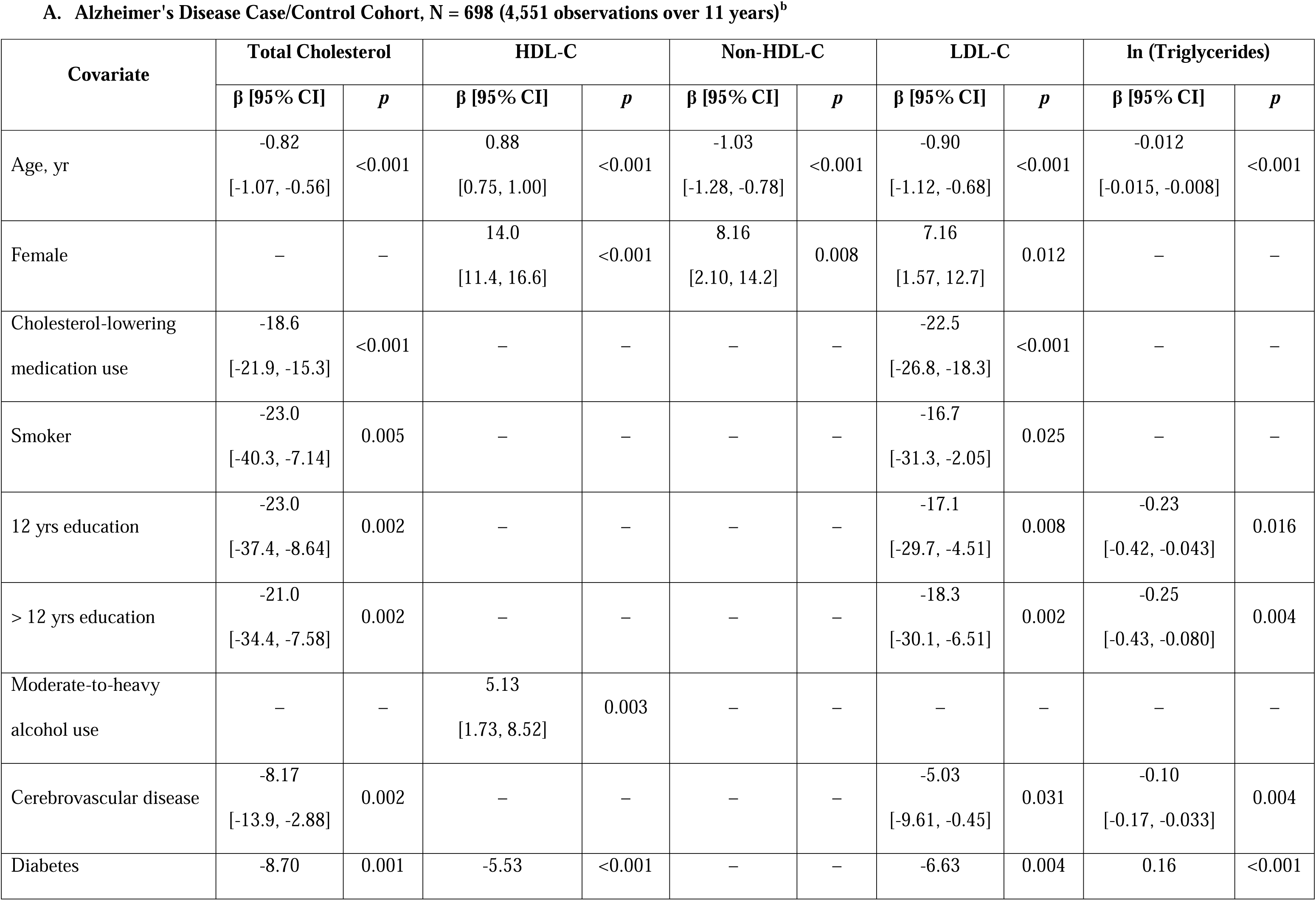
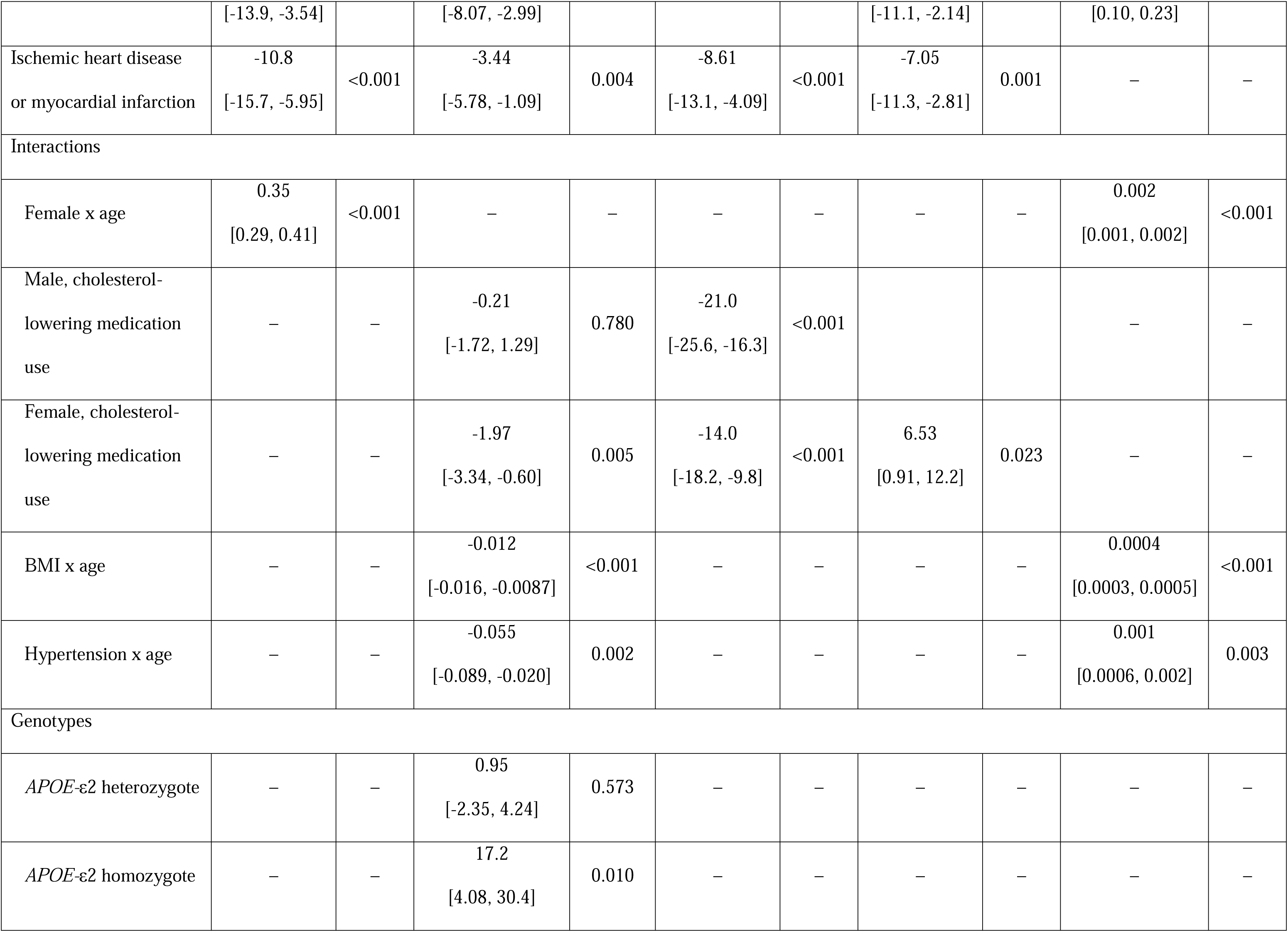
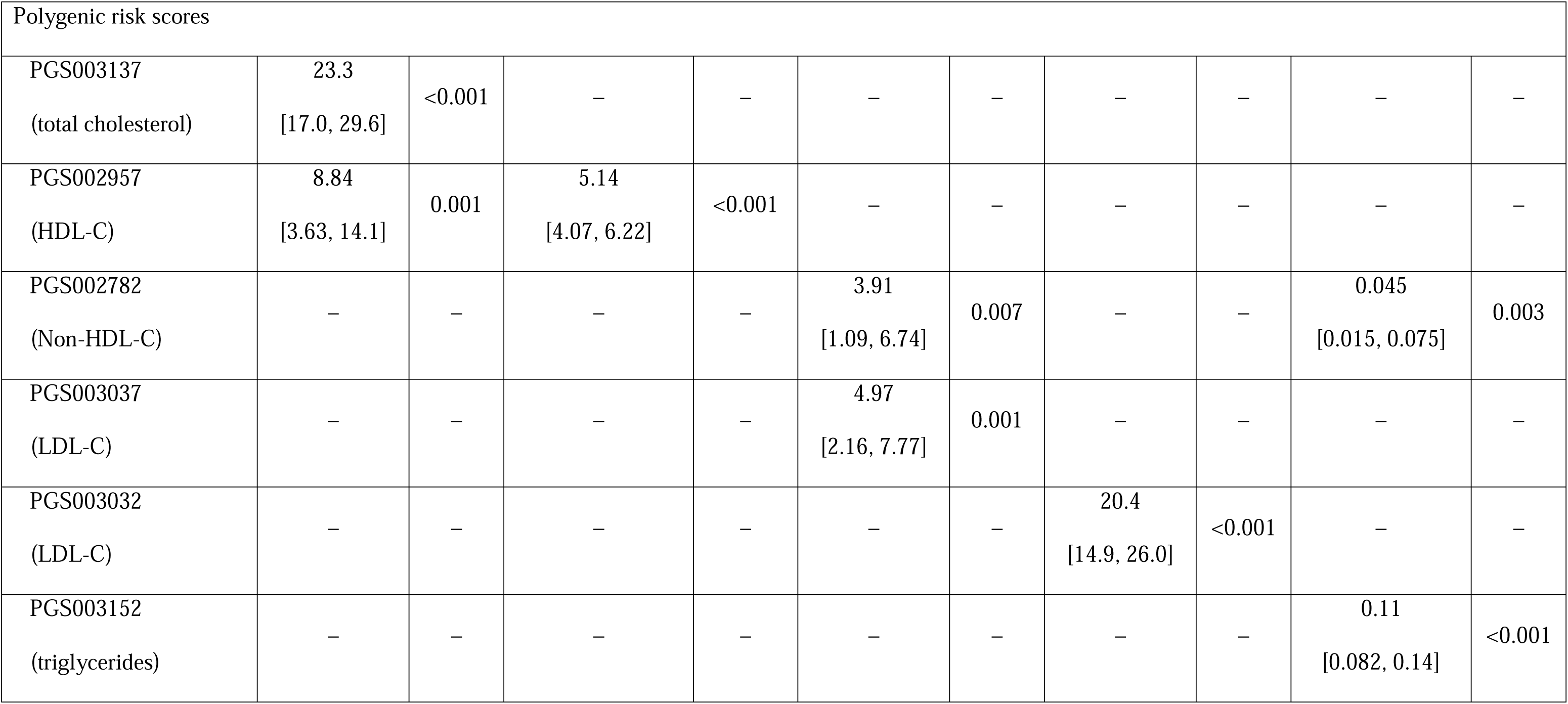
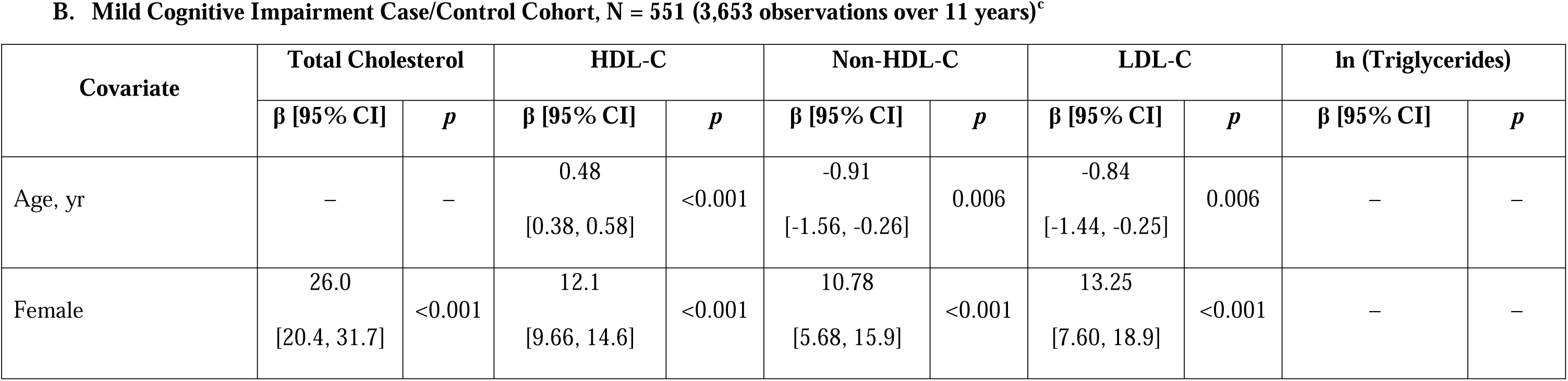
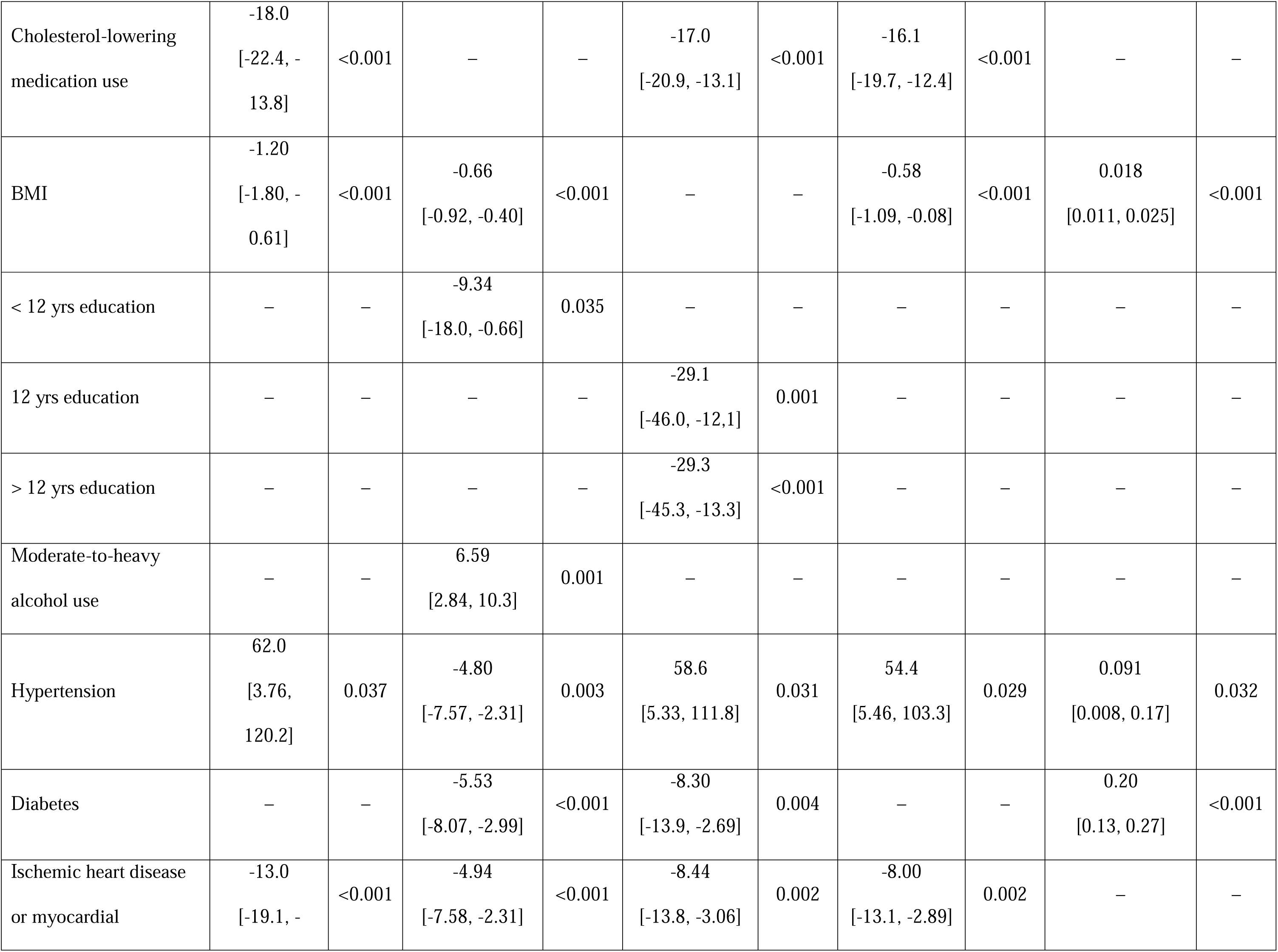
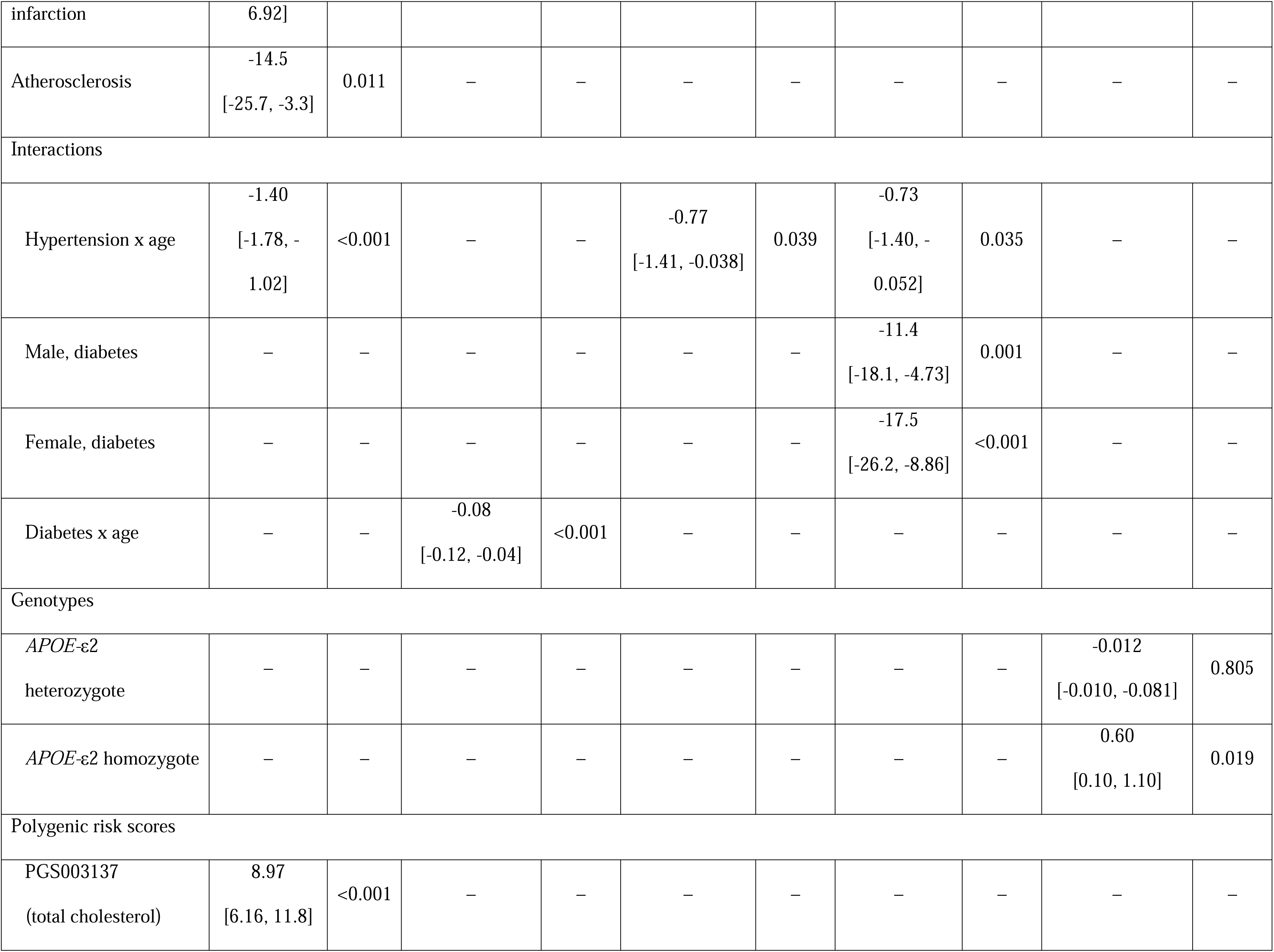
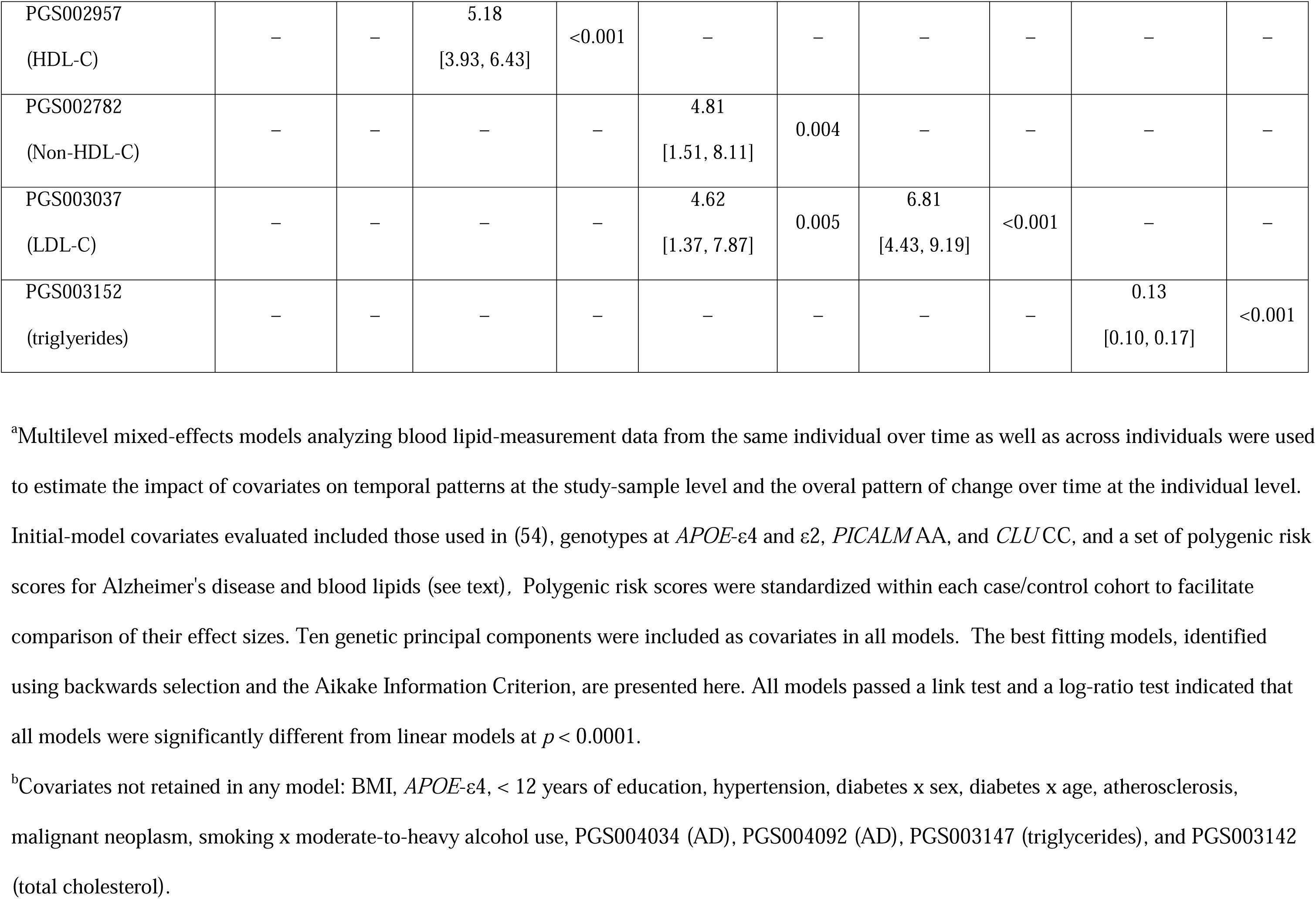
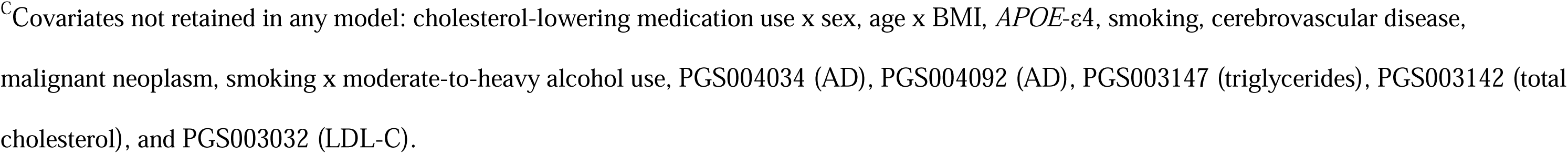
Longitudinal Correlates of Lipid Concentrations^a^.

When genetic associations were evaluated*, APOE*-ε4, *PICALM*, and *CLU* genotypes and PRS for AD failed to show an association with the concentration of any lipid in either cohort. In the AD cohort, *APOE*-ε2 homozygous status was positively associated with HDL-C, and in the MCI cohort, it was positively associated with ln(triglycerides), but with relatively wide 95% CIs that reflect the small number of individuals with this genotype. Most striking were the associations with lipid PRS. In both the AD and MCI cohorts, a PRS for each type of lipid showed robust positive associations with longitudinal concentrations of that lipid.

### Associations of Lipid Trajectories with AD and MCI Cases

In both the AD and MCI case/control patient cohorts, good fits for all lipid subtypes were achieved with three trajectory-group models (**Figure 3**, **Table 4**). Although four-group models showed a better fit for some lipid subtypes, some groups in those models had fewer than 5% of patients, so three trajectory-group models were used in all analyses to ensure clinical relevance. In all models, trajectory group 3, corresponding to the highest levels of lipid subtype, also had the smallest number of patients and the widest 95% confidence intervals (CIs) (**Figure 4**).

**Figure 3:**
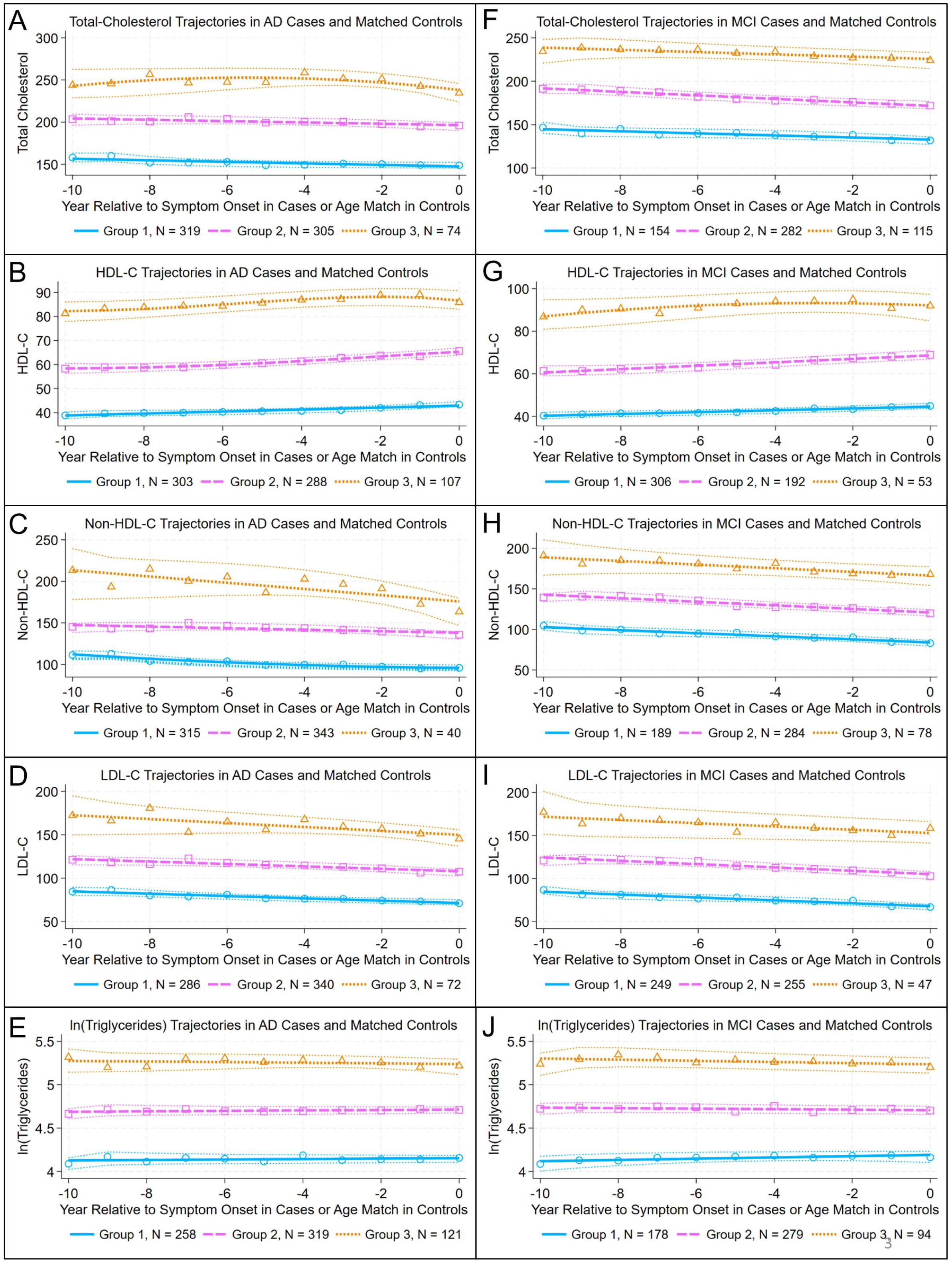
Lipid trajectories in AD and MCI case/control cohorts. Controls were propensity matched (see text and **Table 2** for details) with cases at age of first symptom onset. Total cholesterol and lipid subtype measurements collected over the decade prior to the year of first symptom onset (cases) and matching age (controls) were fit to group-based trajectory models that were developed using blood-cholesterol lowering medication use as a time-dependent covariate. The plots show the best-fitting trajectory for each group for mean lipid values at each year (Group 1: solid lines with open circles; Group 2: long-dashed lines with open squares; Group 3: short-dashed lines with open triangles). For each trajectory, 95% confidence intervals (polynomial-fit) are indicated (dotted lines). Trajectories are shown for total cholesterol (**A, F**); HDL-C (**B**, **G**); non-HDL-C (**C**, **H**); LDL-C, (**D**, **I**); and ln(triglycerides), (**E**, **J**) for the AD, MCI case/control cohorts, respectively. Compare to **Table 3**.

**Figure 4:**
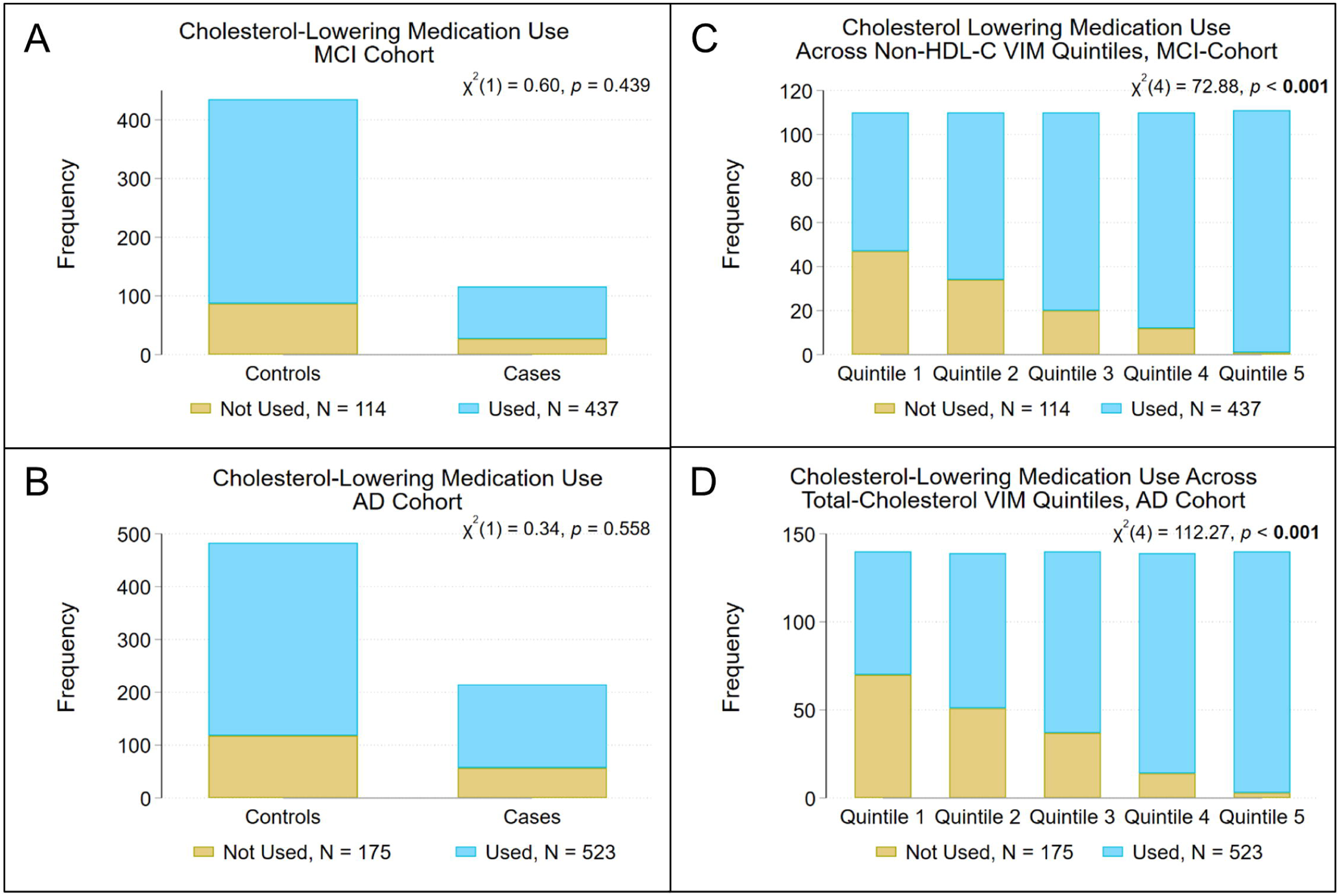
Blood-cholesterol lowering medication use in the AD and MCI case/control cohorts. Blood-cholesterol lowering medication use at age of first symptom onset (cases) did not differ from controls, at age match, in either the MCI **(A**) or AD (**B**) cohorts. Blood-cholesterol lowering medication use increased with increasing quintile of non-HDL-C variability independent of the mean (VIM) in the MCI case/control cohort (**C**) and with increasing quintile of total cholesterol VIM in the AD case/control cohort (**D**). Compare to **Table 5**.

**Table 4.** Groups identified in group-based trajectory models for lipid levels in the AD and MCI case/control cohorts.

| Lipid | Group Attribute | MCI case/control cohort, N = 551 |  |  | AD case/control cohort, N = 698 |  |  |
| --- | --- | --- | --- | --- | --- | --- | --- |
|  |  | Group 1 | Group 2 | Group 3 | Group 1 | Group 2 | Group 3 |
| Total Cholesterol | N (% of total) | 154 (27.9) | 282 (51.2) | 115 (20.9) | 319 (45.7) | 305 (43.7) | 74 (10.6) |
|  | Fit<sup>a</sup> | linear | linear | linear | linear | linear | quadratic |
|  | Average posterior probability | 0.951 | 0.938 | 0.959 | 0.968 | 0.955 | 0.951 |
|  | Odds of correct classification<sup>b</sup> | 50.5 | 14.6 | 90.0 | 35.7 | 27.1 | 133.3 |
|  | Observed probability<sup>c</sup> | 0.293 | 0.497 | 0.210 | 0.452 | 0.437 | 0.111 |
| HDL-C | N (% of total) | 306 (55.5) | 192 (34.8) | 53 (9.6) | 303 (43.4) | 288 (41.3) | 107 (15.3) |
|  | Fit<sup>a</sup> | linear | linear | quadratic | linear | quadratic | quartic |
|  | Average posterior probability | 0.991 | 0.987 | 0.985 | 0.990 | 0.982 | 0.980 |
|  | Odds of correct classification<sup>b</sup> | 84.5 | 148.1 | 627.3 | 125.6 | 79.6 | 264.3 |
|  | Observed probability<sup>c</sup> | 0.553 | 0.350 | .097 | 0.439 | 0.409 | 0.152 |
| Non-HDL-C | N (% of total) | 189 (34.3) | 284 (51.5) | 78 (14.2) | 315 (45.1) | 343 (49.1) | 40 (5.7) |
|  | Fit<sup>a</sup> | linear | linear | linear | quadratic | linear | linear |
|  | Average posterior probability | 0.941 | 0.934 | 0.958 | 0.954 | 0.943 | 0.982 |
|  | Odds of correct classification<sup>b</sup> | 30.8 | 13.2 | 137.6 | 25.1 | 17.2 | 911.1 |
|  | Observed probability<sup>c</sup> | 0.354 | 0.501 | 0.146 | 0.456 | 0.483 | 0.061 |
| LDL-C | N (% of total) | 249 (45.2) | 255 (46.3) | 47 (8.5) | 286 (41.0) | 340 (48.7) | 72 (10.3) |
|  | Fit<sup>a</sup> | linear | linear | linear | linear | linear | linear |
|  | **Average posterior probability** | 0.921 | 0.909 | 0.918 | 0.938 | 0.944 | 0.951 |
|  | **Odds of correct classification<sup>b</sup>** | 14.1 | 11.6 | 120.1 | 21.9 | 17.9 | 168.9 |
|  | **Observed probability<sup>c</sup>** | 0.446 | 0.463 | 0.091 | 0.403 | 0.491 | 0.105 |
| **ln(Triglycerides)** | **N (% of total)** | 178 (32.3) | 279 (50.6) | 94 (17.1) | 258 (37.0) | 319 (45.7) | 121 (17.3) |
|  | **Fit<sup>a</sup>** | linear | linear | linear | intercept | intercept | linear |
|  | **Average posterior probability** | 0.964 | 0.960 | 0.956 | 0.965 | 0.966 | 0.949 |
|  | **Odds of correct classification<sup>b</sup>** | 56.18 | 23.12 | 104.47 | 46.5 | 33.9 | 89.5 |
|  | **Observed probability<sup>c</sup>** | 0.328 | 0.498 | 0.173 | 0.367 | 0.464 | 0.169 |

<sup>a</sup>Modeled using a censored normal probability distribution for the dependent variable and blood-cholesterol lowering medication use as a time-dependent covariate in the decade prior to symptom onset, in cases, or matching age, in controls
<sup>b</sup>Based on the weighted posterior probability
<sup>c</sup>Group probability based on the posterior probabilities

**Table 5.** Chi-square tests of association of MCI and AD with lipid trajectory groups.

| Lipid | Trajectory Group | MCI Case/Control Group |  |  | AD Case/Control Group |  |  |
| --- | --- | --- | --- | --- | --- | --- | --- |
|  |  | Controls<sup>a</sup><br>N = 435 | Cases<sup>a</sup><br>N = 116 | *p*<sup>b</sup> | Controls<sup>a</sup><br>N = 483 | Cases<sup>a</sup><br>N = 215 | *p*<sup>b</sup> |
| Total Cholesterol | 1 | 116 (26.7) | 38 (32.8) | 0.095 | 221 (45.8) | 98 (45.6) | 0.396 |
|  | 2 | 233 (53.6) | 49 (42.2) |  | 206 (42.6) | 99 (46.0) |  |
|  | 3 | 86 (19.8) | 29 (25.0) |  | 56 (11.6) | 18 (8.4) |  |
| HDL-C | 1 | 233 (53.6) | 73 (62.9) | 0.089 | 209 (43.3) | 94 (43.7) | **0.001\*** |
|  | 2 | 155 (35.6) | 37 (31.9) |  | 185 (38.3) | 103 (47.9) |  |
|  | 3 | 47 (10.8) | 6 (5.4) |  | 89 (18.4) | 18 (8.4) |  |
| Non-HDL-C | 1 | 150 (34.5) | 39 (33.6) | 0.234 | 229 (47.4) | 86 (40.0) | 0.167 |
|  | 2 | 229 (52.6) | 55 (47.4) |  | 226 (40.0) | 117 (54.4) |  |
|  | 3 | 56 (12.9) | 22 (19.0) |  | 28 (5.8) | 12 (5.6) |  |
| LDL-C | 1 | 198 (45.5) | 51 (44.0) | 0.955 | 204 (42.2) | 82 (38.1) | 0.528 |
|  | 2 | 200 (46.0) | 55 (47.4) |  | 232 (48.0) | 108 (50.2) |  |
|  | 3 | 37 (8.5) | 10 (8.6) |  | 47 (9.7) | 25 (11.6) |  |
| ln(Triglycerides) | 1 | 150 (34.5) | 28 (24.1) | 0.050 | 178 (36.8) | 80 (37.2) | 0.632 |
|  | 2 | 209 (48.0) | 70 (60.3) |  | 217 (44.9) | 102 (47.4) |  |
|  | 3 | 76 (17.5) | 18 (15.5) |  | 88 (18.2) | 33 (15.4) |  |

<sup>a</sup>Number (percent of the total number of cases or controls)
<sup>b</sup> $\chi^2$ test. An asterisk identifies *p* values remaining significant after Bonferroni adjustment.

The number of AD cases differed across HDL-C trajectory groups (χ^2^(2)=13.14, *p*=0.001, significant following Bonferroni adjustment), but not across trajectory groups for total cholesterol, non-HDL-C, LDL-C or ln(triglycerides) (**Table 5**). HDL-C trajectory group 3, with the highest HDL-C levels, had relatively fewer cases (8.4%) than controls (18.4%), suggesting that the HDL-C group 3 trajectory is associated with a protective effect from AD. HDL-C trajectory group 2, with intermediate HDL-C levels, had relatively more cases (47.9%) than controls (38.3%). HDL-C trajectory group 1, with the lowest HDL-C levels, had similar numbers of cases (43.7%) and controls (43.3%). No significant difference in numbers of MCI cases was observed across trajectory groups for any lipid subtype (**Table 5**).

### Association of AD or MCI risk with Lipid Variability Independent of the Mean (VIM) Quintiles

The number of AD cases differed across VIM quintiles for total cholesterol (χ^2^(4)=17.29, *p*=0.002, significant after Bonferroni adjustment) and non-HDL-C (χ^2^(4)=11.34, *p*=0.023), with the greatest number of cases found in the lowest quintile (least variability) for each of these lipid types. The number of AD cases did not differ across VIM quintiles for HDL-C, LDL-C, or ln(triglycerides). For MCI, the greatest number of cases was found in the lowest VIM quintile for total cholesterol (χ^2^(4)=10.92, *p*=0.027), non-HDL-C (χ^2^(4)=13.73, *p*=0.008), and LDL-C (χ^2^(4)=10.48, *p*=0.033). The number of MCI cases did not differ across VIM quintiles for HDL-C or ln(triglycerides) (**Table 6**, **Tables S3-S6**). These results suggest that lower total cholesterol and non-HDL-C variability are associated with increased risk of AD, and that lower total cholesterol, non-HDL-C and LDL-C variability are associated with increased risk of MCI.

**Table 6.**
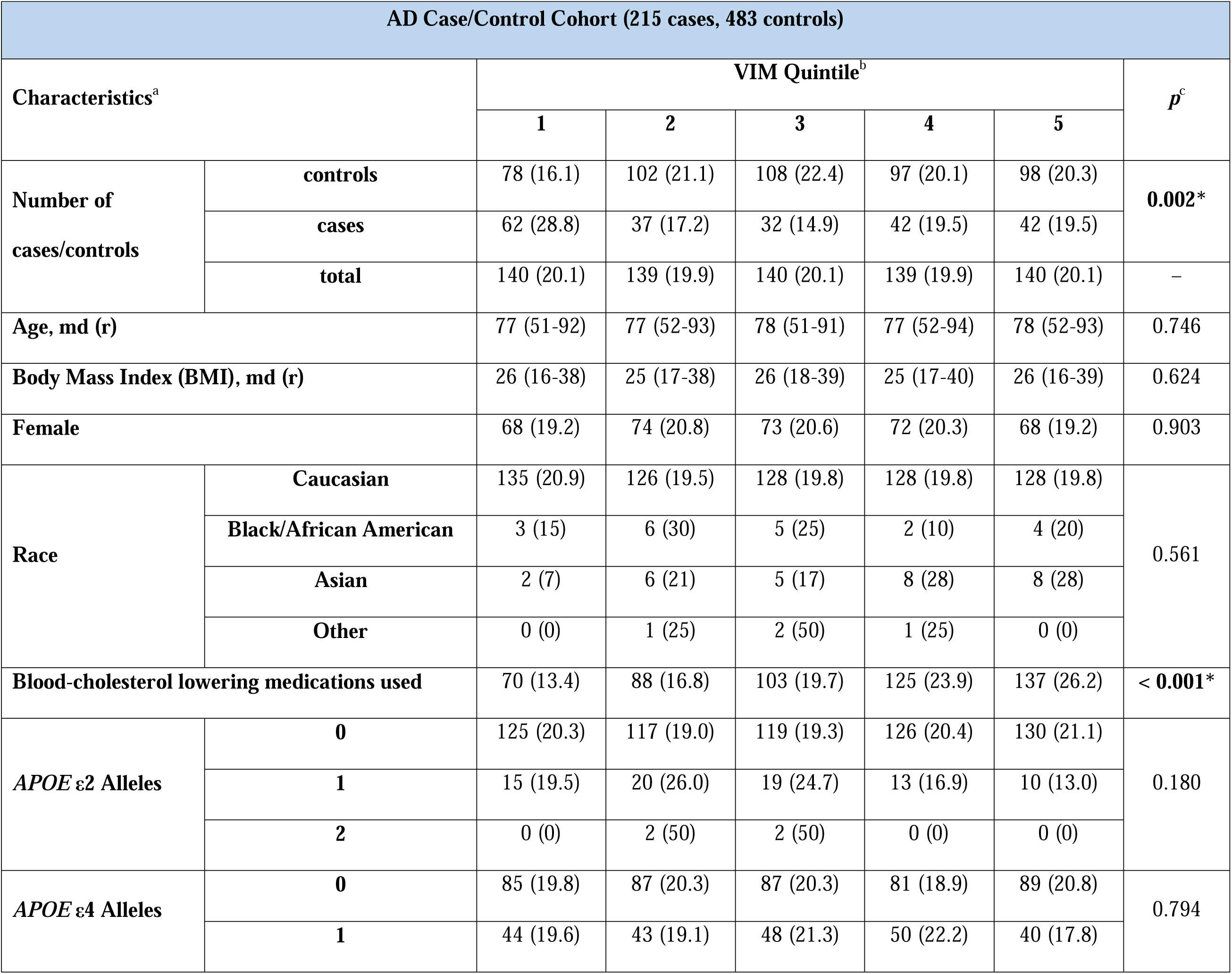
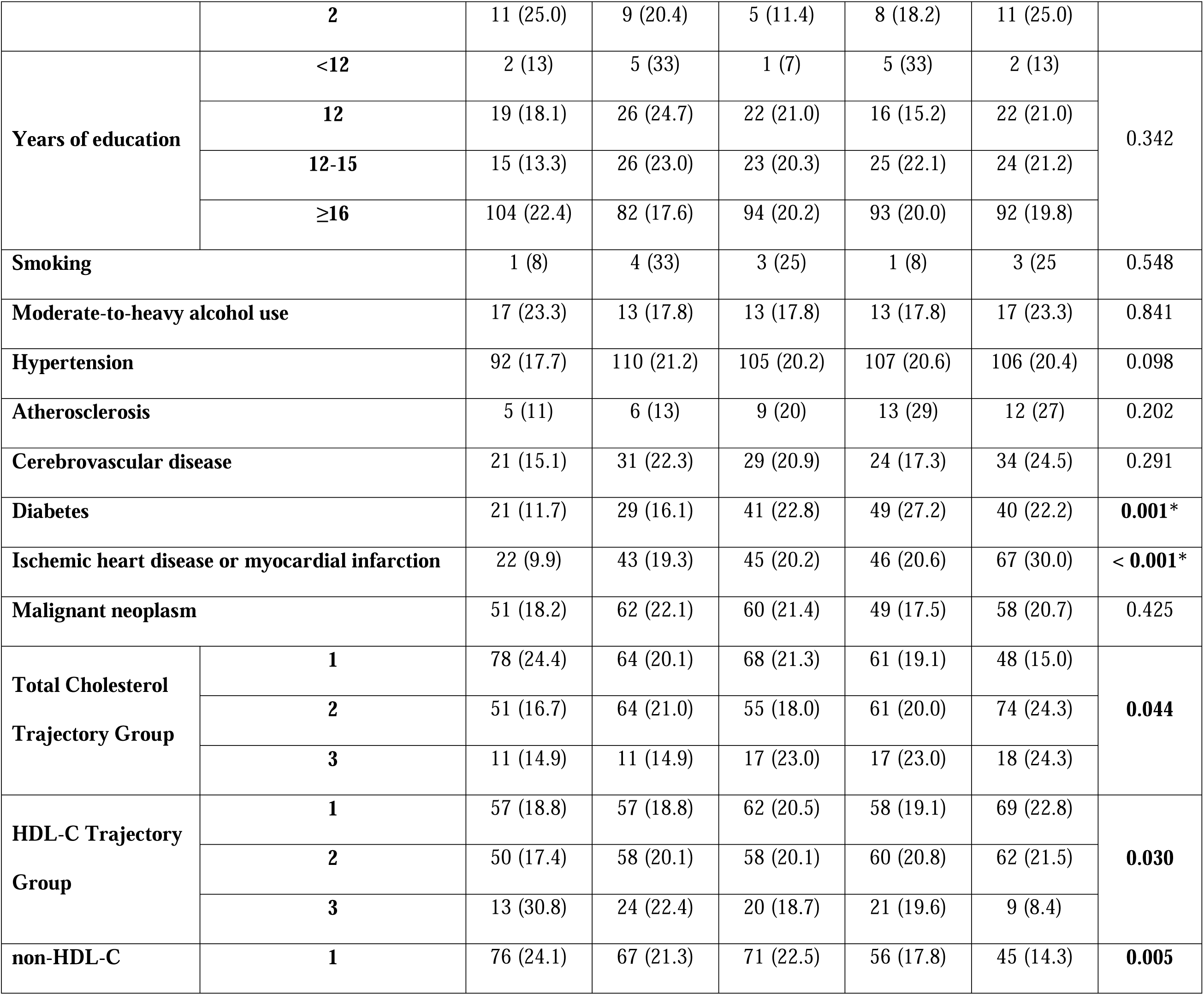
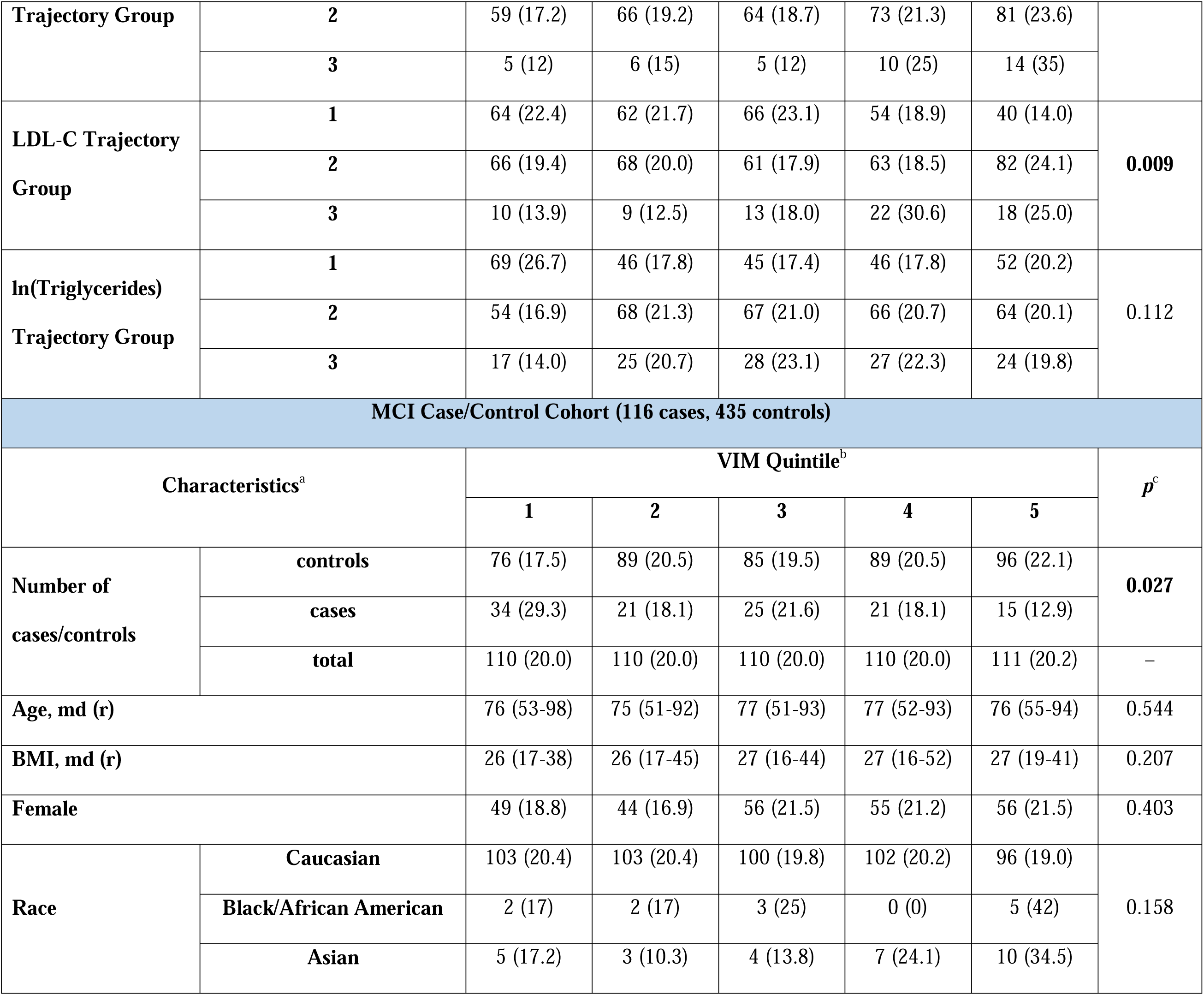
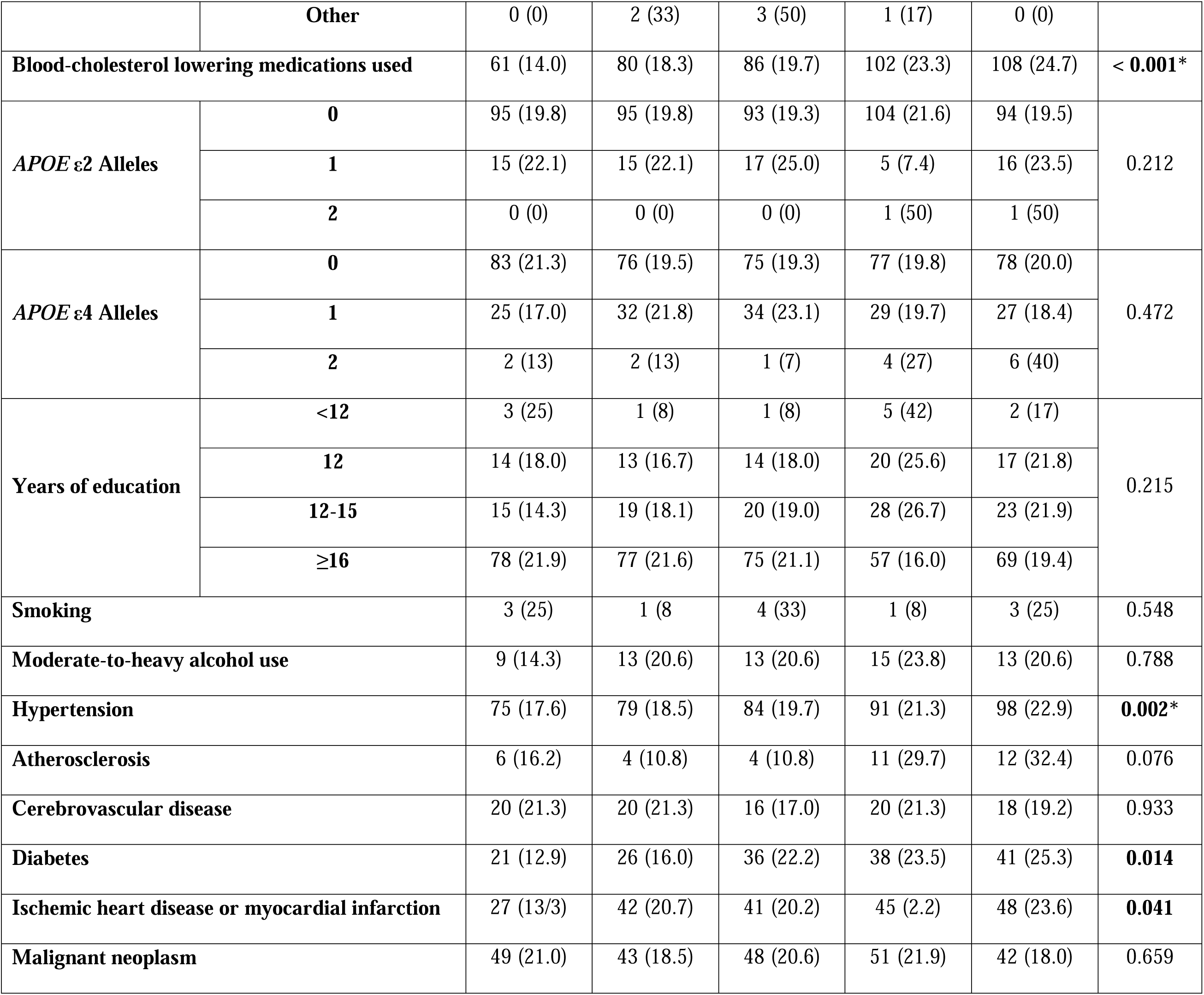
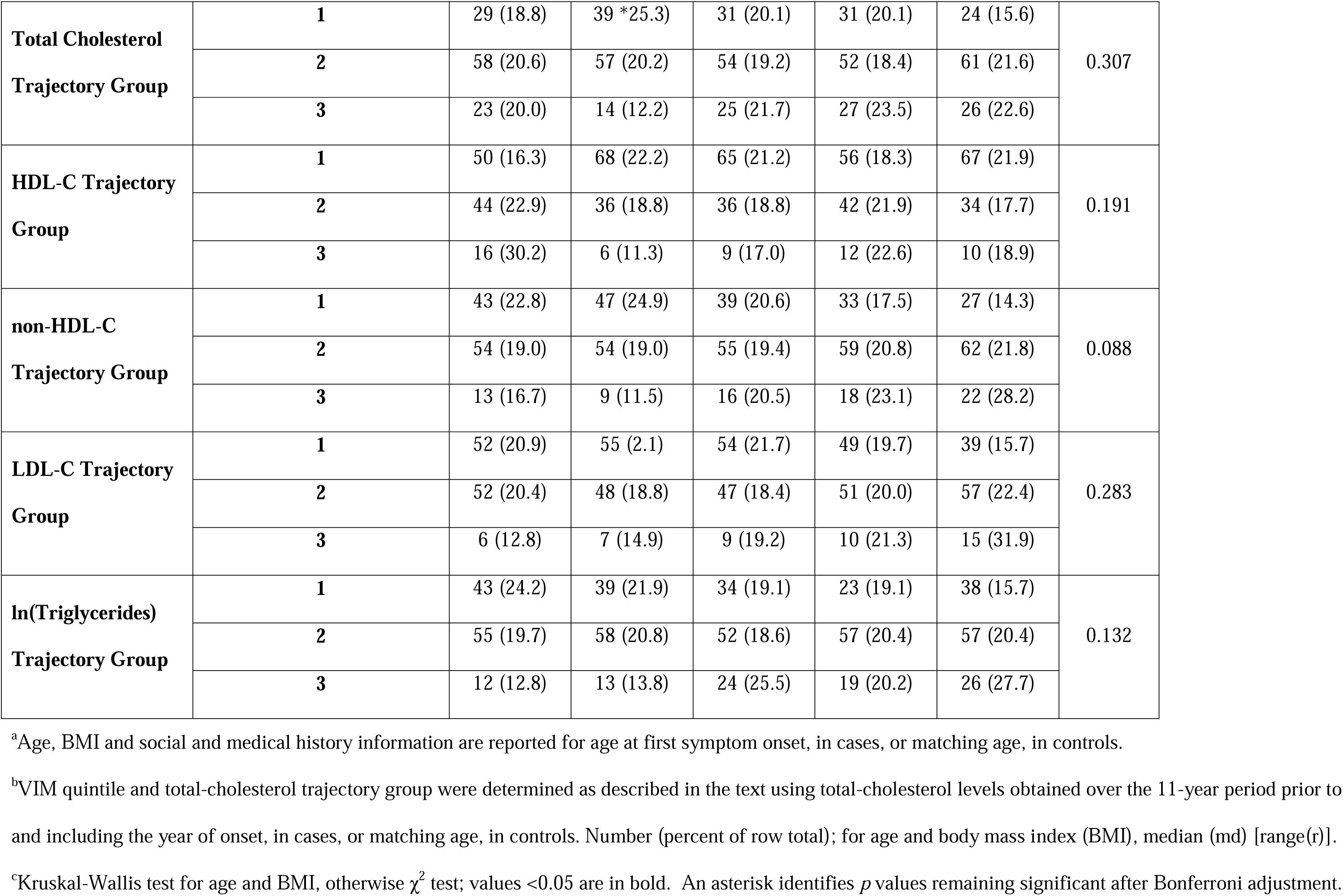
Cohort characteristics by quintile of total-cholesterol variability independent of the mean (VIM).

Consistent with Moser *et al.* (42), patient characteristics previously reported to be associated with longitudinal lipid levels (67) or risk of AD or MCI also differed across lipid VIM quintiles (**Table 6, Tables S3-S6)**. The characteristic that showed the strongest association with VIM quintile, and which remained significant after Bonferroni adjustment for multiple testing in each cohort, was use of cholesterol-lowering medication, which did not differ between cases and controls (**Figure 4**, **Table 4**).

Cholesterol-lowering medication use increased in both the MCI and AD case/control cohorts with increasing VIM quintiles for total cholesterol (MCI: χ^2^(4)=75.09, *p*<0.001; AD: χ^2^(4)=112.26, *p*<0.001), non-HDL-C (MCI: χ^2^(4)=72.88, *p*<0.001; AD: χ^2^(4)=113.10, *p*<0.001) and LDL-C (MCI: χ^2^(4)=69.11, *p*<0.001; AD: χ^2^(4)=99.37, *p*<0.001).

Several other characteristics showed associations that survived Bonferroni adjustment (**Table 5**). In the MCI cohort, an association of race with LDL-C VIM quintile (χ^2^(12)=32.70, *p*<0.001) reflected the near absence of non-Caucasians in the lowest quintile. The incidence of hypertension also increased with increasing VIM quintile of total cholesterol (χ^2^(4)=16.73, *p*=0.002), non-HDL-C (χ^2^(4)=24.76, *p*<0.001), and LDL-C (χ^2^(4)=19.13, *p*=0.001). In the AD cohort, the number of patients with diabetes increased with increasing VIM quintile for total cholesterol (χ^2^(4)=18.24, *p*=0.001), non-HDL-C (χ^2^(4)=24.13, *p*<0.001) and LDL-C (χ^2^(4)=24.82, *p*<0.001), and the number of patients with ischemic heart disease or myocardial infarction increased with increasing VIM quintile for total cholesterol (χ^2^(4)=33.42, *p*<0.001). Although similar patterns were seen in the MCI cohort, these were not significant after Bonferroni adjustment.

Associations of VIM quintile with total-cholesterol, HDL-C, non-HDL-C, and LDL-C trajectory groups, *APOE*-ε4 genotype, history of malignant neoplasm, atherosclerosis, age, or alcohol use also did not retain significance after Bonferroni adjustment in the MCI cohort.

### Models for AD and MCI Risk

We developed separate risk models for AD and MCI. **Table 7** and **Figure 5** show the results for the best fitting models.

**Figure 5:**
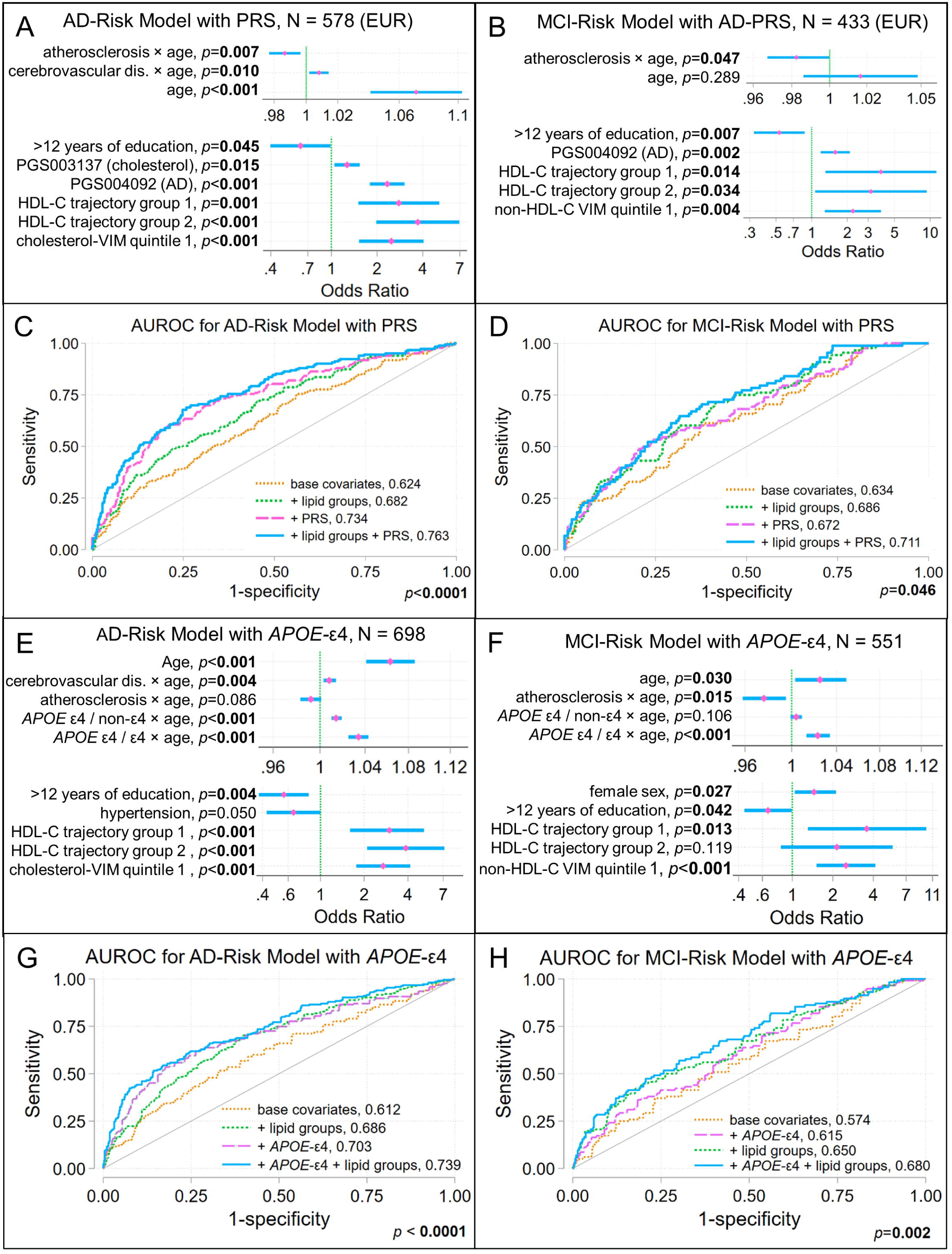
Models evaluating the contributions of lipid trajectory group and lipid variability independent of the mean (VIM) to risk of AD or MCI. Covariate-adjusted logistic regression models were developed using backwards selection and the Akaike information criterion to evaluate whether lipid trajectory groups and lipid VIM showed associations with AD or MCI risk. The covariates evaluated in these models are described in the text. Odds ratios (pink diamonds), 95% confidence intervals (blue bars), and significance are shown for covariates retained in the best fitting models for AD risk (**A**) and MCI risk (**B**) that considered polygenic risk scores for AD, including the APOE region, and lipids. Since these risk scores were mostly developed using data from genome-wide association studies in European (EUR) populations, these models were developed for the subset of study subjects with EUR ancestry. Panels **E** and **F** show odds ratios and 95% confidence intervals for covariates retained in models of AD risk (**E**) and MCI risk (**F**) that considered *APOE*-ε4 genetic status without polygenic risk scores and included all ancestries. Panels **C**, **D**, **G**, and **H** show the area under the receiver operating characteristic curve (AUROC) and scores for models with (i) only the base covariates, (ii) base covariates and polygenic risk scores or *APOE*-ε4 genotype, (iii) base covariates with lipid groups (lipid trajectory and VIM quintile groups), and (iv) base covariates, polygenic risk scores or *APOE*-ε4 genotype, and lipid groups. The Bonferroni-corrected *p* value is from a test of equality of area under these four curves. Compare panels **A**, **B**, **E**, and **F** to **Table 6**.

**Table 7.** Modeling the contribution of lipid trajectory groups and lipid VIM to AD-risk and MCI-risk.^a^.

|  | Odds Ratio, 95% Confidence Interval | *p* |
| --- | --- | --- |
| **AD-risk model including polygenic risk scores** |  |  |
| Age | 1.071, 1.041–1.103 | **<0.001** |
| Cerebrovascular disease × age | 1.008, 1.002–1.014 | **0.010** |
| Atherosclerosis × age | 0.987, 0.977–0.996 | **0.007** |
| PGS004092 (AD) | 2.333, 1.793–3.035 | **<0.001** |
| PGS003137 (total cholesterol) | 1.270, 1.048–1.539 | **0.015** |
| > 12 years of education | 0.627, 0.397–0.989 | **0.045** |
| HDL-C Trajectory Group (relative to group 3) |  |  |
| 1 | 2.781, 1.506–5.138 | **0.001** |
| 2 | 3.706, 1.974–6.954 | **<0.001** |
| Cholesterol VIM quintile 1 (relative to quintiles 2-5) | 2.480, 1.520–4.047 | **<0.001** |
| **AD-risk model including *APOE* genotypes** |  |  |
| Age | 1.063, 1.041–1.086 | **<0.001** |
| Cerebrovascular disease × age | 1.008, 1.003–1.014 | **0.004** |
| Atherosclerosis × age | 0.992, 0.983–1.001 | 0.086 |
| *APOE* ε4 genotype × age (relative to non-ε4 genotypes) |  |  |
| *APOE* ε4 heterozygote | 1.014, 1.010–1.019 | **<0.001** |
| *APOE* ε4 homozygote | 1.034, 1.025–1.043 | **<0.001** |
| > 12 years of education | 0.560, 0.377–0.831 | **0.004** |
| Hypertension | 0.653, 0.426–0.999 | 0.050 |
| HDL-C trajectory group (relative to group 3) |  |  |
| 1 | 2.983, 1.599–5.162 | **<0.001** |
| 2 | 3.868, 2.097–7.133 | **<0.001** |
| Cholesterol VIM quintile 1 | 2.704, 1.765–4.144 | **<0.001** |
| **MCI-risk model including polygenic risk scores** |  |  |
| PGS004092 (AD) | 1.580, 1.189–2.100 | **0.002** |
| Age | 1.017, 0.986, 1.048 | 0.289 |
| Atherosclerosis × age | 0.983, 0.967–0.999 | **0.047** |
| > 12 years of education | 0.532, 0.325–0.873 | **0.012** |
| HDL-C trajectory group (relative to group 3) |  |  |
| 1 | 3.841, 1.308–11.28 | **0.014** |
| 2 | 3.159, 1.068–9.342 | **0.038** |
| Non-HDL-C VIM quintile 1 | 2.228, 1.290–3.846 | **0.004** |
| **MCI-risk model including *APOE* genotypes** |  |  |
| Age | 1.025, 1.003–1.049 | **0.030** |
| Atherosclerosis × age | 0.976, 0.958–0.995 | **0.015** |
| *APOE* ε4 genotype × age (relative to non-ε4 genotypes) |  |  |
| *APOE* ε4 heterozygote | 1.004, 0.999–1.009 | 0.106 |
| *APOE* ε4 homozygote | 1.023, 1.013–1.034 | **<0.001** |
| sex | 1.451, 1.042–2.019 | **0.027** |
| college | 0.660, 0.441–0.986 | **0.042** |
| HDL-C trajectory group (relative to group 3) |  |  |
| 1 | 3.571, 1.306–9.765 | **0.013** |
| 2 | 2.138, 0.823–5.556 | 0.119 |
| Non-HDL-C VIM quintile 1 | 2.500, 1.512–4.133 | **<0.001** |

<sup>a</sup>The table presents the results of logistic regression models of AD- and MCI-risk in the AD and MCI case/control cohorts, respectively, developed as described in the text. Models with polygenic risk scores as covariates (*case ~ covariates + lipid-trajectory groups + 10 genetic PCs + polygenic risk scores*) used only individuals with EUR ancestry, as most of these scores were developed in European populations, while models with *APOE* genotypes as covariates (*case ~ covariates + lipid-trajectory groups + APOE genotypes*) used the full case/control cohorts.

One set of models included PRS (*PRS models*) previously developed using summary statistics from GWAS of AD or lipid levels; this set of models only used data from individuals with EUR ancestry. In the PRS model for AD, increased risk was associated with lower HDL-C trajectory groups, the lowest VIM quintile for total cholesterol, increasing age, cerebrovascular disease × age, higher AD PRS score, and higher total cholesterol PRS score, while atherosclerosis × age and > 12 years of education were associated with reduced risk (**Figure 5A**). In the PRS model for MCI, increased risk was associated with lower HDL-C trajectory groups, the lowest VIM quintile for non-HDL-C, and higher AD PRS score, while more than 12 years of education and atherosclerosis × age were associated with decreased risk (**Figure 5B**). The predictive performance of models incorporating only base covariates or those including base covariates and PRS improved for both AD and MCI when the lipid trajectory groups were included (**Figure 5C,D**).

A second set of models that evaluated the contribution of *APOE* status (*APOE models*) included all ancestries (**Table S2**). In the APOE model for AD, increased risk was associated with the lower HDL-C trajectory groups, lowest VIM quintile of total cholesterol, and *APOE-*ε4 × age. Having more than 12 years of education was associated with decreased risk, and atherosclerosis × age had no significant effect (**Figure 5E**). In both models, the best fit was obtained if a history of hypertension was included, although this association did not reach significance. The APOE model for MCI showed similar associations, but only the lowest HDL-C level trajectory group was associated with increased risk. An increased risk of MCI with *APOE-*ε4 × age only reached significance for *APOE-*ε4 homozygotes. Female sex was also associated with increased risk for MCI (**Figure 5F**). The predictive performance of models incorporating only base covariates or base covariates with APOE status improved for both AD and MCI when lipid trajectory groups were included (**Figure 5G,H**).

In both APOE and PRS models, the ORs and CIs associated with HDL-C trajectory groups 1 and 2 are stated relative to HDL-C trajectory group 3 (**Table 7**, **Figure 5A,B,E,F**). About 10% of the MCI case/control cohort and about 15% of the AD case/control cohort was assigned to trajectory group 3 (**Table 4**, **Figure 3B,G**). The relatively large CIs for the HRs associated with risk of MCI or AD in HDL-C trajectory groups 1 and 2, which reflect uncertainty, arise in part because of the relatively small size of HDL-C trajectory group 2 in each case/control cohort, which reflects how group-based trajectory models identified latent groups in these cohorts.

Model-predicted relationships between AD risk and HDL-C trajectory group or VIM quintile for total cholesterol relative to genetic factors were visualized by plotting predicted margins for each model. **Figure 6A–D** shows the adjusted AD risk prediction with CIs for each lipid group for patients at age 75 over the range of AD PRS (PGS004092) and total cholesterol PRS (PGS003137) scores. The density distribution of each standardized PRS in cases and controls is shown in **Figure 6E–F**. For individuals whose standardized AD PRS scores lay in the interval [-1.2, 1.5] (∼10^th^ − 93^rd^ percentiles) or whose total cholesterol standardized PRS scores lay in the interval [-1.7, 1.8] (∼4^th^ − 96^th^ percentiles), AD risk was lower in HDL-C trajectory group 3 (highest levels) than in HDL-C trajectory groups 1 and 2 (lower levels), and higher in total cholesterol VIM quintile 1 (least variability) than quintiles 2-5. For more extreme (higher or lower) PRS risk scores, the CIs of AD risk overlapped, suggesting that at very low or high levels of genetic AD risk, lipid status has no distinguishable effect.

**Figure 6:**
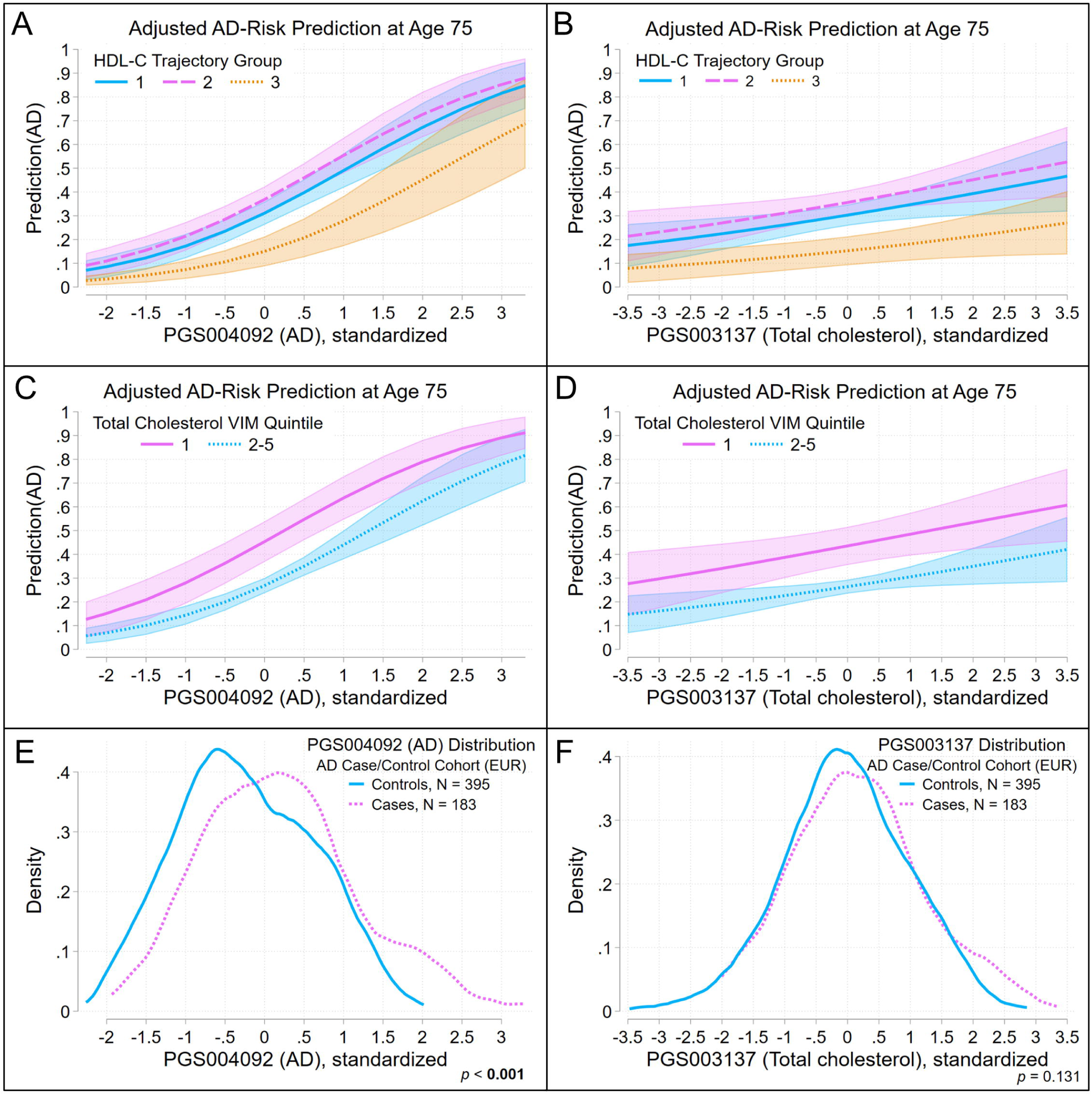
Contributions of HDL-C trajectory group and lowest VIM quintile for total cholesterol to AD risk, relative to AD and total cholesterol polygenic risk scores. The covariate-adjusted AD-risk model summarized in **Figure 5A** and **Table 6** was used to estimate contributions to AD risk relative to polygenic risk scores for AD and total cholesterol. Panel (A) shows the predictive margins for HDL-C trajectory groups and panel (**C**) for quintiles of total-cholesterol VIM over a range of values of PGS004092 (AD) at age 75 and at mean values of other covariates. Panel (**B**) shows the predictive margins for HDL-C trajectory groups and panel (**D**) for the lowest quintile of total-cholesterol VIM over a range of values of PGS003137 (total cholesterol) at age 75 and at mean values of other covariates. Lines show the predicted AD risk; shaded areas indicate 95% confidence intervals. In panels **E** and **F**, kernel-density plots obtained using an Epanechnikov kernel function illustrate the distribution of each polygenic risk score in AD cases and matched controls. A Kolmogorov-Smirnov test evaluated the equality of the distribution of each polygenic risk score in cases and controls.

Figure 7 shows the adjusted AD risk prediction with CIs for each lipid group over a range of ages for patients with no *APOE-*ε4 alleles, *APOE-*ε4 heterozygotes, and *APOE-*ε4 homozygotes. For AD models, HDL-C trajectory group 3 or total cholesterol VIM quintiles 2-5 were associated with lower risk in participants aged 64–85 with no *APOE-*ε4 alleles (Figure 7A**,B**) and in *APOE-*ε4 heterozygotes (Figure 7C**,D**), but not for *APOE-*ε4 homozygotes (Figure 7E**,F**) suggesting that the genetic risk conferred by *APOE-*ε4 in homozygotes, but not in heterozygotes, overrides risk associated with lipid levels. However, the wide, overlapping CIs for AD risk associated with lipid groups in *APOE-*ε4 homozygotes could reflect the small number of these individuals in this cohort. The distribution of *APOE-*ε4 genotypes is similar across HDL-C trajectory groups and the VIM quintiles of total cholesterol (Figure 7G**,H**).

**Figure 7:**
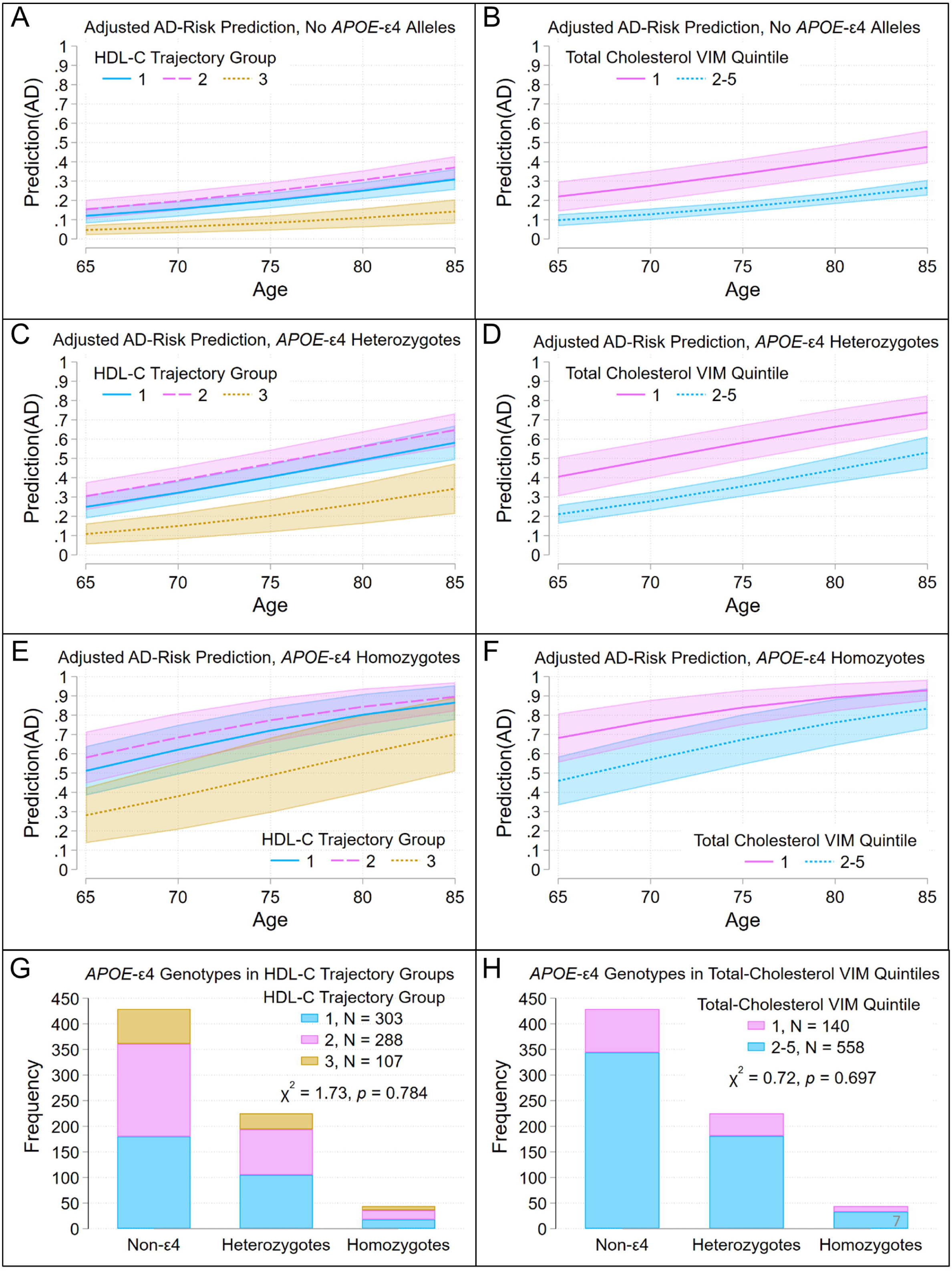
Contributions of HDL-C trajectory groups and the lowest quintile of total cholesterol VIM to AD risk, relative to *APOE*-ε4 genotype. The covariate-adjusted AD-risk model summarized in **Figure 4E** and **Table 6** was used to estimate contributions to AD risk relative to *APOE*-ε4 genotype. Panels at left show the predictive margins for HDL-C trajectory groups over ages 65 to 85 and at mean values of other covariates in non-*APOE*-ε4 genotypes (**A**), *APOE*-ε4 heterozygotes (**C**), and *APOE*-ε4 homozygotes (**E**). Panels at right show the predictive margins for quintiles of total cholesterol VIM over ages 65 to 85 and at mean values of other covariates in non-*APOE*-ε4 genotypes (**B**), *APOE*-ε4 heterozygotes (**D**), and *APOE*-ε4 homozygotes (**F**). Lines indicate predicted AD risk; shaded areas indicate 95% confidence intervals. Bar plots (bottom panels) show the distribution of HDL-C trajectory groups (**G**) and lowest total-cholesterol VIM quintile groups (**H**) across *APOE*-ε4 genotypes. A χ^2^-contingency test evaluated whether the distribution of HDL-C trajectory groups or total-cholesterol VIM quintiles differed across *APOE*-ε4 genotypes.

For MCI models, the predicted margins were also plotted (Figure 8). Here, the CIs for MCI risk in the 5^th^ and 95^th^ percentiles of AD PRS scores in patients aged 65–85 were distinct, but both overlapped with the CIs of the 50^th^ percentile (Figure 8A). Figure 8B**,C** shows the adjusted MCI risk prediction with CIs for each lipid group for patients at age 75 over the range of standardized AD PRS (PGS004092). The density distribution of the standardized PRS in cases and controls is shown in Figure 8D. While the CIs for MCI risk at age 75 in HDL-C trajectory group 2 (intermediate concentration) overlapped with those for trajectory groups 1 (highest concentration) and 3 (lowest concentration) across the range of PRS scores, these CIs were distinct for individuals in trajectory groups 1 and 3 whose scores were in the interval [-1, 1.5] (∼15^th^ − 95^th^ percentiles) (Figure 8B). Furthermore, the CIs for MCI risk in the first and second-fifth non-HDL-C quintiles at age 75 did not overlap for scores near median (∼43^rd^ to 79^th^ percentile) values (Figure 8C). This too suggests that the effect of lipid status is tempered by genetic AD risk: it has no distinguishable effect at very low or high levels of genetic AD risk.

**Figure 8:**
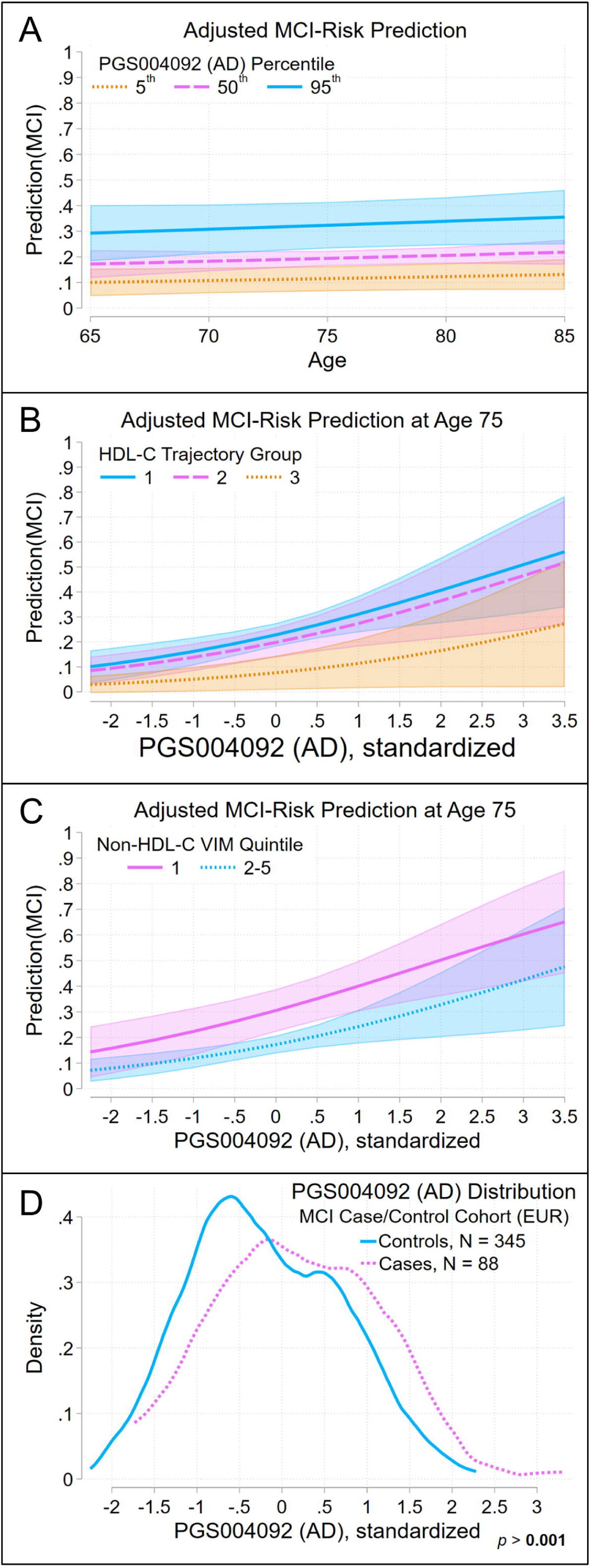
Contributions of HDL-C trajectory groups and the lowest quintile of total non-HDL-C VIM to MCI risk, relative to an AD polygenic risk score. The covariate-adjusted MCI-risk model summarized in **Figure 4B** and **Table 6** was used to estimate contributions to MCI risk relative to an AD polygenic risk score (PRS, PGS004092). (**A**) Predictive margins for 5^th^, 50^th^, and 95^th^ percentile AD PRS scores for subjects aged 65-85, at mean values of other covariates. (**B**) Predictive margins of HDL-C trajectory groups at age 75 over a range of AD PRS scores and at mean values of other covariates. (**C**) Predictive margins of the lowest quintile of total-cholesterol VIM at age 75 over a range of values of AD PRS scores and at mean values of other covariates. Lines illustrate predicted MCI risk; shaded areas indicate 95% confidence intervals. In panel **D**, kernel-density plots obtained using an Epanechnikov kernel function illustrate the distribution of the AD PRS scores in MCI cases and matched controls. A Kolmogorov-Smirnov test evaluated the equality of the distribution of the PRS in cases and controls.

The CIs for MCI risk by HDL-C trajectory group for each *APOE-*ε4 genotype in patients aged 50–85 overlapped (Figure 9A**,C****,E**). The CIs for MCI risk in non-HDL-C VIM quintile 1 and quintiles 2-5 did not overlap over the AD PRS score range [-0.35, 0.1] (∼40^th^ − 80^th^ percentiles) in non-*APOE-*ε4 genotypes over age 64 (Figure 9B), or in *APOE-*ε4 heterozygotes over age 66 (Figure 9D); however, they did overlap in *APOE-*ε4 homozygotes in patients aged 50–85 (Figure 9F), although, again, this may reflect the small number of *APOE-*ε4 homozygotes in this cohort. The distribution of *APOE-*ε4 genotypes was similar across HDL-C trajectory groups (Figure 9G), and across non-HDL-C VIM quintiles 1 and 2-5 (Figure 9H).

**Figure 9:**
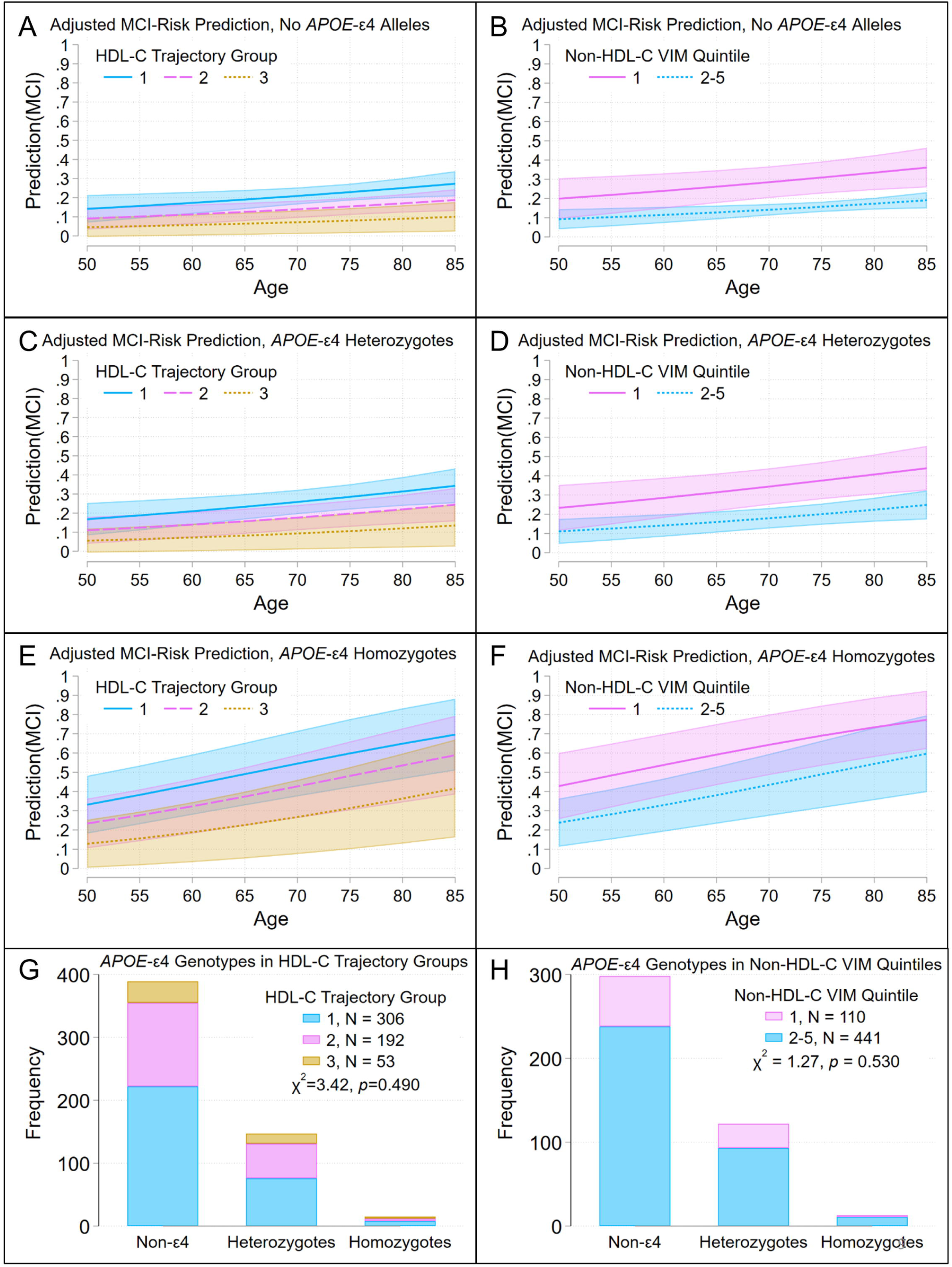
Contributions of HDL-C trajectory groups and the lowest quintile of non-HDL-C VIM to MCI risk, relative to *APOE*-ε4 genotype. The covariate adjusted MCI-risk model summarized in **Figure 5F** and **Table 6** was used to estimate contributions to MCI risk, relative to *APOE*ε4 genotype. Panels at left show the predictive margins for HDL-C trajectory groups for subjects aged 50-85 and at mean values of other covariates in (**A**) non-*APOE*-ε4 genotypes, (**C**) *APOE*-ε4 heterozygotes, and (**E**) *APOE*-ε4 homozygotes. Panels at right show the predictive margins for the lowest quintile of non-HDL-C VIM for ages 50-85 and at mean values of other covariates in (**B**) non-*APOE*-ε4 genotypes, (**D**) *APOE*-ε4 heterozygotes, and (**F**) *APOE*-ε4 homozygotes. Lines illustrate the predicted MCI risk; shaded areas indicate 95% confidence intervals. Bar plots (bottom panels) show the distribution of (**G**) HDL-C trajectory groups and (**H**) lowest total-cholesterol VIM quintile across *APOE*-ε4 genotypes. χ^2^-contingency tests evaluated whether the distribution of HDL-C trajectory groups or non-HDL-C VIM quintile groups differed across *APOE*-ε4 genotypes.

Taken together, these results indicate that the increased risk for both AD and MCI associated with lower HDL-C levels and low VIM for total cholesterol over the decade prior to symptom onset was somewhat independent of genetic and other risk factors.

## DISCUSSION

We analyzed clinically well-characterized AD and MCI case/control cohorts to address whether trajectories and VIM of blood lipid levels in the decade prior to appearance of first cognitive symptoms were associated with risk of AD and/or MCI. We used group-based trajectory models to group patients by their longitudinal lipid levels (total cholesterol, HDL-C, LDL-C, non-HDL-C and triglycerides) and by lipid VIM quintile. We then assessed whether group membership contributed to disease risk by developing risk models that included these groups, *APOE* genotype or PRS for AD and lipid levels, and known longitudinal correlates of blood-lipid levels. In these models, groups in trajectories with lower HDL-C levels or with the lowest quintile of total cholesterol VIM were significantly associated with increased risk of AD and MCI. Including these lipid-trajectory and VIM groups also improved model predictive performance.

Our results provide an important real-world perspective on aging-related cognitive outcomes that are influenced by lifestyle, environmental, cellular, and genetic factors that regulate lipid metabolism and lipid levels; genetic factors conferring risk of AD and MCI; and comorbidities associated with aging and cardiovascular health. In this patient population, increased risk of AD was also associated with older age, cerebrovascular disease × age, higher PRS_AD_, and higher PRS_total cholesterol_, while lower risk was associated with atherosclerosis × age and more than 12 years of education. Some comorbidities previously associated with AD, such as diabetes, were not retained in our risk models, while some measures of cardiovascular health, such as atherosclerosis appeared to be protective for AD. Though seemingly counterintuitive, this may suggest that medications used to treat conditions such as hypertension, atherosclerosis, or diabetes also lower the risk of Alzheimer’s disease, as suggested previously (9).

In this study, patients with the lowest quintile of total cholesterol VIM showed increased AD risk (OR[CI]: ∼2.5[1.5-4.2]), and patients with the lowest quintile of non-HDL-C VIM showed increased risk of MCI (∼2.3[1.3-4] (**Table 6**). Cholesterol and non-HDL-C VIM are strongly positively correlated (*r* = 0.93 in each cohort), so the retention of VIM quintiles of different lipids in each model reflects that which gave the best model fit. However, Moser *et al.* (42) found that the highest quintiles of total cholesterol and triglyceride VIM had an increased risk of incident AD/AD-related dementia (AD/ADRD). Several important methodological differences might underlie these different findings: (i) Our study calculated VIM based on at least three lipid measurements obtained over the ten years immediately prior to first-symptom onset in cases or matching age in controls. Moser *et al.* calculated VIM for a community population based on at least three lipid measurements over the previous five years and evaluated risk of AD/ADRD over the following 12.9 years. (ii) Our study evaluated only cases where AD and MCI were confirmed through longitudinal, multi-year follow-up, and we excluded non-AD dementia. Moser *et al*. evaluated incident AD/ADRD by ICD code. (iii) Our study evaluated risk of AD or MCI using a logistic regression model with retrospective data on lipid trajectories and VIM during the decade prior to first-symptom onset. Moser *et al*. evaluated risk of incident AD/ADRD in a large, carefully described population by analyzing survival free of AD/ADRD using Cox proportional hazards regression. Although the methodologies used in these two studies are not directly comparable, our approach has the distinct strengths of evaluating lipid levels nearer to the time of first symptom onset and using matched controls, although we evaluated data from a smaller population.

When we used our data to model disease risk, the risk associated with different lipid trajectory groups was indistinguishable at high (or low) PRS_AD_ and PRS_total cholesterol_, and in *APOE*-ε4 homozygotes (Figures 5-8). This may be partly explained by our sample size, since larger CIs were observed in *APOE*-ε4 homozygotes where estimates were based on relatively few individuals. However, under the ApoE cascade hypothesis (82), *APOE* genetic status impacts a cascade of events at the cellular and systems levels, leading to aging-related pathogenesis. *APOE*-ε4 homozygosity or higher PRS_AD_ (which includes the *APOE* region) or PRS_total cholesterol_ percentiles may be associated with impaired lipid metabolism, increased cellular stress, and dysregulation of intracellular trafficking, leading to perturbation of cellular lipid homeostasis. Individuals with these genetic factors might be expected to have AD or MCI risk that supersedes the effect of lipid levels, across different lipid groups.

These results extend our understanding of the complex set of relationships between variation in lipid levels and comorbidities associated with AD and MCI. It will be important to further clarify the relative contributions of genetic factors and blood lipid profiles to risk of AD or MCI.

### Strengths and Limitations

One of the strengths of this study is that DSM-IV criteria were used to diagnose (cases) and exclude patients with dementias that were not AD or MCI, and that patients were followed longitudinally by neurologists using SCDS toolkits designed for recording clinical data on neurological disorders at point of care. Hence, we are confident in the clinical status of patients in our study groups. Our approach also allowed us to reliably identify the year of first symptom onset, which cannot be readily inferred when ICD codes are used as a proxy for diagnosis, and which was critical to retrospectively evaluating lipid data stored in the EHR. We were also able to identify matched controls without neurodegenerative diseases. Recently, there have been efforts to develop a biological definition of AD using a combination of biomarkers that include Aβ proteinopathy, assessed from CSF or plasma Aβ assays or amyloid PET imaging, and phosphorylated and secreted AD tau, assessed from CSF or plasma assays for p-tau217, ptau181, and p-tau231 (83). Our approach complements the efforts to define a biological basis for AD. Since the above mentioned biomarkers can be used to define the biological characteristics and onset of AD, it would be valuable to address the associations between the first appearance of these biomarkers and prior lipid concentration trajectories and variation in future prospective studies.

A second strength of this study is the use of group-based trajectory models to identify groups with different longitudinal lipid trajectories. The model-fit statistics for these models provide confidence for group assignments within each case/control cohort. In the AD case/control cohort, lower mean HDL-C concentrations (∼40 mg/dL) in trajectory Group 1 were associated with increased AD risk relative to the higher mean concentrations (>80 mg/dL) in trajectory Group 3 (OR[CI] ∼3.8[2.0-7.3]; compare Figure 2B, **Table 6**).

A limitation of this study is that the time frame over which we were able to retrospectively obtain blood lipid measurements and our sample size prevented us from performing a prospective study to assess survival free of AD or MCI during aging. However, as exemplified by Duncan *et al.* (67), who assessed associations between risk of cardiovascular disease and blood lipid trajectories over a 35-year period, a study using this approach may provide considerable additional insight.

The size of our case/control cohorts (AD, N = 698; MCI, N = 551) is a second limitation of this study, and our requirement that at least 5% of each cohort be present in the smallest group constrained the number of trajectory groups. Indeed, additional lipid-level trajectory groups were identified in a study with a larger cohort (67). For this reason, we caution against extending the mean lipid values from our lipid trajectory groups to other cohorts.

### Future Directions

Several pragmatic questions arise from this work. First, whether risk of disease, more specifically genetic risk, can be reduced by modifying blood-lipid levels. Second, whether disease trajectory after diagnosis can be altered by modifying lipid levels, including by managing other comorbidities associated with high or low blood lipid levels. Addressing such questions requires detailed, clinical characterization of patients over time using point-of-care SCDS tools, such as the one used here. This study thus supports longitudinal documentation of clinical changes during the evolution of disease processes, which can then be evaluated in the context of genetic risk factors as well as aging, comorbidity, and treatment effects.

## Abbreviations

24-HC: 24-hydroxycholesterol
27-HC: 27-hydroxycholesterol
AD: Alzheimer’s disease
CI: 95% confidence interval
HDL: High-density lipoprotein
HDL-C: High density lipoprotein cholesterol
LDL: Low-density lipoprotein
LDL-C: Low-density lipoprotein cholesterol
MCI: Mild cognitive impairment
OR: Odds ratio
PRS: Polygenic risk score
VIM: Variability independent of the mean

## Data Availability

Summary statistics are available from the corresponding author upon reasonable request, following institutional approval.

## Acknowledgements

The authors thank the medical assistants, nurses, neurologists, EHR Optimization and Enterprise Data Warehouse programmers, administrators, and research personnel at Endeavor Health (formerly NorthShore University HealthSystem) who contributed to the quality improvement and practice[based research initiative using the EHR. The authors would also like to thank Alexander Epshteyn for retrieving EHR data, Anna Pham for data curation, and Ann Barlow, PhD, for assistance with writing and editing. Finally, the authors thank the neurology patients who daily inspire us to improve and innovate in our clinical practice.

## Author Contributions

Bruce Chase: Conceptualization, Methodology. Software, Validation, Formal analysis, Investigation, Data curation, Writing-original draft, Writing-review and editing. Roberta Frigerio: Conceptualization, Resources, Writing-review and editing. Chad J. Yucus: Conceptualization, Investigation, Writing-review and editing. Smita Patel: Investigation, Writing-review and editing. Demetrius Maraganore: Conceptualization, Investigation, Resources, Writing-review and editing. Alan R. Sanders: Conceptualization, Writing-review and editing. Jubao Duan: Conceptualization, Methodology, Resources, Writing-review and editing. Katerina Markopoulou: Conceptualization, Investigation, Writing-review and editing.

