## Supplemental Material for "Lipid Trajectories Improve Risk Models for Alzheimer’s Disease and Mild Cognitive Impairment"

**Item** **Page**

**Table S1: DodoNA memory-toolkit data elements**[a]

| Data Element | Recorded As |
|---|---|
| **History** | |
| Encounter date | date |
| Birth date | |
| Sex | M/F |
| Allergies/Contraindications | details |
| Tobacco Use | former/daily/some days/unknown, smokeless, passive exposure |
| Alcohol use | yes/not currently/never, drinks per week |
| Drug use | yes/not currently/never, type, per week |
| Sexual activity | Active/ever active, birth control/protection, partners |
| **Social History** | |
| Occupational history | details |
| Marital status | |
| Children | |
| Years of education | |
| Social drivers of health | alcohol use, food insecurity, stress, housing stability, interpersonal safety, financial resource strain, transportation needs, intimae partner violence, utilities, health literacy |
| **Demographics** | |
| nature of visit | initial, interval, annual |
| domicile | home, nursing home, religious community, other |
| self-repoted race/ethnicity | details |
| **Vitals** | |
| Blood pressure (systolic, diastolic) | mm Hg |
| Temperature | $^0$F |
| Pulse | Pulse rate |
| Respiration | Respiration rate |
| Weight | kg |
| Height | m |
| Body mass index (BMI) | kg/m$^2$ |
| SpO2 | percent |
| Pain score / location | details |
| **Medical History** | |
| Neurological | details |
| Sleep related | |
| Cardiovascular | |
| Autoimmune disease | |
| Behavioral | |
| Hormonal | |
| Medication | |
| B12 deficiency | |
| Folate deficiency | |
| Heart disease | |

| | |
|---|---|
| Liver failure | |
| Renal disease | |
| Normal pressure hydrocephalus | |
| Deafness | |
| Glaucoma | |
| Macular degeneration | |
| Other | |
| **Surgical history** | |
| **Family history** | |
| Allergies, arthritis, blood disease, cancer, dermatological, diabetes, endocrine, ENT, hypertension, neurological, OB/GYNE, osteoporosis, psychiatry, pulmonary/lung, smoking, stroke, thyroid disease, vision, other | Presence / absence of each in enumerated first-degree relatives |
| **Objective Test Scores** | |
| Barthel index | informant and individual question and total scores |
| FAQ | |
| Geriatric Depression Scale | |
| Short test of mental status | individual question and total scores |
| Montreal cognitive assessment test | |
| Unified Parkinson's Disease Rating Scale | |
| **Initial Symptoms** | |
| Informant | patient, spouse, sibling, offspring, friend, other |
| Year of onset | year, don't know |
| Initial symptoms | memory difficulty, word-finding difficulty, vision change, other cognitive difficulty, behavioral change, gait disorder/falls, apraxia, involuntary movements, urinary incontinence, other (specify) |
| Mode of onset | don't know, abrupt onset, insidious onset |
| Course of symptoms | don't know, slow progression, rapid progression, stepwise progression, static, improving, resolved |
| **Current Symptoms** | |
| Memory | none, recent ("short term") memory loss, repeating questions/comments, losing, misplacing items, remote memory loss, semantic memory loss, procedural memory loss |
| Language | none, word finding difficulties, name recall, reading/writing, gloval paraphasias, phenomic paraphasias |
| Visual Spatial | none, facial recognition deficit, visual spatial deficit, navigation difficulties/getting lost |
| Attention/Executive | non, disorientation, organization difficulties/problem solving deficit, distractibility/inattention, dyscalculia, judgment impaired |

| | |
|---|---|
| Gait disorder | non, shiffling, uses cane, uses walker, uses wheelchair, decreased balance, other (specify) |
| Falls | none, mechanical, orthostatic lightheadedness, assicated injuries, other (specify) |
| Type of involuntary movements | non, tremor, myoclonus, chorea, other (specify) |
| Apraxia | none, right side predominant, left side predominant, no predominance |
| Behavioral and psychological | none, depression/dysphoria, apathy/indifference, anxiety, elation/euphoria, irritability/lability, agitation/aggression, disinhibition, psychomotor disturbance (perseveration, obsessive-compulsive), hoarding, hallucinations (visual, auditory), delusions (persecution, misidentification, reduplicatoin), dleep disturbance, appetitie, other (specify) |
| Additional symptoms | non, seizures, anosima, autonomic, wandering, other (specify) |
| **Hospital visits** | |
| Emergency department visits | past year (yes/no) |
| Hospitalizations | |
| **Diagnostic studies** | CT head, MRI brain scanning, PET glucose, PET amyloid, DaTscan, EEG, neuropsychology, polysomnography, genetic testing (*APOE*, *APP*, *PS1*, *PS2*, other), CSF amyloid/tau ratio |
| **Clinical Impression** | |
| Cognitive impairment | Yes / No |
| Meets DSM-IV criteria for Mild Cognitive Impairment (MCI) | |
| Subtype of MCI | Amnestic type, single domain / Non-amnestic type, single domain / Amnestic type, multi-domain / non-amnestic type, multi-domain |
| Meets DSM-IV criteria for dementia | Yes / No |
| Alzheimer's disease | |
| Vascular dementia | |
| Lewy body dementia | |
| Frontotemporal dementia | |
| Primary progressive aphasia | |
| Other dementia / other cause | detail of diagnosis: parkinsonism, stroke, normal pressure hydrocephalus, cortical basal degeneration, progressive supranuclear palsy, stroke, alcohol use, end-stage-renal disease, subdural hematoma, B12 deficiency, psychiatric disorder, radiation therapy, etc. |
| Functional Assessment Staging Tool (FAST) | 1, 2, 3, 4, 5, 6a-e, 7a-f |

[a]Demographics, family history and AAO were collected only at study enrollment. All other data elements were collected at study enrollment and at each annual follow-up.

**Table S2. Ancestry information in AD and MCI case/control cohorts.**

| AD Case/Control Cohort | | | | | | | | | |
|---|---|---|---|---|---|---|---|---|---|
| | | | Ancestry | | | | | | |
| | | | African | Admixed American | East Asian | European | Greater Middle East | South Asian | Unknown |
| **N** | | controls | 9 (1.9) | 0 (0) | 16 (3.3) | 395 (81.8) | 12 (2.5) | 12 (2.5) | 39 (8.1) |
| | | cases | 3 (1.4) | 0 (0) | 6 (2.8) | 183 (85.1) | 3 (1.4) | 2 (0.9) | 18 (8.4) |
| | | total | 12 (1.7) | 0 (0) | 22 (3.8) | 578 (96.7) | 15 (2.5) | 14 (2.3) | 57 (9.5) |
| **Age at first lipid measurement, md (r)** | | controls | 57 (42-79) | – | 68 (50-83) | 68 (43-89) | 74 (60-85) | 68 (51-82) | 73 (47-85) |
| | | cases | 74 (59-76) | – | 66 (60-82) | 72 (46-87) | 62 (60-71) | 69 (63-75) | 77 (60-84) |
| **Match age, controls onset age, cases; md(r)** | | controls | 70 (58-86) | – | 74 (58-87) | 77 (52-94) | 81 (72-89) | 77 (61-89) | 81 (52-93) |
| | | cases | 78 (65-78) | – | 76 (66-89) | 79 (52-94) | 72 (70-75) | 75 (71-79) | 83 (67-91) |
| **Age at last follow-up/death, md (r)** | | controls | 69 (52-95) | – | 77 (65-101) | 80 (55-103) | 84 (70-99) | 79 (75-92) | 83 (60-98) |
| | | cases | 85 (70-92) | – | 83 (74-93) | 86 (59-98) | 78 (77-88) | 79 (77-81) | 90 (70-100) |
| **BMI, md (r)** | | controls | 29 (24-34) | – | 24 (16-34) | 26 (18-40) | 25 (23-30) | 25 (20-32) | 26 (18-37) |
| | | cases | 29 (26-29) | – | 26 (22-27) | 25 (16-39) | 22 (22-25) | 24 (22-26) | 26 (20-32) |
| **Female** | | controls | 5 (1) | – | 10 (2.1) | 206 (42.7) | 4 (0.8) | 3 (0.6) | 15 (3.1) |
| | | cases | 3 (1.4) | – | 3 (1.4) | 93 (43.3) | 2 (0.9) | 2 (0.9) | 9 (4.2) |
| **Blood-cholesterol lowering medications used** | | controls | 2 (0.4) | – | 15 (3.1) | 297 (61.5) | 11 (2.3) | 10 (2.1) | 30 (6.2) |
| | | cases | 1 (0.5) | – | 5 (2.3) | 133 (61.9) | 1 (0.5) | 2 (0.9) | 16 (7.4) |
| ***APOE*-ε2 alleles** | **0** | controls | 7 (1.4) | – | 15 (3.1) | 345 (71.4) | 11 (2.3) | 9 (1.9) | 32 (6.6) |
| | | cases | 3 (1.4) | – | 4 (1.9) | 168 (78.1) | 3 (1.4) | 2 (0.9) | 18 (8.4) |
| | **1** | controls | 2 (0.4) | – | 1 (0.2) | 47 (9.7) | 0 (0) | 3 (0.6) | 7 (1.4) |
| | | cases | 0 (0) | – | 2 (0.9) | 15 (7) | 0 (0) | 0 (0) | 0 (0) |
| | **2** | controls | 0 (0) | – | 0 (0) | 3 (0.6) | 1 (0.2) | 0 (0) | 0 (0) |
| | | cases | 0 (0) | – | 0 (0) | 0 (0) | 0 (0) | 0 (0) | 0 (0) |
| | **0** | controls | 4 (0.8) | – | 10 (2.1) | 264 (54.7) | 11 (2.3) | 10 (2.1) | 28 (5.8) |

| | | | | | | | | | |
|---|---|---|---|---|---|---|---|---|---|
| | | cases | 1 (0.5) | – | 5 (2.3) | 84 (39.1) | 1 (0.5) | 2 (0.9) | 9 (4.2) |
| ***APOE*-ε4 alleles** | **1** | controls | 4 (0.8) | – | 5 (1) | 116 (24) | 1 (0.2) | 2 (0.4) | 11 (2.3) |
| | | cases | 2 (0.9) | – | 0 (0) | 73 (34) | 2 (0.9) | 0 (0) | 9 (4.2) |
| | **2** | controls | 1 (0.2) | – | 1 (0.2) | 15 (3.1) | 0 (0) | 0 (0) | 0 (0) |
| | | cases | 0 (0) | – | 1 (0.5) | 26 (12.1) | 0 (0) | 0 (0) | 0 (0) |
| **Years of education** | **< 12** | controls | 0 (0) | – | 0 (0) | 14 (2.9) | 0 (0) | 0 (0) | 0 (0) |
| | | cases | 0 (0) | – | 0 (0) | 4 (1.9) | 0 (0) | 1 (0.5) | 0 (0) |
| | **12** | controls | 5 (1) | – | 4 (0.8) | 46 (9.5) | 1 (0.2) | 0 (0) | 2 (0.4) |
| | | cases | 1 (0.5) | – | 2 (0.9) | 40 (18.6) | 0 (0) | 0 (0) | 2 (0.9) |
| | **12–15** | controls | 1 (0.2) | – | 2 (0.4) | 69 (14.3) | 3 (0.6) | 1 (0.2) | 5 (1) |
| | | cases | 1 (0.5) | – | 0 (0) | 27 (12.6) | 0 (0) | 0 (0) | 4 (1.9) |
| | **≥ 16** | controls | 3 (0.6) | – | 10 (2.1) | 266 (55.1) | 8 (1.7) | 11 (2.3) | 32 (6.6) |
| | | cases | 1 (0.5) | – | 4 (1.9) | 112 (52.1) | 3 (1.4) | 1 (0.5) | 12 (5.6) |
| **Smoking** | | controls | 2 (0.4) | – | 0 (0) | 7 (1.4) | 0 (0) | 1 (0.2) | 1 (0.2) |
| | | cases | 0 (0) | – | 0 (0) | 1 (0.5) | 0 (0) | 0 (0) | 0 (0) |
| **Moderate-to-heavy alcohol use** | | controls | 0 (0) | – | 0 (0) | 51 (10.6) | 1 (0.2) | 0 (0) | 0 (0) |
| | | cases | 0 (0) | – | 0 (0) | 19 (8.8) | 0 (0) | 0 (0) | 2 (0.9) |
| **Hypertension** | | controls | 9 (1.9) | – | 15 (3.1) | 288 (59.6) | 11 (2.3) | 9 (1.9) | 32 (6.6) |
| | | cases | 3 (1.4) | – | 5 (2.3) | 130 (60.5) | 0 (0) | 2 (0.9) | 16 (7.4) |
| **Atherosclerosis** | | controls | 1 (0.2) | – | 0 (0) | 30 (6.2) | 1 (0.2) | 1 (0.2) | 2 (0.4) |
| | | cases | 0 (0) | – | 0 (0) | 7 (3.3) | 0 (0) | 1 (0.5) | 2 (0.9) |
| **Cerebrovascular disease** | | controls | 1 (0.2) | – | 2 (0.4) | 70 (14.5) | 1 (0.2) | 2 (0.4) | 7 (1.4) |
| | | cases | 1 (0.5) | – | 1 (0.5) | 48 (22.3) | 1 (0.5) | 0 (0) | 5 (2.3) |
| **Diabetes** | | controls | 1 (0.2) | – | 11 (2.3) | 89 (18.4) | 3 (0.6) | 5 (1) | 14 (2.9) |
| | | cases | 1 (0.5) | – | 3 (1.4) | 45 (20.9) | 0 (0) | 2 (0.9) | 6 (2.8) |
| **Ischemic heart disease or myocardial infarction** | | controls | 2 (0.4) | – | 5 (1) | 124 (25.7) | 9 (1.9) | 5 (1) | 17 (3.5) |
| | | cases | 0 (0) | – | 2 (0.9) | 52 (24.2) | 0 (0) | 1 (0.5) | 6 (2.8) |

| | | | | | | | | | |
|---|---|---|---|---|---|---|---|---|---|
| **Malignant neoplasm** | | controls | 4 (0.8) | – | 1 (0.2) | 182 (37.7) | 4 (0.8) | 0 (0) | 14 (2.9) |
| | | cases | 1 (0.5) | – | 1 (0.5) | 66 (30.7) | 1 (0.5) | 0 (0) | 6 (2.8) |
| **Race** | Caucasian | controls | 0 (0) | – | 1 (0.2) | 392 (81.2) | 11 (2.3) | 4 (0.8) | 34 (7) |
| | | cases | 0 (0) | – | 1 (0.5) | 183 (85.1) | 3 (1.4) | 0 (0) | 16 (7.4) |
| | Black/African American | controls | 9 (1.9) | – | 0 (0) | 0 (0) | 1 (0.2) | 0 (0) | 5 (1) |
| | | cases | 3 (1.4) | – | 0 (0) | 0 (0) | 0 (0) | 0 (0) | 2 (0.9) |
| | Asian | controls | 0 (0) | – | 15 (3.1) | 0 (0) | 0 (0) | 8 (1.7) | 0 (0) |
| | | cases | 0 (0) | – | 5 (2.3) | 0 (0) | 0 (0) | 1 (0.5) | 0 (0) |
| | Other | controls | 0 (0) | – | 0 (0) | 3 (0.6) | 0 (0) | 0 (0) | 0 (0) |
| | | cases | 0 (0) | – | 0 (0) | 0 (0) | 0 (0) | 1 (0.5) | 0 (0) |
| **MCI Case/Control Cohort** | | | | | | | | | |
| | | | **Ancestry** | | | | | | |
| | | | **African** | **Admixed American** | **East Asian** | **European** | **Greater Middle East** | **South Asian** | **Unknown** |
| **N** | | controls | 7 (1.6) | 2 (0.5) | 16 (3.7) | 345 (79.3) | 14 (3.2) | 13 (3) | 38 (8.7) |
| | | cases | 2 (1.7) | 2 (1.7) | 4 (3.4) | 88 (75.9) | 4 (3.4) | 1 (0.9) | 15 (12.9) |
| | | total | 9 (1.6) | 4 (0.7) | 20 (3.6) | 443 (78.6) | 18 (3.3) | 14 (1.2) | 53 (9.6) |
| **Age at first lipid measurement, md (r)** | | controls | 68 (67-83) | 49 (49) | 65 (58-83) | 68 (41-89) | 71 (57-85) | 65 (53-82) | 68 (47-83) |
| | | cases | 72 (67-77) | 64 (62-67) | 80 (68-88) | 71 (46-92) | 72 (62-75) | 67 | 73 (46-81) |
| **Match age, controls onset age, cases; md(r)** | | controls | 78 (75-93) | 59 (59) | 72 (68-92) | 77 (53-94) | 78 (66-89) | 74 (59-87) | 77 (56-93) |
| | | cases | 78 (75-82) | 71 (71-72) | 89 (75-93) | 77 (51-98) | 78 (68-81) | 72 | 79 (55-86) |
| **Age at last follow-up/death, md (r)** | | controls | 80 (76-95) | 58 (52-64) | 74 (67-101) | 79 (53-103) | 80 (66-99) | 79 (59-92) | 81 (64-98) |
| | | cases | 85 (82-89) | 78 (78) | 97 (81-99) | 84 (58-104) | 86 (77-89) | 85 | 86 (64-96) |
| **BMI, md (r)** | | controls | 27 (24-44) | 28 (28) | 25 (21-35) | 27 (16-52) | 26 (22-36) | 25 (21-38) | 27 (19-36) |
| | | cases | 33(28-38) | 37 (33-40) | 25 (24-26) | 27 (16-41) | 28 (22-36) | 21 | 26 (16-35) |
| **Female** | | controls | 2 (0.5) | 2 (0.5) | 9 (2.1) | 167 (38.4) | 5 (1.1) | 4 (0.9) | 13 (3) |
| | | cases | 0 (0) | 2 (1.7) | 2 (1.7) | 45 (38.8) | 2 (1.7) | 0 (0) | 7 (6) |

| | | | | | | | | | |
|---|---|---|---|---|---|---|---|---|---|
| **Blood-cholesterol lowering medications used** | | controls | 4 (0.9) | 0 (0) | 14 (3.2) | 274 (63) | 13 (3) | 12 (2.8) | 31 (7.1) |
| | | cases | 2 (1.7) | 2 (1.7) | 3 (2.6) | 64 (55.2) | 4 (3.4) | 1 (0.9) | 13 (11.2) |
| ***APOE*-ε2 alleles** | **0** | controls | 4 (0.9) | 2 (0.5) | 16 (3.7) | 306 (70.3) | 12 (2.8) | 9 (2.1) | 32 (7.4) |
| | | cases | 2 (1.7) | 2 (1.7) | 3 (2.6) | 77 (66.4) | 3 (2.6) | 0 (0) | 13 (11.2) |
| | **1** | controls | 3 (0.7) | 0 (0) | 0 (0) | 39 (9) | 2 (0.5) | 4 (0.9) | 5 (1.1) |
| | | cases | 0 (0) | 0 (0) | 1 (0.9) | 10 (8.6) | 1 (0.9) | 1 (0.9) | 2 (1.7) |
| | **2** | controls | 0 (0) | 0 (0) | 0 (0) | 0 (0) | 0 (0) | 0 (0) | 1 (0.2) |
| | | cases | 0 (0) | 0 (0) | 0 (0) | 1 (0.9) | 0 (0) | 0 (0) | 0 (0) |
| ***APOE*-ε4 alleles** | **0** | controls | 3 (0.7) | 2 (0.5) | 15 (3.4) | 246 (56.6) | 13 (3) | 11 (2.5) | 26 (6) |
| | | cases | 2 (1.7) | 1 (0.9) | 4 (3.4) | 52 (44.8) | 3 (2.6) | 1 (0.9) | 10 (8.6) |
| | **1** | controls | 4 (0.9) | 0 (0) | 1 (0.2) | 93 (21.4) | 0 (0) | 2 (0.5) | 11 (2.5) |
| | | cases | 0 (0) | 1 (0.9) | 0 (0) | 29 (25) | 1 (0.9) | 0 (0) | 5 (4.3) |
| | **2** | controls | 0 (0) | 0 (0) | 0 (0) | 6 (1.4) | 1 (0.2) | 0 (0) | 1 (0.2) |
| | | cases | 0 (0) | 0 (0) | 0 (0) | 7 (6) | 0 (0) | 0 (0) | 0 (0) |
| **Years of education** | **< 12** | controls | 0 (0) | 0 (0) | 0 (0) | 8 (1.8) | 1 (0.2) | 0 (0) | 0 (0) |
| | | cases | 1 (0.9) | 0 (0) | 0 (0) | 1 (0.9) | 0 (0) | 0 (0) | 1 (0.9) |
| | **12** | controls | 4 (0.9) | 2 (0.5) | 2 (0.5) | 42 (9.7) | 1 (0.2) | 2 (0.5) | 3 (0.7) |
| | | cases | 0 (0) | 0 (0) | 1 (0.9) | 19 (16.4) | 1 (0.9) | 0 (0) | 1 (0.9) |
| | **12–15** | controls | 2 (0.5) | 0 (0) | 4 (0.9) | 67 (15.4) | 3 (0.7) | 3 (0.7) | 6 (1.4) |
| | | cases | 1 (0.9) | 1 (0.9) | 0 (0) | 16 (13.8) | 1 (0.9) | 0 (0) | 1 (0.9) |
| | **≥ 16** | controls | 1 (0.2) | 0 (0) | 10 (2.3) | 228 (52.4) | 9 (2.1) | 8 (1.8) | 29 (6.7) |
| | | cases | 0 (0) | 1 (0.9) | 3 (2.6) | 52 (44.8) | 2 (1.7) | 1 (0.9) | 12 (10.3) |
| **Smoking** | | controls | 2 (0.5) | 0 (0) | 0 (0) | 6 (1.4) | 0 (0) | 0 (0) | 1 (0.2) |
| | | cases | 0 (0) | 0 (0) | 0 (0) | 3 (2.6) | 0 (0) | 0 (0) | 0 (0) |
| **Moderate-to-heavy alcohol use** | | controls | 0 (0) | 0 (0) | 0 (0) | 48 (11) | 1 (0.2) | 0 (0) | 1 (0.2) |
| | | cases | 0 (0) | 0 (0) | 0 (0) | 11 (9.5) | 2 (1.7) | 0 (0) | 0 (0) |
| **Hypertension** | | controls | 7 (1.6) | 1 (0.2) | 14 (3.2) | 256 (58.9) | 13 (3) | 11 (2.5) | 32 (7.4) |

| | | | | | | | | | |
|---|---|---|---|---|---|---|---|---|---|
| | | cases | 2 (1.7) | 2 (1.7) | 4 (3.4) | 70 (60.3) | 3 (2.6) | 1 (0.9) | 11 (9.5) |
| **Atherosclerosis** | | controls | 2 (0.5) | 0 (0) | 0 (0) | 28 (6.4) | 3 (0.7) | 1 (0.2) | 1 (0.2) |
| | | cases | 0 (0) | 0 (0) | 0 (0) | 2 (1.7) | 0 (0) | 0 (0) | 0 (0) |
| **Cerebrovascular disease** | | controls | 0 (0) | 0 (0) | 3 (0.7) | 58 (13.3) | 2 (0.5) | 1 (0.2) | 7 (1.6) |
| | | cases | 0 (0) | 0 (0) | 1 (0.9) | 18 (15.5) | 1 (0.9) | 0 (0) | 3 (2.6) |
| **Diabetes** | | controls | 3 (0.7) | 1 (0.2) | 10 (2.3) | 95 (21.8) | 4 (0.9) | 3 (0.7) | 10 (2.3) |
| | | cases | 2 (1.7) | 1 (0.9) | 2 (1.7) | 24 (20.7) | 1 (0.9) | 1 (0.9) | 5 (4.3) |
| **Ischemic heart disease or myocardial infarction** | | controls | 2 (0.5) | 0 (0) | 3 (0.7) | 128 (29.4) | 8 (1.8) | 5 (1.1) | 19 (4.4) |
| | | cases | 0 (0) | 0 (0) | 0 (0) | 28 (24.1) | 3 (2.6) | 0 (0) | 7 (6) |
| **Malignant neoplasm** | | controls | 5 (1.1) | 0 (0) | 3 (0.7) | 162 (37.2) | 4 (0.9) | 2 (0.5) | 12 (2.8) |
| | | cases | 2 (1.7) | 1 (0.9) | 1 (0.9) | 33 (28.4) | 1 (0.9) | 0 (0) | 7 (6) |
| **Race** | Caucasian | controls | 0 (0) | 0 (0) | 1 (0.2) | 344 (79.1) | 13 (3) | 3 (0.7) | 26 (6) |
| | | cases | 0 (0) | 0 (0) | 0 (0) | 88 (75.9) | 4 (3.4) | 0 (0) | 15 (12.9) |
| | Black/African American | controls | 7 (1.6) | 0 (0) | 0 (0) | 0 (0) | 1 (0.2) | 1 (0.2) | 1 (0.2) |
| | | cases | 2 (1.7) | 0 (0) | 0 (0) | 0 (0) | 0 (0) | 0 (0) | 0 (0) |
| | Asian | controls | 0 (0) | 0 (0) | 15 (3.4) | 0 (0) | 0 (0) | 9 (2.1) | 0 (0) |
| | | cases | 0 (0) | 0 (0) | 4 (3.4) | 0 (0) | 0 (0) | 1 (0.9) | 0 (0) |
| | Other | controls | 0 (0) | 2 (0.5) | 0 (0) | 1 (0.2) | 0 (0) | 0 (0) | 1 (0.2) |
| | | cases | 0 (0) | 2 (1.7) | 0 (0) | 0 (0) | 0 (0) | 0 (0) | 0 (0) |

[a]Unless otherwise specified, the table reports patient information at the age of first-symptom onset in cases and at the age of match in controls. Entries are N (percent of the total number of cases or percent the total number of controls). Ancestry was determined during genotype imputation using the TOPMED imputation server. Abbreviations: md, median; r, range; body mass index, BMI.

**Table S3. Cohort characteristics by quintile of non-HDL-C variability independent of the mean (VIM).**

| AD Case/Control Cohort (215 cases, 483 controls) | | | | | | | |
|---|---|---|---|---|---|---|---|
| **Characteristics**[a] | | **VIM Quintile**[b] | | | | | ***p***[c] |
| | | **1** | **2** | **3** | **4** | **5** | |
| **Number of cases/controls** | **controls** | 81 (16.8) | 99 (20.5) | 105 (21.7) | 99 (20.5) | 99 (20.5) | **0.023** |
| | **cases** | 59 (27.4) | 40 (18.6) | 35 (16.3) | 40 (18.5) | 41 (19.1) | |
| | **total** | 140 (20.1) | 139 (19.9) | 140 (20.1) | 139 (19.9) | 140 (20.1) | – |
| **Age, md (r)** | | 76 (51-92) | 77 (52-93) | 78 (52-92) | 78 (52-94) | 78 (52-93) | 0.215 |
| **Body Mass Index (BMI), md (r)** | | 26 (16-38) | 25 (16-39) | 26 (18-39) | 25 (17-40) | 26 (16-39) | 0.700 |
| **Female** | | 76 (21.4) | 72 (20.3) | 65 (18.3) | 67 (18.9) | 75 (21.1) | 0.625 |
| **Race** | **Caucasian** | 130 (20.2) | 129 (20.0) | 130 (20.2) | 128 (19.8) | 128 (19.8) | 0.374 |
| | **Black/African American** | 8 (40) | 3 (15) | 2 (10) | 3 (15) | 4 (20) | |
| | **Asian** | 2 (7) | 5 (17) | 7 (24) | 7 (24) | 8 (28) | |
| | **Other** | 0 (0) | 2 (50) | 1 (25) | 1 (25) | 0 (0) | |
| **Blood-cholesterol lowering medications used** | | 67 (12.8) | 87 (16.6) | 114 (21.8) | 120 (22.9) | 135 (25.8) | **< 0.001*** |
| ***APOE* ε2 Alleles** | **0** | 129 (20.9) | 119 (19.3) | 117 (19.0) | 122 (19.8) | 150 (21.1) | 0.223 |
| | **1** | 11 (14.3) | 19 (24.7) | 21 (27.3) | 16 (20.8) | 10 (13.0) | |
| | **2** | 0 (0) | 1 (25) | 2 (50) | 1 (25) | 0 (0) | |
| ***APOE* ε4 Alleles** | **0** | 81 (18.9) | 91 (21.2) | 85 (19.8) | 83 (19.4) | 89 (20.8) | 0.457 |
| | **1** | 44 (19.6) | 42 (18.7) | 48 (21.3) | 49 (21.8) | 42 (18.7) | |
| | **2** | 15 (34.1) | 6 913.6) | 7 (15.9) | 7 (15.9) | 9 (20.4) | |
| | **<12** | 4 (27) | 2 (13) | 3 (20) | 2 (13) | 4 (27) | 0.837 |

| | | | | | | | |
|---|---|---|---|---|---|---|---|
| **Years of education** | **12** | 24 (22.9) | 22 (21.0) | 17 (16.2) | 19 (18.1) | 23 (21.9) | |
| | **12-15** | 17 (15.0) | 19 (16.8) | 28 (24.8) | 27 (23.9) | 22 (19.5) | |
| | **≥16** | 95 (20.4) | 96 (20.6) | 92 (19.8) | 91 (19.6) | 91 (19.6) | |
| **Smoking** | | 3 (25) | 2 (17) | 4 (33) | 0 (0) | 3 (25) | 0.424 |
| **Moderate-to-heavy alcohol use** | | 16 (21.9) | 10 (13.7) | 15 (20.6) | 16 (21.9) | 16 (21.9) | 0.729 |
| **Hypertension** | | 98 (18.8) | 91 (17.5) | 114 (21.9) | 108 (20.8) | 109 (21.0) | **0.014** |
| **Atherosclerosis** | | 6 (13.3) | 5 (11.1) | 9 (20.0) | 13 (28.9) | 12 (26.7) | 0.203 |
| **Cerebrovascular disease** | | 24 (17.3) | 27 (19.4) | 33 (23.7) | 22 (15.8) | 33 (23.7) | 0.343 |
| **Diabetes** | | 19 (10.6) | 27 (15.0) | 48 (26.7) | 47 (26.1) | 39 (21.7) | < **0.001*** |
| **Ischemic heart disease or myocardial infarction** | | 22 (9.9) | 33 (14.8_ | 55 (24.7) | 43 (19.3) | 70 (31.4) | < **0.001*** |
| **Malignant neoplasm** | | 60 (21.4) | 54 (19.3) | 59 (21.1) | 50 (17.9) | 57 (20.4) | 0.775 |
| **Total Cholesterol Trajectory Group** | **1** | 68 (21.3) | 67 (21.0) | 74 (23.2) | 65 (20.4) | 45 (14.1) | **0.029** |
| | **2** | 58 (19.0) | 61 (20.0) | 49 (16.1) | 58 (19.0) | 79 (25.9) | |
| | **3** | 14 (18.9) | 11 (14.9) | 17 (23.0) | 16 (21.6) | 16 (21.6) | |
| **HDL-C Trajectory Group** | **1** | 58 (19.1) | 58 (19.1) | 65 (21.4) | 59 (19.5) | 63 (20.8) | 0.543 |
| | **2** | 53 (18.4) | 60 (20.8) | 52 (18.1) | 62 (21.5) | 61 (21.2) | |
| | **3** | 29 (27.1) | 21 (19.6) | 23 (21.5) | 18 (16.8) | 16 (15.0) | |
| **non-HDL-C Trajectory Group** | **1** | 67 (21.3) | 66 (21.0) | 71 (22.5) | 58 (18.4) | 54 (16.8) | 0.378 |
| | **2** | 67 (19.5) | 67 (19.5) | 63 (18.4) | 71 (20.7) | 75 (21.9) | |
| | **3** | 6 (15.0) | 6 915.0) | 6 (15.0) | 10 (25.0) | 12 (30.0) | |
| **LDL-C Trajectory Group** | **1** | 58 (20.3) | 59 (20.6) | 70 (24.5) | 54 (18.9) | 45 (15.7) | 0.071 |
| | **2** | 71 (20.9) | 67 (19.7) | 57 (16.8) | 64 (18.8) | 81 (23.8) | |

| | | | | | | | |
|---|---|---|---|---|---|---|---|
| | **3** | 11 (15.3) | 13 (18.1) | 13 (18.1) | 21 (29.2) | 14 (19.4) | |
| **ln(Triglycerides) Trajectory Group** | **1** | 57 (22.1) | 55 (21.3) | 49 (19.0) | 41 (15.9) | 56 (21.7) | 0.657 |
| | **2** | 61 (19.1) | 59 (18.5) | 65 (20.4) | 71 (22.3) | 63 (19.8) | |
| | **3** | 22 (18.2) | 25 (20.7) | 26 (21.5) | 27 (22.3) | 21 (17.4) | |
| **MCI Case/Control Cohort (116 cases, 435 controls)** | | | | | | | |
| **Characteristics**[a] | | **VIM Quintile**[b] | | | | | ***p***[c] |
| | | **1** | **2** | **3** | **4** | **5** | |
| **Number of cases/controls** | **controls** | 74 (17.0) | 85 (19.5) | 93 (21.4) | 89 (20.5) | 94 (21.6) | **0.008** |
| | **cases** | 36 (31.0) | 25 (21.6) | 17 (14.7) | 21 (18.1) | 17 (14.7) | |
| | **total** | 110 (20.0) | 110 (20.0) | 110 (20.0) | 110 (20.0) | 111 (20.2) | – |
| **Age, md (r)** | | 76 (51-91) | 75 (51-98) | 76 (52-89) | 78 (53-94) | 76 (54-94) | **0.019** |
| **BMI, md (r)** | | 26 (17-40) | 26 (17-46) | 27 (19-52) | 26 (16-43) | 27 (16-41) | 0.462 |
| **Female** | | 50 (19.2) | 53 920.4) | 42 (16.20 | 57 (21.9) | 58 (22.3) | 0.211 |
| **Race** | **Caucasian** | 105 920.8) | 103 (20.4) | 98 (19.4) | 101 (20.0) | 97 (19.2) | **0.028** |
| | **Black/African American** | 2 (17) | 0 (0) | 5 942) | 1 (8) | 4 (33) | |
| | **Asian** | 3 (10) | 4 (14) | 4 (14) | 8 (28) | 10 (34) | |
| | **Other** | 0 (0) | 3 (50) | 3 (50) | 0 (0) | 0 (0) | |
| **Blood-cholesterol lowering medications used** | | 63 (14.4) | 76 (17.4) | 90 (20.6) | 98 (22.4) | 110 (25.2) | **< 0.001*** |
| ***APOE* ε2 Alleles** | **0** | 99 (20.6) | 98 (20.4) | 92 (19.1) | 96 (20.0) | 96 (20.0) | 0.773 |
| | **1** | 11 (16.2) | 12 (17.6) | 17 (25.0) | 14 (20.6) | 14 (20.6) | |
| | **2** | 0 (0) | 0 (0) | 1 (50) | 0 (0) | 1 (50) | |
| ***APOE* ε4 Alleles** | **0** | 73 (18.8) | 92 (21.1) | 80 (20.6) | 76 (19.5) | 78 (20.0) | 0.756 |
| | **1** | 34 (23.1) | 27 (18.4) | 28 (19.0) | 30 (20.4) | 28 (19.0) | |

| | | | | | | | |
|---|---|---|---|---|---|---|---|
| | **2** | 3 (20) | 1 (7) | 2 (13) | 4 (27) | 5 (33) | |
| **Years of education** | **<12** | 3 (25) | 0 (0) | 4 (33) | 2 (17) | 3 (25) | 0.654 |
| | **12** | 15 (19.2) | 14 (18.0) | 13 (16.7) | 17 (21.8) | 19 (24.4) | |
| | **12-15** | 16 (15.2) | 19 (18.1) | 22 (21.0) | 26 (24.8) | 22 (21.0) | |
| | **≥16** | 76 (21.4) | 77 (21.6) | 71 (19.9) | 65 (18.3) | 67 (18.8) | |
| **Smoking** | | 3 (25) | 1 (8) | 3 (25) | 2 (17) | 3 (25) | 0.852 |
| **Moderate-to-heavy alcohol use** | | 10 (15.9) | 11 (17.5) | 14 (22.2) | 12 (19.0) | 16 (25.4) | 0.735 |
| **Hypertension** | | 73 (17.1) | 75 (17.6) | 92 (21.6) | 88 (20.6) | 99 (23.2) | < **0.001*** |
| **Atherosclerosis** | | 7 (19) | 4 (11) | 4 (11) | 8 (22) | 14 (38) | **0.048** |
| **Cerebrovascular disease** | | 18 (19.2) | 17 (18.1) | 15 (16.0) | 25 (26.6) | 19 (20.2) | 0.456 |
| **Diabetes** | | 20 (12.4) | 25 (15.3) | 39 (24.1) | 39 (24.1) | 29 (24.1) | **0.005** |
| **Ischemic heart disease or myocardial infarction** | | 28 (13.8) | 36 (17.7) | 45 (22.2) | 45 (22.2) | 49 (24.1) | **0.026** |
| **Malignant neoplasm** | | 49 (21.0) | 46 (19.7 | 49 (21.0) | 47 (20.2) | 42 (18.0) | 0.848 |
| **Total Cholesterol Trajectory Group** | **1** | 31 (20.1) | 31 (20.1) | 36 (23.4) | 26 (16.9) | 30 (19.5) | 0.803 |
| | **2** | 59 (20.9) | 59 (20.9) | 49 (17.4) | 57 (20.2) | 58 (20.6) | |
| | **3** | 20 (17.4) | 20 (17.4) | 25 (21.7) | 27 (23.5) | 23 (20.0) | |
| **HDL-C Trajectory Group** | **1** | 54 (17.7) | 68 (22.2) | 65 (21.2) | 56 (18.3) | 63 (20.6) | 0.262 |
| | **2** | 44 (22.9) | 35 (18.2) | 38 (19.8) | 37 (19.3) | 38 (19.8) | |
| | **3** | 12 (22.6) | 7 (13.2) | 7 (13.2) | 17 (32.1) | 10 (18.9) | |
| **non-HDL-C Trajectory Group** | **1** | 43 (22.8) | 24 (18.5) | 43 (22.8) | 31 (16.4) | 37 (19.6) | 0.457 |
| | **2** | 54 (19.0) | 61 (21.5) | 48 (16.9) | 65 (22.9) | 56 (19.7) | |
| | **3** | 13 (16.7) | 14 (18.0) | 19 (24.4) | 14 (18.0) | 18 (23.1) | |

| | | | | | | | |
|---|---|---|---|---|---|---|---|
| **LDL-C Trajectory Group** | **1** | 49 (19.7) | 47 (18.9) | 58 (23.3) | 47 (18.9) | 48 (19.3) | 0.634 |
| | **2** | 55 921.6) | 53 (20.8) | 41 (16.1) | 54 (21.2) | 52 (20.4) | |
| | **3** | 6 (12.8) | 10 (21.3) | 11 (23.4) | 9 (19.2) | 11 (23.4) | |
| **ln(Triglycerides) Trajectory Group** | **1** | 40 (22.5) | 35 (19.7) | 35 (19.7) | 35 (19.7) | 33 (18.5) | 0.632 |
| | **2** | 59 (21.2) | 57 (20.4) | 55 (19.7) | 54 (19.4) | 54 (19.4) | |
| | **3** | 11 (11.7) | 18 (19.2) | 20 (21.3) | 21 (22.3) | 24 (25.5) | |

[a]Age, BMI and social and medical history information are reported for age at first symptom onset, in cases, or matching age, in controls.

[b]VIM quintile and total-cholesterol trajectory group were determined as described in the text using total-cholesterol levels obtained over the 11-year period prior to and including the year of onset, in cases, or matching age, in controls. Number (percent of row total); for age and body mass index (BMI), median (md) [range(r)].

[c]Kruskal-Wallis test for age and BMI, otherwise $\chi^2$ test; values <0.05 are in bold. An asterisk identifies *p* values remaining significant after Bonferroni adjustment.

**Table S4. Cohort characteristics by quintile of HDL-C variability independent of the mean (VIM).**

| AD Case/Control Cohort (215 cases, 483 controls) | | | | | | | |
|---|---|---|---|---|---|---|---|
| Characteristics[a] | | VIM Quintile[b] | | | | | *p*[c] |
| | | 1 | 2 | 3 | 4 | 5 | |
| Number of cases/controls | controls | 95 (19.7) | 98 (20.3) | 95 (19.7) | 95 (19.7) | 100 (20.7) | 0.949 |
| | cases | 45 (20.9) | 41 (19.1) | 45 (20.9) | 44 (20.5) | 40 (18.6) | |
| | total | 140 (20.1) | 139 (19.9) | 140 (20.1) | 139 (19.9) | 140 (20.1) | – |
| Age, md (r) | | 79 (52-90) | 76 (52-93) | 78 (51-94) | 77 (52-89) | 77 (51-93) | 0.555 |
| Body Mass Index (BMI), md (r) | | 26 (17-36) | 26 (16-39) | 25 (16-40) | 25 (18-39) | 26 (18-37) | 0.373 |
| Female | | 76 (21.4) | 62 (17.5) | 73 (20.6) | 72 (20.3) | 72 (20.3) | 0.559 |
| Race | Caucasian | 132 (20.5) | 127 (19.7) | 128 (19.8) | 128 (19.8) | 130 (20.2) | **0.032** |
| | Black/African American | 2 (10) | 2 (10) | 3 915) | 5 (25) | 8 (40) | |
| | Asian | 6 (21) | 9 (31) | 9 (31) | 3 (10) | 2 (7) | |
| | Other | 0 (0) | 1 (25) | 0 (0) | 3 (75) | 0 (0) | |
| Blood-cholesterol lowering medications used | | 104 (19.9) | 104 (19.9) | 102 (19.5) | 108 (20.6) | 105 (20.1) | 0.922 |
| *APOE* ε2 Alleles | 0 | 127 (20.6) | 124 (20.1) | 119 (19.3) | 123 (19.9) | 124 (20.1) | 0.153 |
| | 1 | 13 (16.9) | 12 (15.6) | 21 (27.3) | 15 (19.5) | 16 (20.8) | |
| | 2 | 0 (0) | 3 (75) | 0 (0) | 1 (25) | 0 (0) | |
| *APOE* ε4 Alleles | 0 | 86 (20.0) | 88 (20.5) | 88 (20.5) | 85 (19.8) | 82 (19.1) | 0.477 |
| | 1 | 49 (21.8) | 46 (20.4) | 39 (17.3) | 44 (19.6) | 47 (20.9) | |
| | 2 | 5 (11.4) | 5 (11.4) | 13 (29.6) | 10 (22.7) | 11 (25.0) | |
| | <12 | 3 (20) | 4 (27) | 3 (20) | 2 (13) | 3 (20) | 0.855 |

| | | | | | | | |
|---|---|---|---|---|---|---|---|
| **Years of education** | **12** | 23 (21.9) | 19 (18.1) | 21 (20.0) | 26 (24.8) | 16 (15.2) | |
| | **12-15** | 23 (20.4) | 28 (24.8) | 19 (16.8) | 23 (20.4) | 20 (17.7) | |
| | **≥16** | 91 (19.6) | 88 (18.9) | 97 (20.9) | 88 (18.9) | 101 (21.7) | |
| **Smoking** | | 3 (25) | 3 (25) | 2 (17) | 2 (17) | 2 (17) | 0.972 |
| **Moderate-to-heavy alcohol use** | | 10 (13.7) | 9 (12.3) | 18 (24.7) | 11 (15.1) | 25 (24.3) | **0.007** |
| **Hypertension** | | 106 (20.4) | 107 (20.6) | 104 (20.0) | 94 (18.1) | 109 (21.0) | 0.303 |
| **Atherosclerosis** | | 4 (8.9) | 9 (20.0) | 7 (15.6) | 11 (24.4) | 14 (31.1) | 0.141 |
| **Cerebrovascular disease** | | 26 (18.7) | 29 (20.9) | 32 (23.0) | 25 (18.0) | 27 (19.4) | 0.852 |
| **Diabetes** | | 35 (19.4) | 42 (23.3) | 31 (17.2) | 34 (18.9) | 38 (21.1) | 0.609 |
| **Ischemic heart disease or myocardial infarction** | | 37 (16.6) | 43 (19.3) | 43 (19.3) | 47 (21.1) | 53 (23.8) | 0.331 |
| **Malignant neoplasm** | | 49 (17.5) | 53 (18.9) | 62 (22.1) | 61 (21.8) | 55 (19.6) | 0.459 |
| **Total Cholesterol Trajectory Group** | **1** | 62 919.4) | 57 (17.9) | 64 (20.1) | 62 (19.4) | 74 (23.2) | 0.431 |
| | **2** | 64 (21.0) | 70 (23.0) | 62 (20.3) | 57 (18.7) | 52 (17.0) | |
| | **3** | 14 (18.9) | 12 (16.2) | 14 (18.9) | 20 (27.0) | 14 (18.9) | |
| **HDL-C Trajectory Group** | **1** | 65 (21.4) | 68 (22.4) | 53 (17.5) | 64 (21.1) | 53 (17.5) | 0.067 |
| | **2** | 50 (17.4) | 48 (16.7) | 60 (20.8) | 58 (20.1) | 72 (25.0) | |
| | **3** | 25 (23.4) | 23 (21.5) | 27 (25.2) | 17 (15.9) | 15 (14.0) | |
| **non-HDL-C Trajectory Group** | **1** | 57 (18.1) | 67 (21.3) | 63 (20.0) | 59 (18.7) | 69 (21.9) | 0.346 |
| | **2** | 75 (21.9) | 63 (18.4) | 73 (21.3) | 67 (19.5) | 65 (19.0) | |
| | **3** | 8 (20.0) | 9 (22.4) | 4 (10.0) | 13 (32.5) | 6 (15.0) | |
| **LDL-C Trajectory Group** | **1** | 56 (19.6) | 50 (17.5) | 57 (19.9) | 57 (19.9) | 66 (23.1) | 0.427 |
| | **2** | 72 (21.2) | 73 (21.5) | 72 (21.2) | 62 (18.2) | 61 (17.9) | |

| | | | | | | | |
|---|---|---|---|---|---|---|---|
| | **3** | 12 (16.7) | 16 (22.2) | 11 (15.3) | 20 (27.8) | 13 (18.1) | |
| **ln(Triglycerides) Trajectory Group** | **1** | 54 (20.9) | 54 (20.9) | 49 (19.0) | 50 (19.4) | 51 (19.8) | 0.882 |
| | **2** | 58 (18.2) | 59 (18.5) | 71 (22.3) | 65 (20.4) | 66 (20.7) | |
| | **3** | 28 (23.1) | 26 (21.5) | 20 (16.5) | 24 (19.8) | 23 (19.1) | |
| **MCI Case/Control Cohort (116 cases, 435 controls)** | | | | | | | |
| **Characteristics**[a] | | **VIM Quintile**[b] | | | | | ***p***[c] |
| | | **1** | **2** | **3** | **4** | **5** | |
| **Number of cases/controls** | **controls** | 86 (19.8) | 91 (20.9) | 84 (19.3) | 86 (19.8) | 88 (20.2) | 0.832 |
| | **cases** | 24 (20.7) | 19 (16.4) | 26 (22.4) | 24 (20.7) | 23 (19.8) | |
| | **total** | 110 (20.0) | 110 (20.0) | 110 (20.0) | 110 (20.0) | 111 (20.2) | – |
| **Age, md (r)** | | 75 (51-98) | 76 (53-93) | 75 (53-92) | 76 (55-94) | 77 (51-94) | 0.680 |
| **BMI, md (r)** | | 27 (18-41) | 27 (16-43) | 26 (17-40) | 27 (18-52) | 26 (16-41) | 0.571 |
| **Female** | | 45 (17.3) | 50 (19.2) | 52 (20.0) | 56 (21.5) | 57 (21.9) | 0.516 |
| **Race** | **Caucasian** | 103 (20.4) | 97 (19.2) | 103 (20.4) | 100 (19.8) | 101 (20.0) | 0.808 |
| | **Black/African American** | 2 (17) | 2 (17) | 1 (8) | 3 (25) | 4 (33) | |
| | **Asian** | 3 (10) | 10 (35) | 5 (17) | 6 (21) | 5 (17) | |
| | **Other** | 2 (33) | 1 (17) | 1 (17) | 1 (17) | 1 (17) | |
| **Blood-cholesterol lowering medications used** | | 85 (19.4) | 85 (19.4) | 87 (19.9) | 92 (21.0) | 88 (20.1) | 0.770 |
| ***APOE* ε2 Alleles** | **0** | 96 (20.0) | 99 (20.6) | 98 (20.4) | 92 (19.1) | 96 (20.0) | 0.643 |
| | **1** | 13 (19.1) | 10 (14.7) | 12 (17.6) | 18 (26.5) | 15 (22.1) | |
| | **2** | 1 (50) | 1 (50) | 0 (0) | 0 (0) | 0 (0) | |
| ***APOE* ε4 Alleles** | **0** | 83 (21.3) | 71 (18.2) | 78 (20.0) | 83 (21.3) | 74 (19.0) | **0.022** |
| | **1** | 25 (17.0) | 39 (26.5) | 29 (19.7) | 25 (17.0) | 29 (19.7) | |

| | | | | | | | |
|---|---|---|---|---|---|---|---|
| | **2** | 2 (13) | 0 (0) | 3 (20) | 2 (13) | 8 (53) | |
| **Years of education** | **<12** | 3 (25) | 2 (17) | 3 (25) | 3 (25) | 1 (8) | 0.693 |
| | **12** | 15 (19.2) | 15 (19.2) | 15 (19.2) | 10 (12.8) | 23 (29.5) | |
| | **12-15** | 17 (16.2) | 22 (21.0) | 20 (19.0) | 25 (23.8) | 21 (20.0) | |
| | **≥16** | 75 (21.1) | 71 (19.9) | 72 (20.2) | 72 (20.2) | 66 (18.5) | |
| **Smoking** | | 3 (25) | 2 (17) | 3 (25) | 3 (25) | 1 (7) | 0.847 |
| **Moderate-to-heavy alcohol use** | | 11 (17.5) | 8 (12.7) | 8 (12.7) | 17 (27.0) | 19 (30.2) | 0.054 |
| **Hypertension** | | 79 (18.5) | 90 (21.1) | 79 (18.5) | 89 (20.8) | 90 (21.1) | 0.147 |
| **Atherosclerosis** | | 3 (8.1) | 4 (10.8) | 7 (18.9) | 12 (32.4) | 11 (29.7) | 0.052 |
| **Cerebrovascular disease** | | 16 (17.0) | 20 (21.3) | 16 917.0) | 27 (28.7) | 15 (15.9) | 0.170 |
| **Diabetes** | | 28 (17.3) | 38 (23.5) | 29 (17.9) | 41 (25.3) | 26 (16.0) | 0.095 |
| **Ischemic heart disease or myocardial infarction** | | 34 (16.8) | 34 (16.8) | 42 (20.7) | 52 (25.6) | 41 (20.2) | 0.073 |
| **Malignant neoplasm** | | 49 (21.0) | 46 (19.7) | 42 (18.0) | 48 (20.6) | 48 (20.6) | 0.891 |
| **Total Cholesterol Trajectory Group** | **1** | 28 (18.2) | 26 (16.9) | 27 (17.5) | 37 (24.0) | 36 (23.4) | 0.643 |
| | **2** | 59 (20.9) | 56 (19.9) | 59 (20.9) | 54 (19.2) | 54 (19.2) | |
| | **3** | 23 (20.0) | 28 (24.4) | 24 (20.9) | 19 (16.5) | 21 (18.3) | |
| **HDL-C Trajectory Group** | **1** | 70 (22.9) | 64 (20.9) | 59 (19.3) | 62 (20.3) | 51 (16.7) | 0.320 |
| | **2** | 31 (16.2) | 34 (17.7) | 39 (20.3) | 38 (19.8) | 50 (26.0) | |
| | **3** | 9 (17.0) | 12 (22.6) | 12 (22.6) | 10 (18.9) | 10 (18.9) | |
| **non-HDL-C Trajectory Group** | **1** | 34 (18.0) | 34 (18.0) | 32 (16.9) | 43 (22.8) | 46 (24.3) | 0.520 |
| | **2** | 58 (20.4) | 60 21.1) | 64 (22.5) | 51 (18.0) | 51 (18.0) | |
| | **3** | 18 (23.1) | 16 (20.5) | 14 (18.0) | 16 (20.5) | 14 (18.0) | |

| | | | | | | | |
|---|---|---|---|---|---|---|---|
| **LDL-C Trajectory Group** | **1** | 44 (17.7) | 49 (19.7) | 46 (18.5) | 56 (22.5) | 54 (21.7) | 0.636 |
| | **2** | 56 (22.0) | 51 (20.0) | 57 (22.4) | 46 (18.0) | 45 (17.7) | |
| | **3** | 10 (21.3) | 10 (21.3) | 7 (14.9) | 8 (17.0) | 12 (25.5) | |
| **ln(Triglycerides) Trajectory Group** | **1** | 36 (20.2) | 37 (20.8) | 35 (19.7) | 34 (19.1) | 36 (20.2) | 0.663 |
| | **2** | 53 (19.0) | 53 (19.0) | 55 (19.7) | 54 (19.4) | 64 (22.9) | |
| | **3** | 21 (22.3) | 20 (21.3) | 20 (21.3) | 22 (23.4) | 11 (11.7) | |

[a]Age, BMI and social and medical history information are reported for age at first symptom onset, in cases, or matching age, in controls.

[b]VIM quintile and HDL trajectory group were determined as described in the text using HDL levels obtained over the 11-year period prior to and including the year of onset, in cases, or matching age, in controls. Number (percent of row total); for age and body mass index (BMI), median (md) [range(r)].

[c]Kruskal-Wallis test for age and BMI, otherwise $\chi^2$ test; values <0.05 are in bold. An asterisk identifies *p* values remaining significant after Bonferroni adjustment.

**Table S5. Cohort characteristics by quintile of LDL-C variability independent of the mean (VIM).**

| AD Case/Control Cohort (215 cases, 483 controls) | | | | | | | |
|---|---|---|---|---|---|---|---|
| Characteristics[a] | | VIM Quintile[b] | | | | | $p$[c] |
| | | 1 | 2 | 3 | 4 | 5 | |
| Number of cases/controls | controls | 88 (18.2) | 96 (19.9) | 100 (20.7) | 99 (20.5) | 100 (20.7) | 0.468 |
| | cases | 52 (24.2) | 43 (20.0) | 40 (18.6) | 40 (18.6) | 40 (18.6) | |
| | total | 140 (20.1) | 139 (19.9) | 140 (20.1) | 139 (19.9) | 140 (20.1) | – |
| Age, md (r) | | 76 (52-92) | 77 (51-93) | 78 (52-92) | 78 (52-89) | 78 (52-94) | 0.413 |
| Body Mass Index (BMI), md (r) | | 26 (16-39) | 25 (16-39) | 26 (18-40) | 25 (17-38) | 26 (16-39) | 0.579 |
| Female | | 79 (22.2) | 68 (19.2) | 65 (18.3) | 65 (18.3) | 78 (22.0) | 0.258 |
| Race | Caucasian | 129 (20.0) | 131 (20.3) | 129 (20.0) | 128 (19.8) | 129 (19.8) | 0.159 |
| | Black/African American | 7 (35) | 4 920) | 1 (5) | 3 (15) | 5 (25) | |
| | Asian | 3 (10) | 4 914) | 7 (24) | 8 (28) | 7 (24) | |
| | Other | 1 (25) | 0 (0) | 3 (75) | 0 (0) | 0 (0) | |
| Blood-cholesterol lowering medications used | | 74 (14.2) | 85 (16.2) | 106 (20.3) | 122 (23.3) | 136 (26.0) | **< 0.001*** |
| *APOE* ε2 Alleles | 0 | 124 (20.1) | 117 (19.0) | 122 (19.8) | 127 (20.6) | 127 (20.6) | 0.509 |
| | 1 | 15 (19.5) | 21 (27.3) | 16 (20.8) | 12 (15.6) | 13 (16.9) | |
| | 2 | 1 (25) | 1 (25) | 2 (50) | 0 (0) | 0 (0) | |
| *APOE* ε4 Alleles | 0 | 88 (20.5) | 83 (19.4) | 88 (20.5) | 80 (18.6) | 90 (21.0) | 0.656 |
| | 1 | 39 (17.3) | 49 (21.8) | 45 (20.0) | 51 (22.7) | 41 (18.2) | |
| | 2 | 13 (29.6) | 7 (15.9) | 7 (15.9) | 8 (18.2) | 9 (20.4) | |
| | <12 | 5 (33) | 1 (7) | 2 (13) | 4 (27) | 3 (20) | 0.646 |

| | | | | | | | |
|---|---|---|---|---|---|---|---|
| **Years of education** | **12** | 20 (19.0) | 23 (21.9) | 22 (21.0) | 15 (14.3) | 25 (23.8) | |
| | **12-15** | 16 (14.2) | 25 (22.1) | 22 (19.5) | 26 (23.0) | 24 (21.2) | |
| | **≥16** | 99 (21.3) | 90 (19.4) | 94 (20.2) | 94 (20.2) | 88 (18.9) | |
| **Smoking** | | 3 (25) | 2 (17) | 4 (33) | 1 (8) | 2 (17) | 0.703 |
| **Moderate-to-heavy alcohol use** | | 13 (17.8) | 12 (16.4) | 14 (19.2) | 14 (19.2) | 20 (27.4) | 0.567 |
| **Hypertension** | | 95 (18.3) | 99 (19.0) | 108 (20.8) | 110 (21.2) | 108 (20.8) | 0.156 |
| **Atherosclerosis** | | 6 (13.3) | 5 (11.1) | 11 (24.4) | 11 (24.4) | 12 (26.7) | 0.291 |
| **Cerebrovascular disease** | | 24 (17.3) | 31 (22.3) | 33 (23.7) | 23 (16.60 | 28 (20.1) | 0.504 |
| **Diabetes** | | 17 (9.4) | 36 (20.0) | 38 (21.1) | 53 (29.4) | 36 (20.0) | < **0.001*** |
| **Ischemic heart disease or myocardial infarction** | | 30 (13.4) | 31 (13.9) | 51 (22.9) | 48 (21.5) | 63 (28.2) | < **0.001*** |
| **Malignant neoplasm** | | 59 (21.1) | 55 (19.6) | 61 (21.8) | 49 (17.5) | 56 (20.0) | 0.677 |
| **Total Cholesterol Trajectory Group** | **1** | 68 (21.3) | 74 (23.2) | 71 (22.3) | 61 (19.1) | 45 (14.1) | **0.025** |
| | **2** | 57 (18.7) | 55 (18.0) | 52 (17.0) | 63 (20.7) | 78 (25.6) | |
| | **3** | 15 (20.3) | 10 (13.5) | 17 (23.0) | 15 (20.3) | 17 (23.0) | |
| **HDL-C Trajectory Group** | **1** | 58 (19.1) | 57 (18.8) | 61 (20.1) | 68 (22.4) | 59 (19.5) | 0.414 |
| | **2** | 53 (18.4) | 57 (19.8) | 61 (21.2) | 53 (18.4) | 64 (22.2) | |
| | **3** | 29 *27.1) | 25 (23.4) | 18 (16.8) | 18 (16.8) | 17 (15.9) | |
| **non-HDL-C Trajectory Group** | **1** | 70 (22.2) | 71 (22.5) | 68 (21.6) | 53 (16.8) | 53 (16.8) | **0.021** |
| | **2** | 64 (18.7) | 65 (19.0) | 63 (18.4) | 79 (23.0) | 72 (21.0) | |
| | **3** | 6 (15.0) | 3 (7.5) | 9 (22.5) | 7 (17.5) | 15 (37.5) | |
| **LDL-C Trajectory Group** | **1** | 59 (20.6) | 64 (22.4) | 67 (23.4) | 54 (18.9) | 42 (14.7) | **0.042** |
| | **2** | 72 (21.2) | 62 (18.2) | 60 (17.6) | 65 (19.1) | 81 (23.8) | |

| | | | | | | | |
|---|---|---|---|---|---|---|---|
| | **3** | 9 (12.5) | 13 (18.1) | 13 (18.1) | 20 (27.8) | 17 (23.6) | |
| **ln(Triglycerides) Trajectory Group** | **1** | 57 (22.1) | 61 (23.60 | 44 (17.0) | 44 (17.0) | 52 (20.2) | 0.079 |
| | **2** | 59 (18.5) | 62 (19.4) | 72 (22.6) | 60 (18.8) | 66 (20.7) | |
| | **3** | 24 (19.8) | 16 (13.2) | 24 (19.8) | 35 (28.9) | 22 (18.2) | |
| **MCI Case/Control Cohort (116 cases, 435 controls)** | | | | | | | |
| **Characteristics**[a] | | **VIM Quintile**[b] | | | | | ***p***[c] |
| | | **1** | **2** | **3** | **4** | **5** | |
| **Number of cases/controls** | **controls** | 76 (17.5) | 87 (20.0) | 89 (20.5) | 87 (20.0) | 96 (22.1) | **0.033** |
| | **cases** | 34 (29.3) | 23 (19.8) | 21 (18.1) | 23 (19.8) | 15 (12.9) | |
| | **total** | 110 (20.0) | 110 (20.0) | 110 (20.0) | 110 (20.0) | 111 (20.2) | – |
| **Age, md (r)** | | 75 (51-98) | 76 (51-92) | 77 (52-93) | 77 (53-94) | 76 (55-94) | 0.276 |
| **BMI, md (r)** | | 26 (17-40) | 26 (18-46) | 26 (17-52) | 27 (16-44) | 27 (16-42) | 0.566 |
| **Female** | | 47 (18.1) | 45 (17.3) | 52 (20.0) | 54 (20.8) | 62 (23.8) | 0.190 |
| **Race** | **Caucasian** | 109 (21.6) | 100 (19.7) | 97 (19.2) | 100 (19.7) | 98 (19.4) | **0.001*** |
| | **Black/African American** | 1 (8) | 1 (8) | 4 (33) | 1 (8) | 5 (42) | |
| | **Asian** | 0 (0) | 8 (27.6) | 4 (13.8) | 9 (31.0) | 8 (27.6) | |
| | **Other** | 0 (0) | 1 (17) | 5 (83) | 0 (0) | 0 (0) | |
| **Blood-cholesterol lowering medications used** | | 64 (14.6) | 82 (18.8) | 81 (18.5) | 100 (22.9) | 110 (25.2) | **< 0.001*** |
| ***APOE* ε2 Alleles** | **0** | 99 (20.6) | 94 (19.5) | 93 (19.3) | 99 (20.6) | 96 (20.0) | 0.642 |
| | **1** | 11 (16.2) | 16 (23.5) | 17 (25.0) | 10 (14.7) | 14 (20.6) | |
| | **2** | 0 (0) | 0 (0) | 0 (0) | 1 (50) | 1 (50) | |
| ***APOE* ε4 Alleles** | **0** | 79 (20.3) | 80 (20.6) | 76 (19.5) | 76 (19.5) | 78 (20.0) | 0.926 |
| | **1** | 28 (19.0) | 29 (19.7) | 31 (21.1) | 31 (21.1) | 28 (19.0) | |

| | | | | | | | |
|---|---|---|---|---|---|---|---|
| | **2** | 3 (20) | 1 (7) | 3 (20) | 3 (20) | 5 (33) | |
| **Years of education** | **<12** | 1 (8) | 1 (8) | 3 (25) | 3 (25) | 4 (33) | 0.272 |
| | **12** | 17 (21.8) | 9 (11.5) | 17 (21.8) | 17 (21.8) | 18 (23.1) | |
| | **12-15** | 13 (12.4) | 24 (22.9) | 19 (18.1) | 22 (21.0) | 27 (25.7) | |
| | **≥16** | 79 (22.2) | 76 (21.4) | 71 (19.9) | 68 (19.1) | 62 (17.4) | |
| **Smoking** | | 2 (17) | 1 (8) | 4 (33) | 2 (17) | 3 (25) | 0.698 |
| **Moderate-to-heavy alcohol use** | | 11 (17.5) | 11 (17.5) | 12 (19.0) | 14 (22.2) | 15 (23.8) | 0.889 |
| **Hypertension** | | 74 (17.3) | 79 (18.5) | 84 919.7) | 91 (21.3) | 99 (23.2) | **0.001*** |
| **Atherosclerosis** | | 6 (16.2) | 3 (8.1) | 6 (16.2) | 12 (32.4) | 10 (27.0) | 0.117 |
| **Cerebrovascular disease** | | 18 (19.2) | 18 (19.2) | 13 (13.8) | 26 (27.7) | 19 (20.2) | 0.233 |
| **Diabetes** | | 20 (12.4) | 27 (16.7) | 35 (21.6) | 42 (25.9) | 38 (23.5) | **0.009** |
| **Ischemic heart disease or myocardial infarction** | | 33 (16.3) | 40 (19.7) | 34 (16.8) | 50 (24.6) | 46 (22.7) | 0.078 |
| **Malignant neoplasm** | | 52 (22.3) | 43 (18.4) | 47 (20.2) | 45 (19.3) | 46 (19.7) | 0.789 |
| **Total Cholesterol Trajectory Group** | **1** | 41 (26.6) | 32 (20.8) | 30 (19.5) | 30 (19.5) | 21 (13.6) | 0.091 |
| | **2** | 54 (19.2) | 59 (20.9) | 52 (18.4) | 56 (19.9) | 61 (21.6) | |
| | **3** | 15 (13.0) | 19 (16.5) | 28 (24.4) | 24 (20.9) | 29 (25.2) | |
| **HDL-C Trajectory Group** | **1** | 58 (19.0) | 62 (20.3) | 67 (21.9) | 57 (19.6) | 62 (20.3) | 0.435 |
| | **2** | 40 (20.8) | 40 (20.8) | 38 (19.8) | 37 (19.3) | 38 (19.3) | |
| | **3** | 12 (22.6) | 8 (15.1) | 5 (9.4) | 16 (30.2) | 12 (22.6) | |
| **non-HDL-C Trajectory Group** | **1** | 50 (26.5) | 42 (22.2) | 33 (17.5) | 36 (19.0) | 38 (14.8) | **0.018** |
| | **2** | 51 (18.0) | 55 (19.4) | 55 (19.4) | 62 (21.8) | 61 (21.5) | |
| | **3** | 9 (11.5) | 13 (16.7) | 22 (28.2) | 12 (15.4) | 22 (28.2) | |

| | | | | | | | |
|---|---|---|---|---|---|---|---|
| **LDL-C Trajectory Group** | **1** | 60 (24.1) | 51 (20.5) | 44 (17.7) | 57 (22.9) | 37 (14.9) | **0.020** |
| | **2** | 46 (18.0) | 51 (20.0) | 51 (20.0) | 45 (17.6) | 62 (24.3) | |
| | **3** | 4 (8.5) | 8 (17.0) | 15 (31.9) | 8 (17.0) | 12 (25.5) | |
| **ln(Triglycerides) Trajectory Group** | **1** | 43 (24.2) | 39 (21.9) | 29 (16.3) | 36 (20.2) | 31 (17.4) | 0.139 |
| | **2** | 57 (20.4) | 53 (19.0) | 61 (21.9) | 55 (19.7) | 53 (19.0) | |
| | **3** | 10 (10.6) | 18 (19.2) | 20 (21.3) | 19 (20.2) | 27 (28.7) | |

[a]Age, BMI and social and medical history information are reported for age at first symptom onset, in cases, or matching age, in controls.

[b]VIM quintile and LDL-C trajectory group were determined as described in the text using LDL-C levels obtained over the 11-year period prior to and including the year of onset, in cases, or matching age, in controls. Number (percent of row total); for age and body mass index (BMI), median (md) [range(r)].

[c]Kruskal-Wallis test for age and BMI, otherwise $\chi^2$ test; values <0.05 are in bold. An asterisk identifies *p* values remaining significant after Bonferroni adjustment.

**Table S6. Cohort characteristics by quintile of ln(triglycerides) variability independent of the mean (VIM).**

| AD Case/Control Cohort (215 cases, 483 controls) | | | | | | | |
|---|---|---|---|---|---|---|---|
| **Characteristics**[a] | | **VIM Quintile**[b] | | | | | ***p***[c] |
| | | **1** | **2** | **3** | **4** | **5** | |
| **Number of cases/controls** | **controls** | 90 (18.6) | 103 (21.3) | 100 (20.7) | 95 (19.7) | 95 (19.7) | 0.456 |
| | **cases** | 50 (23.3) | 36 (16.7) | 40 (18.6) | 44 (20.5) | 45 (20.9) | |
| | **total** | 140 (20.1) | 139 (19.9) | 140 (20.1) | 139 (19.9) | 140 (20.1) | – |
| **Age, md (r)** | | 77 (52-92) | 78 (52-93) | 77 (51-92) | 78 (51-94) | 77 (52-93) | 0.979 |
| **Body Mass Index (BMI), md (r)** | | 25 (16-39) | 26 (16-38) | 26 (19-40) | 26 (17-39) | 25 (16-39) | 0.821 |
| **Female** | | 80 (22.5) | 83 (20.6) | 74 (20.8) | 71 (20.0) | 57 (16.1) | 0.079 |
| **Race** | **Caucasian** | 133 (20.6) | 123 (19.1) | 140 (20.2) | 129 (20.0) | 130 (20.2) | 0.687 |
| | **Black/African American** | 3 (15) | 5 (25) | 5 (25) | 3 (15) | 4 (20) | |
| | **Asian** | 4 (14) | 10 (34) | 5 (17) | 6 (21) | 4 (14) | |
| | **Other** | 0 (0) | 1 (25) | 0 (0) | 1 (25) | 2 (50) | |
| **Blood-cholesterol lowering medications used** | | 103 (19.7) | 108 (20.6) | 108 (20.6) | 103 (19.7) | 101 (19.3) | 0.791 |
| ***APOE* ε2 Alleles** | **0** | 127 (20.6) | 121 (19.6) | 126 920.4) | 121 (19.6) | 122 (19.8) | 0.924 |
| | **1** | 12 (15.6) | 17 (22.1) | 13 (16.9) | 18 (23.4) | 17 (22.1) | |
| | **2** | 1 (25) | 1 (25) | 1 (25) | 0 (0) | 1 (25) | |
| ***APOE* ε4 Alleles** | **0** | 82 (19.1) | 85 (19.8) | 86 (20.0) | 85 (19.8) | 91 (21.2) | 0.852 |
| | **1** | 48 (21.3) | 49 (21.8) | 43 (19.1) | 45 (20.0) | 40 (17.8) | |
| | **2** | 10 (22.7) | 5 (11.4) | 11 (25.0) | 9 (20.4) | 9 (20.4) | |
| | **<12** | 0 (0) | 2 (13) | 7 (47) | 3 (20) | 3 (20) | **0.034** |

| | | | | | | | |
|---|---|---|---|---|---|---|---|
| **Years of education** | **12** | 23 (21.9) | 30 (28.6) | 14 (13.3) | 13 (13.3) | 24 (22.9) | |
| | **12-15** | 24 (21.2) | 20 (17.7) | 29 (25.7) | 23 (20.4) | 17 (15.0) | |
| | **≥16** | 93 (20.0) | 87 (18.7) | 90 (19.4) | 99 (21.3) | 96 (20.6) | |
| **Smoking** | | 3 (25) | 4 933) | 1 98) | 3 (25) | 1 (8) | 0.544 |
| **Moderate-to-heavy alcohol use** | | 11 (15.1) | 12 (16.4) | 16 (21.9) | 18 (24.7) | 16 (21.9) | 0.608 |
| **Hypertension** | | 100 (19.2) | 106 (20.4) | 114 (21.8) | 102 (19.6) | 98 (18.8) | 0.196 |
| **Atherosclerosis** | | 5 (11.1) | 8 (17.8) | 10 (22.2) | 14 (31.1) | 8 (17.8) | 0.258 |
| **Cerebrovascular disease** | | 29 (20.9) | 29 (20.9) | 32 (23.0) | 20 (14.4) | 29 (20.9) | 0.461 |
| **Diabetes** | | 32 (17.8) | 47 (26.1) | 36 (20.0) | 36 (20.0) | 29 (16.1) | 0.126 |
| **Ischemic heart disease or myocardial infarction** | | 36 (16.1) | 52 (23.3) | 52 (23.3) | 40 (17.9) | 43 (19.3) | 0.142 |
| **Malignant neoplasm** | | 45 (16.1) | 62 (22.1) | 68 (24.3) | 45 (16.1) | 60 (21.4) | **0.012** |
| **Total Cholesterol Trajectory Group** | **1** | 50 (15.7) | 68 (21.3) | 65 (20.4) | 70 (21.9) | 66 (20.7) | **0.048** |
| | **2** | 78 (25.6) | 58 (19.0) | 54 (17.7) | 52 (17.0) | 63 (20.7) | |
| | **3** | 12 (16.2) | 13 (17.6) | 21 (28.4) | 17 (23.0) | 11 (14.9) | |
| **HDL-C Trajectory Group** | **1** | 53 (17.5) | 67 (22.1) | 55 (18.2) | 58 (19.1) | 70 (23.1) | 0.191 |
| | **2** | 60 (20.8) | 47 (16.3) | 65 (22.6) | 63 (21.9) | 53 (18.4) | |
| | **3** | 27 (25.2) | 25 (23.4) | 20 (18.7) | 18 (16.8) | 17 (15.9) | |
| **non-HDL-C Trajectory Group** | **1** | 51 (16.2) | 77 (24.4) | 61 (19.4) | 64 (20.3) | 62 (19.7) | 0.093 |
| | **2** | 81 (23.6) | 53 (15.4) | 71 (20.7) | 65 (19.0) | 73 (21.3) | |
| | **3** | 8 (20.0) | 9 (22.5) | 8 (20.0) | 10 (25.0) | 5 (12.5) | |
| **LDL-C Trajectory Group** | **1** | 40 (14.0) | 68 (23.8) | 59 (20.6) | 60 (21.0) | 59 (20.6) | 0.108 |
| | **2** | 82 (24.1) | 59 (17.4) | 66 (19.4) | 66 (19.4) | 67 (19.7) | |

| | | | | | | | |
|---|---|---|---|---|---|---|---|
| | **3** | 18 (25.0) | 12 (16.7) | 15 (20.8) | 13 (18.1) | 14 (19.4) | |
| **ln(Triglycerides) Trajectory Group** | **1** | 62 (24.0) | 55 (21.3) | 43 916.7) | 45 (17.4) | 53 (20.5) | 0.445 |
| | **2** | 54 (16.9) | 61 (19.1) | 71 (22.3) | 68 (21.3) | 65 (20.4) | |
| | **3** | 24 (19.8) | 23 (19.0) | 26 (21.5) | 26 (21.5) | 22 (18.2) | |
| **MCI Case/Control Cohort (116 cases, 435 controls)** | | | | | | | |
| **Characteristics**[a] | | **VIM Quintile**[b] | | | | | ***p***[c] |
| | | **1** | **2** | **3** | **4** | **5** | |
| **Number of cases/controls** | **controls** | 85 (19.5) | 92 (21.2) | 89 (20.5) | 87 (20.0) | 82 (18.9) | 0.460 |
| | **cases** | 25 (21.6) | 18 (15.5) | 21 (18.1) | 23 (19.8) | 29 (25.0) | |
| | **total** | 110 (20.0) | 110 (20.0) | 110 (20.0) | 110 (20.0) | 111 (20.2) | – |
| **Age, md (r)** | | 76 (52-92) | 75 (51-98) | 77 (53-91) | 76 (51-94) | 76 (53-93) | 0.704 |
| **BMI, md (r)** | | 27 (17-46) | 27 (18-43) | 27 (17-52) | 26 (17-44) | 26 (16-41) | 0.873 |
| **Female** | | 58 (22.3) | 53 (20.4) | 48 (19.5) | 54 (20.8) | 47 (18.1) | 0.531 |
| **Race** | **Caucasian** | 104 (20.6) | 101 (20.0) | 100 (19.8) | 99 (19.6) | 100 (19.8) | 0.552 |
| | **Black/African American** | 1 (8) | 1 (8) | 3 (25) | 3 (25) | 4 (33) | |
| | **Asian** | 2 (6.9) | 7 (24.1) | 6 (20.7) | 8 (27.6) | 6 (20.7) | |
| | **Other** | 3 (50) | 1 (17) | 1 (16) | 0 (0) | 1 (17) | |
| **Blood-cholesterol lowering medications used** | | 84 (19.2) | 87 (19.9) | 89 (20.4) | 87 (19.9) | 90 (20.6) | 0.914 |
| ***APOE* ε2 Alleles** | **0** | 101 (21.0) | 94 (19.5) | 93 (19.3) | 96 (19.7) | 97 (20.2) | 0.535 |
| | **1** | 8 (1.8) | 16 (23.5) | 17 (25.0) | 14 (20.6) | 13 (19.1) | |
| | **2** | 1 (50) | 0 (0) | 0 (0) | 0 (0) | 1 (50) | |
| ***APOE* ε4 Alleles** | **0** | 75 (19.3) | 75 (19.3) | 83 (21.3) | 78 (20.0) | 78 (20.0) | 0.917 |
| | **1** | 31 (21.1) | 32 (21.8) | 26 (17.7) | 29 (19.7) | 29 (19.7) | |

| | | | | | | | |
|---|---|---|---|---|---|---|---|
| | **2** | 4 (27) | 3 (20) | 1 (7) | 3 (20) | 4 (27) | |
| **Years of education** | **<12** | 4 (33) | 0 (0) | 4 (33) | 1 (8) | 3 (25) | 0.210 |
| | **12** | 15 (19.2) | 15 (19.2) | 12 (15.4) | 15 (19.2) | 21 (26.9) | |
| | **12-15** | 28 (36.7) | 18 (18.1) | 25 (23.8) | 16 (15.2) | 18 (17.1) | |
| | **≥16** | 63 (17.7) | 77 (21.6) | 69 (19.4) | 78 (21.9) | 69 (19.4) | |
| **Smoking** | | 2 (17) | 2 (17) | 4 (33) | 2 (17) | 2 (17) | 0.849 |
| **Moderate-to-heavy alcohol use** | | 9 (14.3) | 13 (20.6) | 12 (19.0) | 13 (20.6) | 16 (25.4) | 0.702 |
| **Hypertension** | | 78 (18.3) | 85 (19.9) | 95 (22.2) | 86 (20.1) | 83 (19.4) | 0.085 |
| **Atherosclerosis** | | 6 (16.2) | 9 (24.3) | 3 (8.1) | 12 (32.4) | 7 (18.9) | 0.161 |
| **Cerebrovascular disease** | | 17 (19.1) | 23 (24.5) | 13 (13.8) | 22 (23.4) | 19 (20.2) | 0.385 |
| **Diabetes** | | 35 (21.6) | 36 (22.2) | 30 (18.5) | 37 (22.8) | 24 (14.8) | 0.256 |
| **Ischemic heart disease or myocardial infarction** | | 39 (19.2) | 43 (21.2) | 34 (16.8) | 47 (23.2) | 40 (19.7) | 0.454 |
| **Malignant neoplasm** | | 40 (17.2) | 53 (22.8) | 51 (21.9) | 53 (22.8) | 36 (15.4) | **0.043** |
| **Total Cholesterol Trajectory Group** | **1** | 27 (17.5) | 36 923.4) | 29 (18.8) | 32 (20.8) | 30 (19.5) | 0.389 |
| | **2** | 62 (22.0) | 54 (19.2) | 63 (22.3) | 53 (18.8) | 50 (17.7) | |
| | **3** | 21 (18.3) | 20 (17.4) | 18 (15.6) | 24 (21.7) | 31 (27.0) | |
| **HDL-C Trajectory Group** | **1** | 62 (20.3) | 60 (19.6) | 62 (20.3) | 62 (20.3) | 60 (19.6) | 0.988 |
| | **2** | 36 (18.8) | 38 (19.8) | 39 (20.3) | 40 (20.8) | 39 (20.3) | |
| | **3** | 12 (22.6) | 12 (22.6) | 9 (17.0) | 8 (15.1) | 12 (22.6) | |
| **non-HDL-C Trajectory Group** | **1** | 32 (16.9) | 50 (26.5) | 34 (18.0) | 42 (22.2) | 31 (16.4) | **0.014** |
| | **2** | 66 (23.2) | 47 (16.6) | 65 (22.9) | 48 (16.9) | 58 (20.4) | |
| | **3** | 12 (15.4) | 11 (16.7) | 11 (14.1) | 20 (25.6) | 22 (28.2) | |

| | | | | | | | |
|---|---|---|---|---|---|---|---|
| **LDL-C Trajectory Group** | **1** | 42 (16.9) | 64 (25.7) | 50 (20.1) | 53 (21.3) | 40 (16.1) | **0.036** |
| | **2** | 57 (22.4) | 40 (15.7) | 54 (21.2) | 46 (18.0) | 58 (22.8) | |
| | **3** | 1 (23.4) | 6 (12.8) | 6 (12.8) | 11 (23.4) | 13 (27.7) | |
| **ln(Triglycerides) Trajectory Group** | **1** | 40 (22.5) | 39 (21.9) | 30 (16.8) | 23 (19.1) | 35 (19.7) | 0.897 |
| | **2** | 50 (17.9) | 55 (19.7) | 59 (21.1) | 57 (20.4) | 58 (20.8) | |
| | **3** | 20 (21.3) | 16 (17.0) | 21 (22.3) | 19 (20.2) | 18 (19.2) | |

[a]Age, BMI and social and medical history information are reported for age at first symptom onset, in cases, or matching age, in controls.

[b]VIM quintile and ln(triglycerides) trajectory group were determined as described in the text using triglyceride levels obtained over the 11-year period prior to and including the year of onset, in cases, or matching age, in controls. Number (percent of row total); for age and body mass index (BMI), median (md) [range(r)].

[c]Kruskal-Wallis test for age and BMI, otherwise $\chi^2$ test; values <0.05 are in bold. An asterisk identifies *p* values remaining significant after Bonferroni adjustment.

**Figure S1**. Lipid measurements in the AD and MCI case-control cohorts.

Group-based trajectory models were developed using lipid measurements obtained a decade prior to the year of onset (cases) or year of age-match (controls). Panels **A** (AD) and **C** (MCI) show the percentages of cases and controls who had lipid measurements each year. Study inclusion criteria required subjects to have at least three lipid measurements. Panels **B** (AD) and **D** (MCI) show how the number of lipid measurements was distributed in cases and controls.

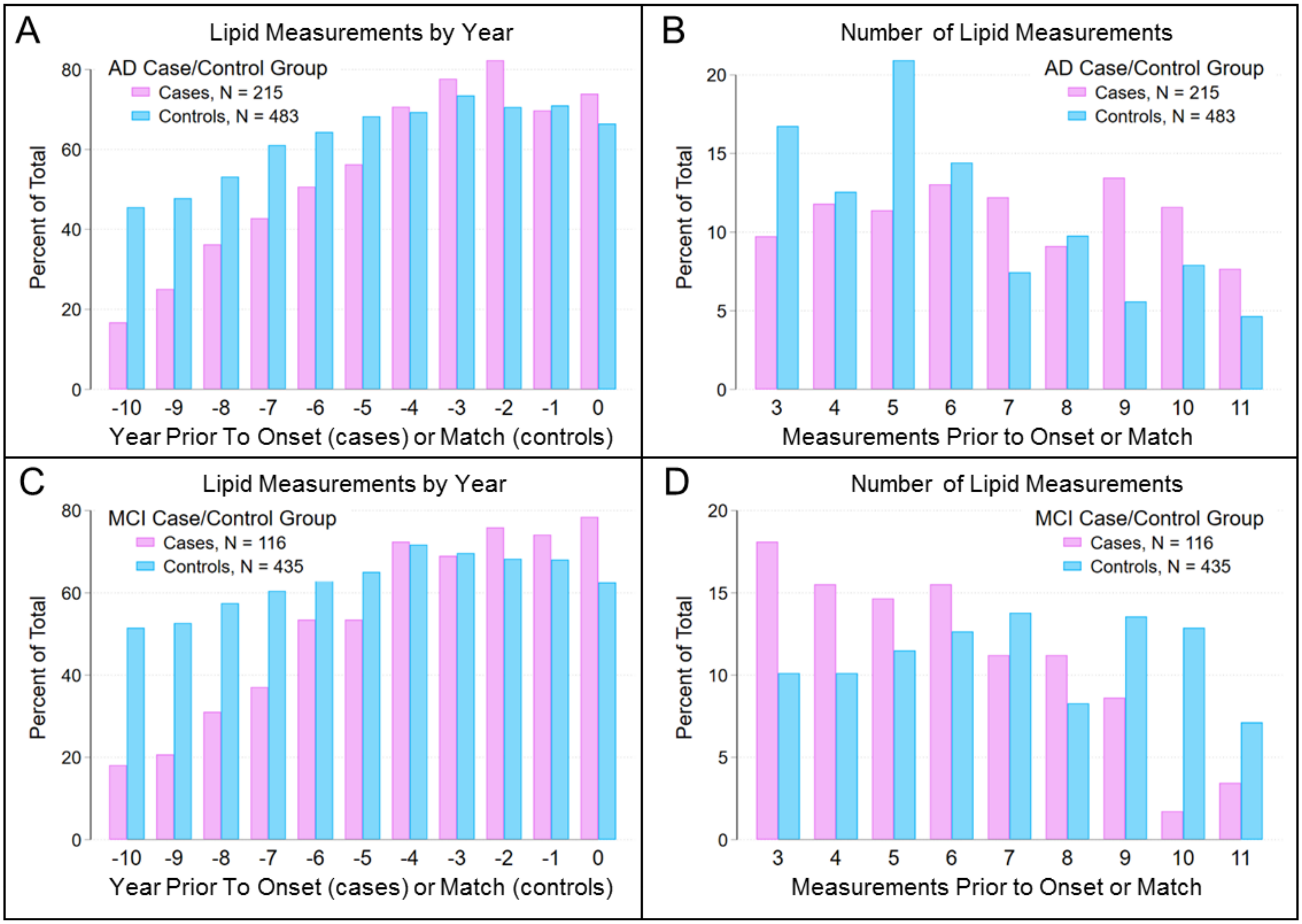
